# Neurotransmission-Related Gene Expression in the Frontal Pole (Brodmann Area 10) is Altered in Subjects with Bipolar Disorder and Schizophrenia

**DOI:** 10.1101/2022.06.03.22275600

**Authors:** Adriana M. Medina, Megan Hastings Hagenauer, David M. Krolewski, Evan Hughes, Liam Cannon Thew Forrester, David M. Walsh, Maria Waselus, Evelyn Richardson, Cortney A. Turner, P. Adolfo Sequeira, Preston M. Cartagena, Robert C. Thompson, Marquis P. Vawter, Blynn G. Bunney, Richard M. Myers, Jack D. Barchas, Francis S.Y. Lee, Alan F. Schatzberg, William E. Bunney, Huda Akil, Stanley J. Watson

## Abstract

Brodmann Area 10 (BA10) is the largest cytoarchitectonic region of the human cortex, performing complex integrative functions. BA10 undergoes intensive adolescent grey matter pruning around the average age of onset for Bipolar disorder (BP) and Schizophrenia (SCHIZ), and its dysfunction is likely to underly aspects of their shared symptomology. In this study, we investigated the role of BA10 neurotransmission-related gene expression in BP and SCHIZ. We performed qPCR to measure the expression of 115 neurotransmission-related targets in control, BP, and SCHIZ post-mortem samples (*n*=72). We chose this method for its high sensitivity to detect low-level expression. We then bolstered our findings by performing a meta-analysis of publicly-released BA10 microarray data (*n*=101) and identified sources of convergence with our qPCR results. To improve interpretation, we compiled an unusually large database of clinical metadata for our samples. We used this data to explore the relationship between BA10 gene expression, therapeutics, substances of abuse, and symptom profiles, and validated these findings with publicly-available datasets. Using these convergent sources of evidence, we identified 20 neurotransmission-related genes that were differentially expressed in BP and SCHIZ in BA10. These results included a large diagnosis-related decrease in two important therapeutic targets with low-levels of expression, HTR2B and DRD4, as well as other findings related to dopaminergic, GABA-ergic and astrocytic function. We also observed that therapeutics may produce differential expression that opposes the effects of diagnosis. In contrast, substances of abuse showed similar effects on BA10 gene expression as BP and SCHIZ, potentially amplifying diagnosis-related dysregulation.

## Introduction

The frontal pole (Brodmann area 10, BA10) is the largest cytoarchitectonic region of the human cortex^1^ and perhaps one of the most evolutionarily advanced. Relative to other apes, human BA10 occupies twice the proportion of the total cortical extension and has much lower cellular density^2,3^. BA10 has a larger number of dendritic spines per cell compared to adjoining areas^4^ and extensive connections with higher order association areas^5^, suggesting high levels of input integration.

During adolescence and young adulthood, these connections are refined during an intensive developmental pruning period, producing large decreases in BA10 cortical thickness^6^. This developmental pruning coincides with the average age of onset of Bipolar Disorder (BP) and Schizophrenia (SCHIZ)^7^, and may be dysregulated in association with psychiatric illness^8^. By adulthood, individuals with both SCHIZ and BP show reduced BA10 grey matter thickness compared to control (CTRL) subjects^8,9^, and reduced BA10 cortical surface area^9^, especially in association with psychosis^8^ and antipsychotic medication^9^. Functionally, subjects with BP and SCHIZ have heightened BA10 resting state activity^10,11^, as well as altered functional connectivity to sensory/association areas and subcortical regions^12,13^.

The role of BA10 dysfunction in psychiatric symptomology is unclear. In general, BA10 is associated with complex cognitive functions that depend on maintaining mental representations of alternative courses of action^5^, such as multi-tasking, prospective memory, time estimation, decision making, task-switching, and flexible emotional control^1,5^. Accordingly, BA10 abnormalities in SCHIZ and BP have been linked to deficits in working memory^11^, cognitive control^14^, and impulse control^15,16^. BA10 also allows for the integration of internally- and externally-obtained cognition^1^, suggesting a potential role in psychotic symptoms including delusions, hallucinations, and disorganized speech.

For the present study, we examined neurotransmission-related gene expression in BA10 in subjects with BP and SCHIZ, due to its relevance to both disease etiology and the design of therapeutics^17–19^. We chose the highly-sensitive method of qPCR, allowing us to both confirm and expand on previous studies tackling the topic with gene expression profiling^20–24^, and targeted 115 transcripts related to the main neurotransmitter systems in the frontal cortex: glutamate, GABA, dopamine, and serotonin.^25^ We then augmented our findings with an a meta-analysis of publicly-available microarray data^20,21^ and an in-depth examination of accompanying clinical characteristics to clarify the role of BA10 in the shared symptomology of the two disorders^26^.

## Methods

This research was overseen and approved by the University of Michigan Institutional Review Board, Pritzker Neuropsychiatric Disorders Research Consortium, and the University of California Irvine Institutional Review Board. Key Resources (**Table S1**) and full methodological details are documented in the supplement.

Human samples were collected through the University of California-Irvine Pritzker Brain Donor Program with informed consent from next of kin (n=72, CTRL: *n*=27, BPD: *n*=21, SCHIZ: *n*=24; **Table S2, Fig S1**^27^). A detailed psychological autopsy was performed using coroner records, medical records, and interviews with next-of-kin (**Appendix 1)**. This information was used to confirm BP and SCHIZ diagnosis (diagnosis criteria:^26^), and ensure absence of neurological or psychiatric disorder in CTRL subjects or their first-degree relatives. Other clinical information was summarized as 49 exploratory variables (**Fig S2**) denoting the presence or absence of 1) medication, 2) exposure to alcohol or drugs of abuse, and 3) diagnosis-related symptoms.

Brains were extracted during autopsy and kept on ice until being sliced into 1 cm thick coronal slabs, then snap-frozen for storage (−80°C) until microdissection. After counterbalancing processing batches by diagnosis, samples were blinded, and the foremost rostral slab from the left hemisphere was sub-dissected to obtain blocks averaging 500 µg containing lateral BA10 (**Fig S3**), a subregion implicated in SCHIZ^13^. RNA was extracted using TRIzol(tm) and purified (RNeasy® Mini Kit). cDNA was synthesized (iScript Reverse Transcription Supermix kit) and analyzed in duplicate via qPCR (Applied Biosystems ViiA 7 real time PCR system) using two sets of ThermoFisher Scientific Taqman Gene Expression Array qPCR cards: 1) “Human GABA Glutamate” (REF#4342259): 84 targets (12 reference genes), 2) “Dopamine Serotonin” (REF#4342253): 31 targets (17 reference genes) (**Table S3**). These cards were preloaded with a complete list of targets for the main neurotransmitter systems in the frontal cortex: glutamate, GABA, dopamine, and serotonin^25^, including receptors, transporters, metabolic enzymes, and other associated molecules. The cards were further customized to include several well-known markers for interneuron subtypes (SST, PVALB, CALB1) and astrocytes (AQP4, GJA1, GFAP, KCNJ10, S100B) to enhance the interpretation of neurotransmission-related data. Prior to differential expression analysis, the qPCR quantification cycle (Cq) data for targets was normalized using the average reference gene expression for each sample to produce -ΔCq values^28^.

For comparison, we performed a meta-analysis of publicly-released BA10 microarray data (re-annotated and re-analyzed: Gene Expression Omnibus #GSE12654^20^: *n*=50, CTRL: n=15, BPD: n=11, SCHIZ: n=13, MDD=11; #GSE17612^21^: *n*=51, CTRL: n=23, SCHIZ: n=28) using random effects modeling^29^ of the Log(2) Fold Change (Log2FC) and sampling variance (standard error(SE)^2^) for each gene (EntrezID) from the *limma*^30^ differential expression output for each dataset.

Both qPCR and microarray differential expression analyses controlled for common sources of biological and technical noise (*all:* pH, PMI, age, gender; *qPCR:* RIN, RNA concentration, Card; *microarray* (when applicable): RNA degradation, Rate of Death, Scan Date) and corrected for false discovery rate (FDR: Benjamini-Hochberg method^31^). All analysis code (Rstudio v1.0.153, R v3.4.1) has been released on Github (*https://github.com/hagenaue/Adriana_FrontalPole, https://github.com/hagenaue/FrontalPole_Microarray*). All statistical tests were performed using two-sided p-value calculations. When possible, findings were directly compared to relevant published datasets^23,23,32–42^ to determine validity and generalizability.

## Results

To evaluate the effects of BP and SCHIZ on neurotransmission-related gene expression in BA10 using highly sensitive methodology, we ran all samples in duplicate on two custom sets of Taqman qPCR cards loaded with 115 targets related to the main neurotransmitter systems in the frontal cortex^25^. We then performed a meta-analysis of previous BA10 microarray studies^20,21^ and used the convergence between our qPCR and microarray results to identify additional differentially-expressed neurotransmission-related genes. To explore the function of these differentially-expressed genes, we compared our findings across diagnosis categories, and explored the similarity between our findings and those from neighboring cortical areas. We then leveraged the unusually large database of metadata associated with our samples to explore the relationship between gene expression and clinical characteristics, using published data to bolster exploratory findings.

A summary of the main findings is presented below. Comprehensive details are in the Supplement, including all de-identifiable sample metadata (64 technical and clinical variables, **Tables S2**) and complete statistical reporting for the full concatenated qPCR and microarray results (**Tables S5, S8)**. The full qPCR datasets (Cq) and normalized results (-ΔCq) have been released on FigShare (*Link TBA*) with associated metadata.

### qPCR Results

#### Balanced Design

Tissue was collected from 72 subjects. Three subjects were later excluded due to poor RNA quality metrics, leaving *n*=69 (CTRL: *n*=26, BP: *n*=21, SCHIZ: *n*=22). Subjects were predominantly Caucasian (91%) and male (88%), although a quarter of the BP group was female (*Diagnosis vs. Sex: p*=0.0174, **Fig S5**). The diagnosis groups were otherwise balanced in regard to critical biological and technical variables (**Table S4, Figs S4-S5**, *p*>0.10: age, brain pH, post-mortem interval (PMI), tissue block weight, RNA concentration, purity (260/280, 260/230), and integrity (RNA integrity number (RIN), 28S/18S). All subjects had experienced a fast death (agonal factor score=0).

#### Effect of Diagnosis

We characterized the effect of diagnosis on the expression of 111 neurotransmission-related target genes while controlling for sources of biological and technical noise (pH, PMI, age, gender, RIN, RNA concentration, Card). As expected, qPCR accurately measured the expression of target genes with extremely low levels of expression (quantification cycles (Cq) between 27-34.6) without decreased accuracy (**Fig S13A-C, Table S3**). This allowed us to accurately characterize the relationship between diagnosis and the expression of neurotransmission-related genes that were not reliably quantifiable within previous microarray and RNA-Seq studies (**Fig S13D-G**).

Indeed, two of our strongest findings were monoamine receptor genes with very low levels of cortical expression: 5-Hydroxytryptamine Receptor 2B (HTR2B, average Cq: 31.8) and Dopamine Receptor D4 (DRD4, average Cq: 33.6). Both of these genes showed a relationship with diagnosis that survived false discovery rate correction (FDR<0.10) due to decreased expression in both BP and SCHIZ (**Fig 1A-B)**. Notably, the magnitude of the decrease in HTR2B in SCHIZ was strikingly large - almost a full halving of normal expression levels.

**Figure 1.**
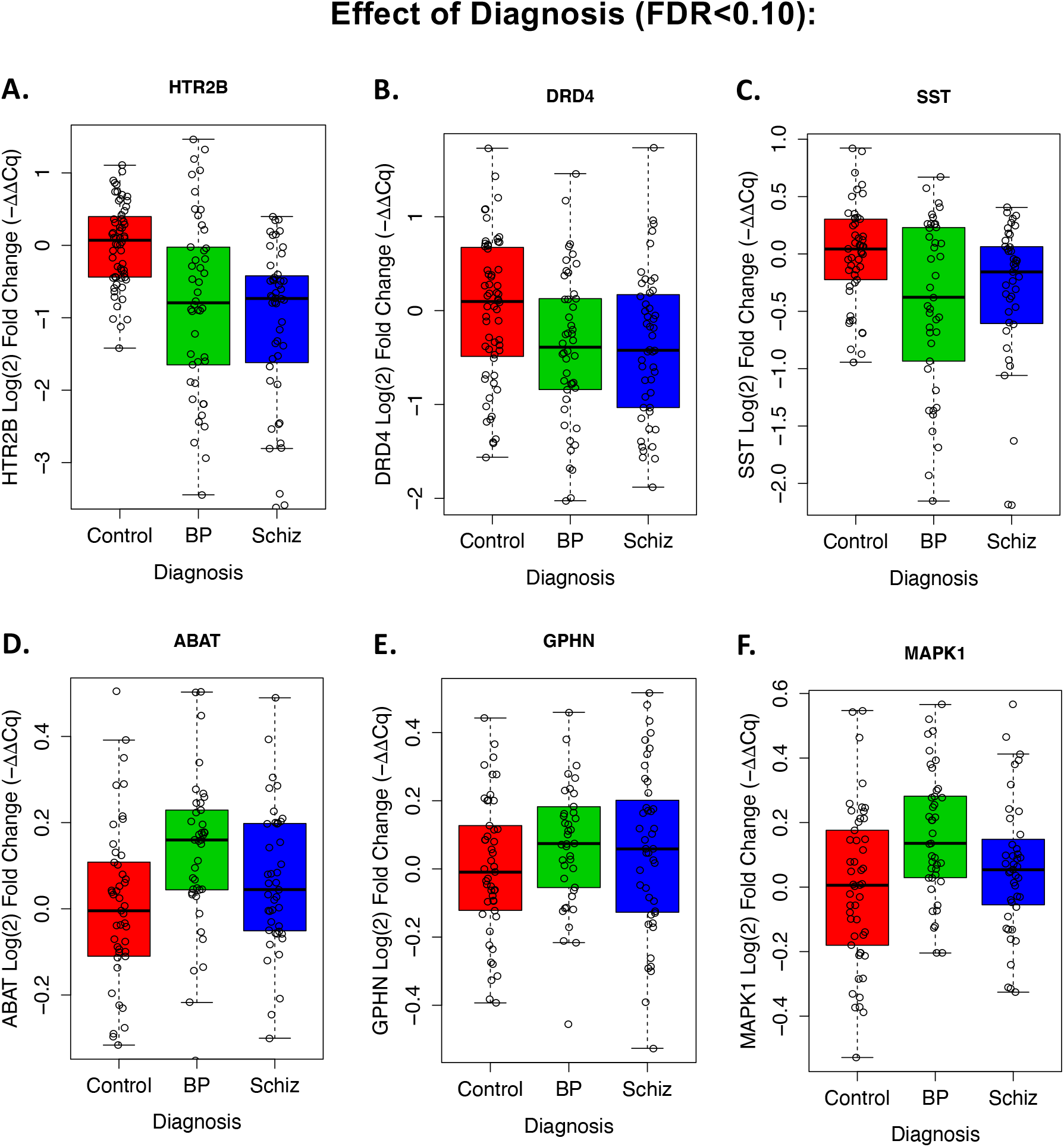
Six target genes showed a relationship with diagnosis that survived false discovery rate correction (FDR<0.10) in our qPCR experiment. Boxplots illustrate the distribution of -ΔΔCq values (log2 fold change (Log2FC), with the average for the CTRL group set as 0) for each sample within each diagnosis group (boxes=first quartile, median, and third quartile, whiskers=range and/or 1.5x the interquartile range if there are outlying data points). Note that each datapoint represents an individual replicate without the additional correction for confounding biological or technical variation provided by our statistical model. **A)** 5-Hydroxytryptamine Receptor 2B (HTR2B) showed an effect of Diagnosis (p=0.000312, FDR=0.0173), with a decrease in both Bipolar Disorder (BP, Log2FC=-0.681, p=0.0270) and Schizophrenia (SCHIZ, Log2FC=-0.980, p=0.000903). **B)** Dopamine Receptor D4 (DRD4) showed an effect of Diagnosis (p=0.00196, FDR=0.0495), with a decrease in both BP (Log2FC=-0.466, p=0.00510) and SCHIZ (Log2FC=-0.347, p=0.0244). **C)** Somatostatin (SST) showed an effect of Diagnosis (p=0.00223, FDR=0.0495), with a trend towards a decrease in BP (Log2FC=-0.308, p=0.0935) and SCHIZ (Log2FC=-0.467, p=0.00689), **D)** 4-aminobutyrate aminotransferase (ABAT) showed an effect of Diagnosis (p=0.00166, FDR=0.0495), with elevation in BP (Log2FC=0.157, p=0.00550) and a trend towards elevation in SCHIZ (Log2FC=0.083, p=0.0993). **E)** Gephyrin (GPHN) showed an effect of Diagnosis (p=0.000283, FDR=0.0173), with elevation in BP (Log2FC=0.156, p=0.00256) and SCHIZ (Log2FC=0.101, p=0.0295), **F)** Mitogen-Activated Protein Kinase 1 (MAPK1) showed an effect of Diagnosis (p=0.00372, FDR=0.0688), with elevation in BP (Log2FC=0.135, p=0.00113) and a trend towards elevation in SCHIZ (Log2FC=0.0869, p=0.0730). Complete statistical reporting for the full concatenated qPCR results can be found in **Table S5**.

Similar to previous studies, we found that somatostatin (SST) showed a relationship with diagnosis, with lower expression in BP and SCHIZ (**Fig 1C)**. Three other genes were related to diagnosis due to elevated expression in both diagnosis groups: 4-aminobutyrate aminotransferase (ABAT), Gephyrin (GPHN), and Mitogen-Activated Protein Kinase 1 (MAPK1) (**Fig 1D-F)**.

### Similarity to Diagnosis Effects in Previous BA10 Microarray Studies

We compared our qPCR results to the published results from previous BA10 microarray studies of BP and SCHIZ^20–24^ (**Table S7**). To increase consistency, we re-annotated and re-analyzed the data from the two studies with publicly-released data^20,21^, and performed a meta-analysis of the SCHIZ effects (Log2FC) in the re-analyzed data.

In general, despite the noise in the microarray data and the lack of sensitivity for low-level expression (**Fig S13F-G**, *e.g*., HTR2B, DRD4), we observed a similar direction of effect in the BA10 microarray datasets for top differentially expressed genes from our qPCR study (SST, ABAT, MAPK1, GPHN). We also observed a weak positive correlation between our full qPCR results and the re-analyzed BA10 microarray results (**Fig 2A, Fig S14**, *qPCR vs. Iwamoto: SCHIZ:* R=0.24, p=0.0179; *BP:* R=0.15, p=0.151: *qPCR vs. Maycox: SCHIZ:* R=0.09, p=0.338*\*not sig*; *qPCR vs. Meta-analysis: SCHIZ:* R=0.24, p=0.0175). Amongst these results, we identified three additional genes that had nominally significant differential expression (p<0.05) in both the qPCR study and BA10 microarray data as well as a consistent direction of effect (GRM5, NSF, SNCA, **Fig 3**). Our meta-analysis also identified six differentially expressed genes (FDR<0.10) that were not included as qPCR targets: a downregulation of TUBB7P, TIE1, HSD17B8, and URM1 and upregulation of CACYBP and GIT2 with SCHIZ (**Fig S16, Table S8**).

**Figure 2.**
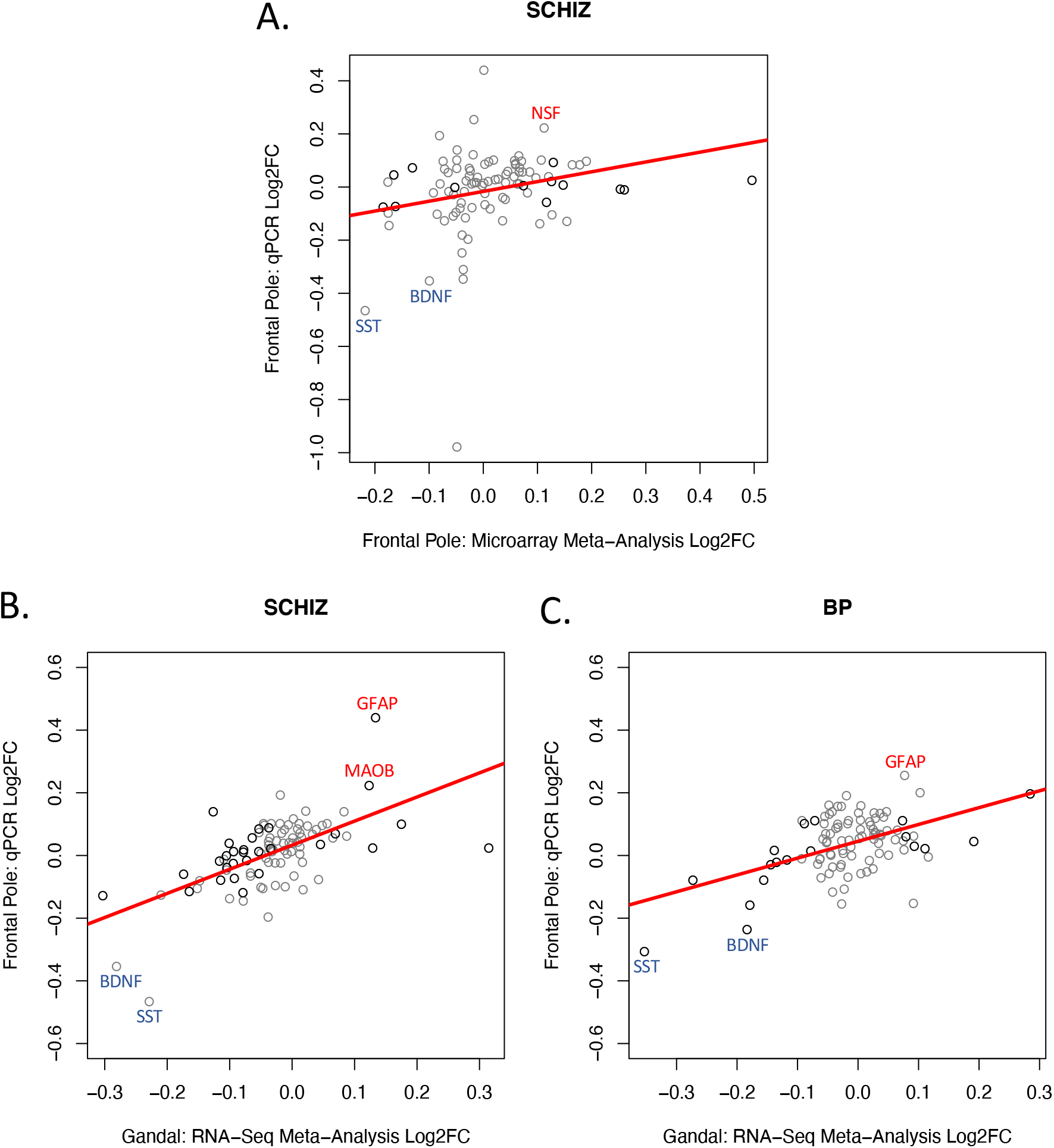
Our qPCR experiment replicated many diagnosis effects observed in the frontal cortex by meta-analyses of differential expression results from less sensitive transcriptional profiling methods (microarray, RNA-Seq). In addition to detecting novel diagnosis effects in low-level expressed genes (HTR2B, DRD4), our qPCR experiment was able to detect diagnosis effects that were observed in the frontal cortex within our current meta-analysis of previous BA10 microarray studies and in a previously-published meta-analysis of RNA-Seq studies of neighboring frontal cortex. **A)** To provide a more robust comparison, we re-annotated and re-analyzed two publicly-available microarray datasets from BA10 ^12,13^ and then performed a meta-analysis to identify the differential expression associated with SCHIZ. A scatterplot is shown illustrating the positive correlation (R=0.243, p=0.0175) between the differential expression (Log2FC) associated with SCHIZ in our BA10 microarray meta-analysis and the differential expression associated with SCHIZ in our qPCR experiment (Log2FC) for all genes that were present in both datasets (n=95). The color of the datapoints signifies whether a gene showed a nominally significant effect of diagnosis in the microarray meta-analysis results (black: p<0.05, grey: p>0.05). **B)** A scatterplot illustrating the positive correlation (R=0.607, p=6.604e-11) between the differential expression associated with SCHIZ (Log2FC) identified in a very large (n=384: 203 CTRL, 181 SCHIZ) meta-analysis of RNA-Seq data from frontal cortical tissue ^23^ and the differential expression associated with SCHIZ in our qPCR experiment (Log2FC) for all genes that were present in both datasets (95 genes). The color of the datapoints signifies whether a gene showed a nominally significant effect of diagnosis in the RNA-Seq meta-analysis results (black: p<0.05, grey: p>0.05). **C)** A scatterplot illustrating the positive correlation (R=0.490, p=4.61e-07) between the differential expression associated with BP (Log2FC) identified in a very large (n=171: 101 CTRL, 70 BP) meta-analysis of RNA-Seq data from frontal cortical tissue ^23^ and the differential expression associated with BP in our qPCR experiment (Log2FC) for all genes that were present in both datasets (95 genes). The color of the datapoints signifies whether a gene showed a nominally significant effect of diagnosis in the RNA-Seq meta-analysis results (black: p<0.05, grey: p>0.05). Datapoints are labeled with official gene symbols for genes that show particularly large Log2FC in both datasets. Full statistical reporting for the correlations between the differential expression observed in different datasets can be found in **Table S9**.

**Figure 3.**
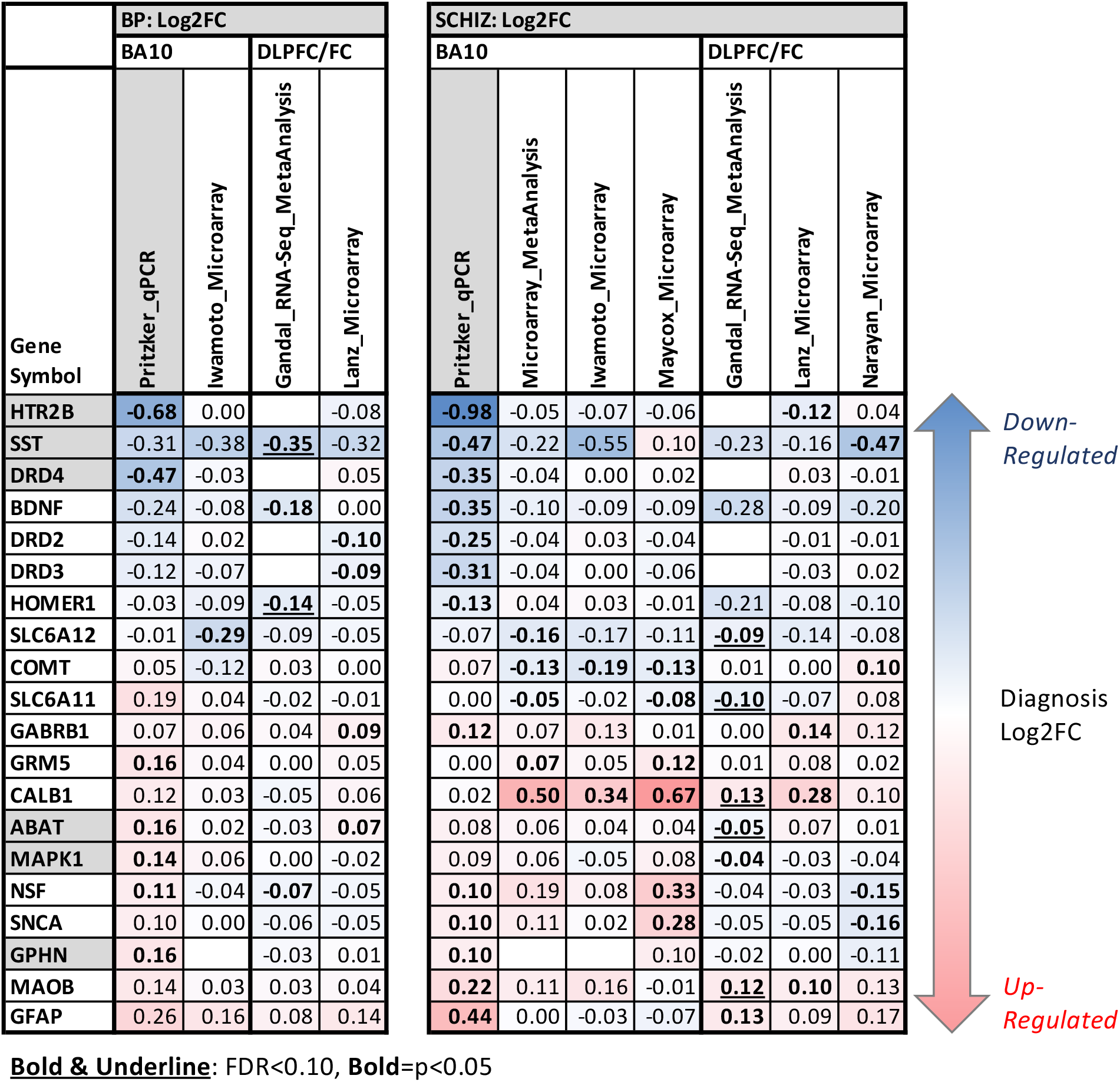
A table overviewing the most supported effects of diagnosis on neurotransmission-related gene expression in BA10. The table illustrates the effects of diagnosis (Log2FC, left: BP, right, SCHIZ) on neurotransmission-related gene expression as identified in our BA10 qPCR experiment (column highlighted in grey), our re-analysis of two BA10 microarray datasets (Iwamoto et al. and Maycox et al. ^12,13^), and our meta-analysis of the SCHIZ results from the two BA10 microarray datasets. These results often paralleled results from neighboring frontal cortex: a large meta-analysis of RNA-Seq data from the DLPFC/frontal cortex (Gandal et al. ^23^) and results from two smaller microarray datasets from the DLPFC with grey matter-focused dissections (Lanz et al. and Narayan et al. ^24,25^ as re-analyzed by ^26^). Note that we did not include the results from the large Gandal et al. ^23^ meta-analysis of microarray data because it was from a more broadly defined anatomical region (“cortex”) and included data from the Maycox et al. ^13^, Lanz et al. ^25^ and Narayan et al. ^24^ studies already in the table. To be included in the table a gene needed to show strong evidence of differential expression in BA10: either FDR<0.10 in our qPCR experiment (gene symbol with grey shading) or p<0.05 (Log2FC in bold text) and consistent direction of effect in two independent datasets from BA10 or in a BA10 dataset and DLPFC/frontal cortex dataset. Throughout the table, red is used to indicate upregulation and blue is used to indicate downregulation. Complete statistical reporting for the full results from our qPCR experiments, microarray re-analysis, and microarray meta-analysis can be found in **Tables S5 & S8**.

### Similarity to Diagnosis Effects in Neighboring Cortical Regions

We observed a strong positive correlation between the diagnosis effects identified in our qPCR study and those previously observed in the dorsolateral prefrontal cortex (DLPFC) and neighboring frontal cortex in a large meta-analysis of RNA-Seq studies (**Fig 2B-C**, n=Log2FC for 95 genes^32^: SCHIZ: R=0.61, p=6.604e-11, BP: R=0.49, p=4.61e-07). As RNA-Seq still lacked sensitivity for measuring low level expression (**Fig S13D-E**), the strength of this correlation likely reflected the statistical power of the RNA-Seq meta-analysis (*SCHIZ vs. CTRL: n=*384; *BP vs. CTRL:* n=171). A comparison of our qPCR findings to smaller DLPFC microarray studies (^33,34^, re-analyzed in^35^), produced weaker positive correlations (all p>0.10: **Fig 4B, Table S9**).

**Figure 4.**
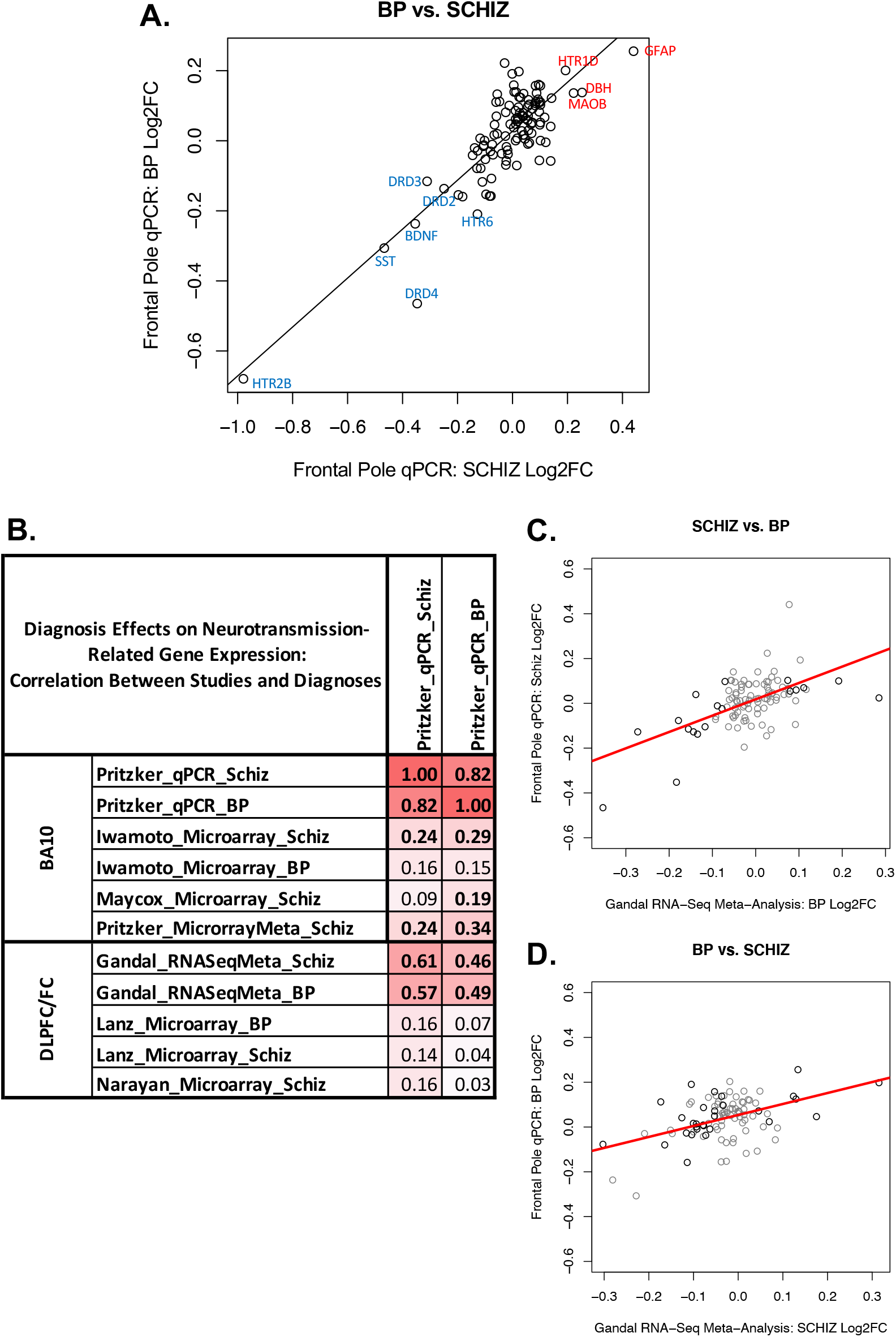
Cross-Diagnosis Comparison: BP and SCHIZ have similar differential expression in BA10 and neighboring cortex. **A**. A scatterplot illustrating the correlation between the differential expression (Log2FC) for BP vs. SCHIZ for the 111 target genes included in our two qPCR datasets. In general, there is a strong positive correlation between the effects of the two diagnoses (R=0.82, p<2e-16). Data points are labeled with official gene symbols for individual genes with particularly large Log2FC for both diagnoses. **B**. A table of correlation coefficients illustrating the similarity between the differential expression (Log2FC) associated with BP and SCHIZ in BA10 in our qPCR study and the differential expression (Log2FC) associated with both BP and SCHIZ in BA10 as measured by microarray (re-analyzed by our laboratory: ^12,13^) and in the DLPFC/frontal cortex as indicated by a meta-analysis of RNA-Seq studies ^23^, or by microarray ^24,25^. Bold text indicates significance (p<0.05). **C-D**. Scatterplots illustrating the cross-diagnosis correlation between the effects of BP and SCHIZ within our qPCR dataset (Log2FC) and the effects of BP and SCHIZ within the results from a large RNA-Seq meta-analysis from neighboring cortex (Log2FC, ^23^) for all genes that were present in both datasets (95 genes). The color of the datapoints signifies whether a gene showed a nominally significant effect of diagnosis in the RNA-Seq meta-analysis results (black: p<0.05, grey: p>0.05). These findings indicate that the similarity between the effects of BP and SCHIZ within our qPCR dataset is not just an artifact due to using the same CTRL group as reference in our analysis, but a valid property of the diagnosis-related differential expression. Full statistical reporting for the correlations between the differential expression observed in association with different diagnoses and different datasets can be found in **Table S9**.

Altogether, there were 13 neurotransmission-related genes that showed at least nominally-significant (p<0.05) diagnosis-related effects in BA10 within either our qPCR experiment or in the re-analyzed BA10 microarray data, as well as similar effects (p<0.05, consistent direction) within the neighboring frontal cortex (**Fig 3**).

### Similarity of Neurotransmission-Related Differential Expression Across Diagnosis Categories

BP and SCHIZ showed strikingly similar effects on gene expression when examining results across all 111 target genes in our qPCR analysis (**Fig 4A**, BP vs. SCHIZ: R=0.82, *p*<2e-16). We also observed a strong correlation between the effects of BP and SCHIZ on neurotransmission-related gene expression within the re-analyzed BA10 microarray data (*Iwamoto:* R=0.43, p=1.32e-05) and within results from previous frontal cortical studies (RNA-Seq meta-analysis^32^: R=0.80, p<2e-16; Microarray^34^: R=0.82, p<2.2e-16, **Table S9**).

Alone, the strong correlation between the effects of different diagnoses within individual datasets is difficult to interpret, because diagnosis groups are typically compared to a shared control group. However, when comparing across datasets, these correlations become more compelling (**Fig4B-D**). For example, the correlation between the effects of SCHIZ within our qPCR dataset and the effects of BP within the RNA-Seq meta-analysis^32^ was almost as large as comparing the effects of SCHIZ within our qPCR dataset to the effects of SCHIZ within the RNA-Seq meta-analysis (*SCHIZ vs. BP:* R=0.57, p=2.39e-09). A similarly strong correlation was observed when comparing the effects of BP in our qPCR study to the effects of SCHIZ in the RNA-Seq meta-analysis (*BP vs. SCHIZ:* R=0.46, p=3.39e-06).

These results suggest that the differential expression associated with BP and SCHIZ in BA10 may reflect symptoms, risk factors, or experiences common to both disorders.

### Exploratory Analyses: Factors Contributing to Diagnosis-Related Gene Expression

To further explore the function of our top diagnosis-related genes (n=20, **Fig 3**), we examined the relationship between gene expression in our qPCR dataset and a rich database of 49 clinical characteristics compiled via in-depth psychological autopsy (**Fig S2**). We evaluated the effect of these clinical variables on gene expression while controlling for diagnosis and relevant biological and technical covariates. Exploratory results with independent validation or support are highlighted below (*details in Supplement)*.

#### Overlap Between the Effects of Diagnosis and Common Therapeutics

One question that typically arises while interpreting diagnosis effects within human post-mortem studies is whether the observed effects are due to the illness itself or its treatment. Indeed, there were a large number of documented effects (FDR<0.05) of therapeutics on the expression of our top 20 diagnosis-related genes within the comprehensive Drug Gene Budger database (https://maayanlab.cloud/DGB/^37^), including 169 antipsychotic effects and 28 antidepressant effects from a variety of experiments. Given this precedent, we explored the effect of exposure to therapeutics in our qPCR dataset while controlling for diagnosis. Although our analyses were underpowered (*antipsychotics:* n=9; *antidepressants:* n=8), we observed some suggestive results that were bolstered by previous evidence (**Fig S17A**). MAOB was decreased with antidepressants (FDR<0.10), paralleling documented functional inhibition^43,44^. We also saw a nominal (p<0.05) decrease in GPHN with antidepressants, and increase in COMT and BDNF with antipsychotics that mirrored the effects in Drug Gene Budger^37^. Notably, the effects of antipsychotics tended to be in the opposite direction of diagnosis effects in BA10 (**Fig S17B-D**). In contrast, the diagnosis-related increase in SNCA could be an artifact of therapeutics, as we found a nominal (p<0.05) increase with antidepressants, and substantial previous documentation of upregulation with both antidepressants and antipsychotics (19 effects: Drug Gene Budger^36^).

#### Overlap Between the Effects of Diagnosis and Substances of Abuse

Similar to many psychiatric studies, our diagnosis groups contained an elevated rate of documented dependence on alcohol and tobacco as well as use of other substances of abuse around the time of death (cannabinoids, opioids, stimulants, **Fig S18A-B**), whether due to therapeutic use, recreation or a substance use disorder. This elevated rate of substance use could partially reflect better documentation, as toxicology analysis was performed more often for psychiatric subjects (CTRL: 12%, SCHIZ: 43%, BP: 36%) due to manner of death, but the high rate of overdose (SCHIZ: 23%, BP: 52%) suggested objectively otherwise.

Therefore, we explored whether diagnosis effects in our qPCR dataset could be better explained by substance use. For our top 20 diagnosis-related genes we examined the effect of recent exposure to substances of abuse while controlling for diagnosis. We found that the effects (Log2FC) were surprisingly consistent across substance categories (median: R=0.77, range: R=-0.06-0.88, **Fig S18D**), in a manner that exceeded the correlated use of those substances (median: R=0.44, range: R=0.068-0.52, **Fig S18C**), suggesting that the results might broadly reflect use behaviors. This was further supported by the similarity between our results and previously documented effects of Opioid Use Disorder (RNA-Seq^39^: R=0.26-0.79, median R=0.58, **Fig 5A, Fig S18D**) and Alcohol Abuse Disorder in the cortex (microarray meta-analysis^32^ & RNA-Seq^38^: R=-0.03-0.69, median R=0.36, **Fig S18D**), but we did not see a similar parallel with the effects of substance use in the smaller Iwamoto BA10 microarray dataset (**Fig S18D**, smoking, heavy alcohol, or heavy drug use).

**Figure 5.**
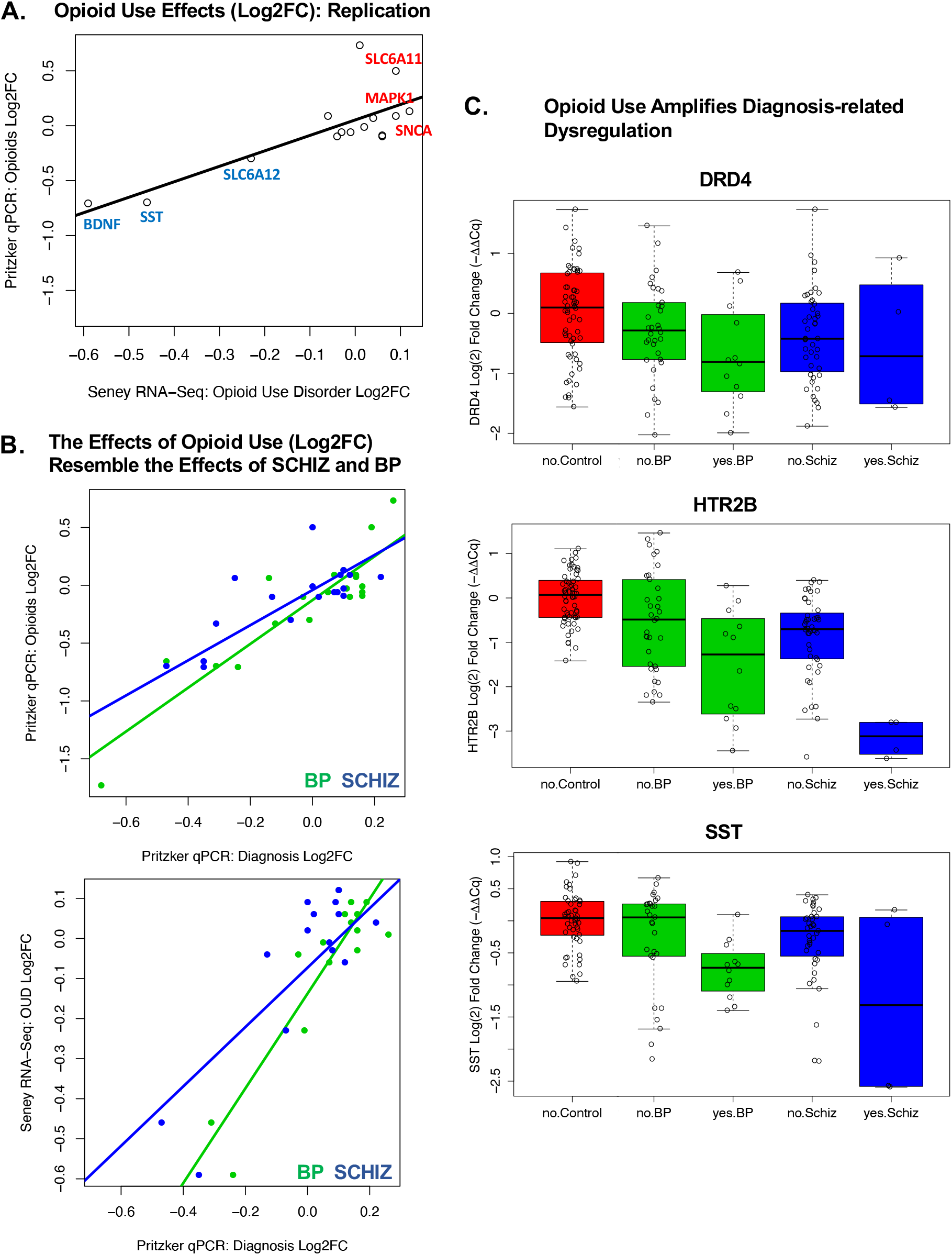
Exploratory: Substances of abuse may be associated with similar dysregulation of neurotransmission-related gene expression in BA10 as SCHIZ and BP. Within a set of exploratory analyses, we estimated the differential expression (Log2FC) in the frontal pole (BA10) in our qPCR dataset associated with a variety of substances of abuse (tobacco, cannabinoids, stimulants, opioids) while controlling for diagnosis. Substance exposure was defined by indication of usage within the subjects’ clinical records, family interviews, toxicology reports, and coroners reports. To reduce false discovery due to multiple comparisons, this exploratory analysis was limited to the 20 genes with the most reliable diagnosis effects in BA10 (listed in **Fig 3**). The pattern revealed by our exploratory analysis was surprisingly consistent across substances (**Fig S18D**), but the largest effects were observed with opioid exposure (n=8 subjects). **A**. Replication: Opioid exposure in our BA10 qPCR dataset was associated with similar differential expression (Log2FC) to what had been observed previously in opioid use disorder in the DLPFC using RNA-Seq ^30^ (n=15 diagnosis-related genes, Pritzker BA10 qPCR vs. Seney DLPFC RNA-Seq: R=0.79, p=0.000499). **B**. Relationship with diagnosis: In general, the effects of diagnosis on gene expression (Log2FC) in BA10 measured in our qPCR study correlated positively with the effects of a variety of substances of abuse (Log2FC; tobacco, cannabinoids, stimulants, opioids) as estimated while controlling for diagnosis in our dataset. The top scatterplot uses opioid exposure as an example to illustrate the similarity between the effects associated with substance use and diagnosis (n=20 diagnosis-related genes, x-axis: Blue=Bipolar Disorder Log2FC, Green=Schizophrenia Log2FC, Opioid Log2FC vs.: BP: R=0.90, p=5.04e-08, SCHIZ: R=0.91, p=2.21e-08). The bottom scatterplot shows that the similarity between the effects of opioid use and diagnosis replicates when using the effects of opioid use disorder measured in the previously published Seney et al. ^30^ dataset (n=15 diagnosis-related genes, Opioid Use Disorder Log2FC vs.: BP: R=0.89, p=6.43e-06, SCHIZ: R=0.76, p=0.000678). **C**. Opioid use appears to amplify diagnosis-related dysregulation: Example boxplots illustrate the effect of opioid exposure within our diagnosis groups for top diagnosis-related genes (DRD4: effect of Diagnosis (p=0.0190), effect of Opioids (p=0.000208); HTR2B: effect of Diagnosis (p=9.94e-05), effect of Opioids (p=9.42e-09); SST: effect of Diagnosis (p=0.0111), effect of Opioids (p=0.000146)). Plotting conventions follow **Fig 1**, with “yes” and “no” indicating exposure to opioids. Differential expression results for other genes and substances of abuse can be found in **Fig S19**. Full statistical reporting for correlations can be found in **Table S9**.

For several genes, the effects of substance use were particularly large (FDR<0.10) and consistent with evidence from larger studies of substance use (Alcohol Abuse Disorder^32,38^ or Opioid Use Disorder^39^) in neighboring frontal cortex (**Fig S19)**. GFAP was increased with a variety of substances (tobacco, simulants, opioids, overdose). HTR2B, SST, and BDNF were decreased with opioid use (**Fig 5C)**, overdose (HTR2B, SST), and nominally other substances. MAPK1 and GABRB1 were increased with stimulant use and nominally other substances. We also observed decreased DRD4 and SLC6A12 and increased SLC6A11 with substance use (FDR<0.10, **Fig 5C, Fig S19**) but lacked independent validation.

Notably, the effects of substance use broadly resembled the effects of diagnosis (**Fig 5B, Table S9**, all relationships R=0.54-0.90, p<0.015), but the diagnosis effects typically persisted after controlling for substance use (**Fig 5C, Fig S20**). These exploratory findings are provocative, suggesting a mechanism by which substance use might converge with diagnosis-related dysregulation in frontal cortical function, but require further independent validation.

#### Overlap Between the Effects of Diagnosis, Symptom Profiles, and Related Behaviors

In general, there were very few symptoms and related behaviors that had significant effects (FDR<0.10) in our dataset that resembled what we had observed for diagnosis. Surprisingly, the few relationships that we did observe that resembled the effects of diagnosis were often broadly related to executive dysfunction: “agitated” (SNCA, MAPK1, SLC6A12, HTR2B), “interactions with the legal system” (GABRB1), “reckless” (NSF), “fatigue” (BDNF), and “trouble concentrating” (GPHN) (**Fig S21A&B, Table S9**). These results are provocative, especially since some of these genes were also differentially expressed in the cortex in other diagnoses (**Fig S21A&C**) or in response to stress hormones (**Fig S21A&D**), but since the exploratory analyses were run while controlling for diagnosis, we may have had limited power to detect relationships with symptoms that more canonically define BP and SCHIZ, such as psychosis and disturbed affect. Further validation is strongly recommended.

## Discussion

Using the highly sensitive method of qPCR, we identified a large downregulation in BP and SCHIZ of two important therapeutic targets with low-levels of expression in BA10, HTR2B and DRD4, and detected weaker downregulation of DRD2 and DRD3. Our qPCR study also highlighted the differential expression of genes related to signaling via GABA and Glutamate, including SST, ABAT, GPHN, and MAPK1. To bolster our findings, we performed a meta-analysis of publicly-released BA10 microarray data^20,21^, and identified sources of convergence with our qPCR results. We then compared our BA10 differential expression results to previous transcriptional profiling experiments in neighboring cortex^32–34^ and found many similar effects. Using these converging sources of evidence, we identified twenty neurotransmission-related genes that were differentially expressed in BP and SCHIZ in BA10.

This differential expression was often similar across diagnoses, suggesting that gene expression associated with BP and SCHIZ in BA10 may reflect symptoms, risk factors, or experiences common across the two disorders. To explore the factors contributing to diagnosis-related differential expression, we leveraged the unusually large database of clinical metadata associated with our qPCR samples to explore the relationship between BA10 gene expression, therapeutics, substances of abuse, and symptom profiles, and validated exploratory findings with publicly-available datasets. We found evidence that therapeutics may produce differential expression that often opposes the effects of diagnosis. We also observed that substances of abuse may be associated with similar effects on gene expression in the frontal cortex as BP and SCHIZ, potentially amplifying diagnosis-related dysregulation.

### Downregulation of serotonin receptor HTR2B in BP and SCHIZ

The largest effect we observed in our study was a surprisingly strong downregulation of the serotonin receptor HTR2B. Subjects with BP and SCHIZ had almost half as much HTR2B expression in BA10 as control subjects. We infer that this expression is probably located in vasculature and microglia based on relative prevalence within previous anatomical and cell culture studies^45–47^. If so, downregulation may impact two key features of SCHIZ and BP observed in the frontal lobe.

First, SCHIZ and BP symptoms correlate with altered patterns of blood flow in BA10^11,14,16^. HTR2B is expressed in endothelial cells of the cerebral vasculature ^47,48^ and regulates blood flow via the release of nitric oxide ^45,49^. Therefore, we hypothesize that HTR2B downregulation could contribute to altered BA10 cerebral blood flow.

Second, during adolescence and early adulthood, the frontal cortex experiences intense pruning that reduces grey matter thickness^6^. This pruning period coincides with the onset of BP and SCHIZ^7^ and may be disrupted in association with psychiatric illness^8^. Since microglia actively engulf synaptic structures during postnatal development, microglial dysfunction could contribute to aberrant pruning^50^. Pure microglial cultures express HTR2B^46^, and in the brain, a lack of HTR2B receptors affects microglial activation and impedes their ability to mediate proper synaptic refinement^51^. These findings suggest that a deficit in HTR2B activity could produce defective grey matter pruning in psychiatric subjects. Conversely, a diagnosis-related decrease in microglia could also produce an overall decrease in HTR2B expression.

In support of the hypothesis that decreased HTR2B could play a role in the development of the disorder, previous exome sequencing studies have identified genetic variants associated with SCHIZ within the HTR2B gene that are predicted to be missense/damaging or stop-gained^52,53^. HTR2B knock-out mice also exhibit an antipsychotic-sensitive behavioral phenotype reminiscent of SCHIZ, including decreased pre-pulse inhibition, dysfunctional social interaction, memory and attentional deficits^54^. Acute exposure to the HTR2B antagonist RS127445 produces similar deficits^54^.

Within our exploratory results, we also observed decreased HTR2B expression associated with substance use, especially opioids, and the symptom of agitation. These findings are intriguing, since stop-gained genetic variants within the HTR2B gene are associated with impulsive behavior, including impulsive aggression, especially under the influence of alcohol^53,55^. HTR2B knock-out mice are also more impulsive and responsive to novelty^53^ and amphetamine^54^, whereas, paradoxically, HTR2B antagonists can decrease responses to many drugs of abuse, including hyperactivity and conditioned place preference^56^. Collectively, these findings suggest that deficits in HTR2B could affect impulsivity and drug-taking behavior, offering one explanation for the common covariation of substance use with BP and SCHIZ.

### Dopaminergic Gene Expression in BP and SCHIZ

The availability of dopamine is pivotal to frontal cortex function^57–59^. Disturbed dopamine function in the frontal cortex is considered an important feature of psychotic disorders, as most antipsychotic treatments target DRD2-like dopamine receptors (DRD2, DRD3, DRD4)^19^. These dopamine receptors are located near genetic risk loci for SCHIZ (genome wide association (DRD2)^60^; linkage and association studies (DRD2, DRD3, DRD4)^61–64^), and contain risk variants predicted to be missense/damaging or stop-gained (DRD2^65^, DRD3^52^), implying that deficits in dopamine function may contribute to the disorder.

However, the relationship between psychotic disorders and the expression of DRD2-like dopamine receptors has been previously difficult to characterize due to their low levels of expression in the frontal cortex^19,66^. Early attempts using PCR produced mixed results, including in BA10^67–69^, although notably *in situ* hybridization detected region-specific, large-magnitude decreases in DRD4 and DRD3 in SCHIZ in nearby BA11^70^. Using more modern qPCR methods, DRD2 was found to be decreased in SCHIZ and increased in BP in the neighboring DLPFC^19^. We show that current qPCR methodology can reliably measure very low levels of expression, including the expression of DRD2-like dopamine receptors, and reveals surprisingly large diagnosis effects in BA10 that dwarf those previously measured with less sensitive methodology (microarray, RNA-Seq).

Of the dopamine receptors, DRD4 showed the largest decrease in BP and SCHIZ. Decreased DRD4 receptor activity could contribute to diagnosis-related disruptions in excitatory/inhibitory balance. An altered excitatory/inhibitory balance in areas associated with cortical inhibitory control is hypothesized to be a key feature of psychiatric illnesses with severe social and cognitive deficits^71,72^. DRD4 agonists and antagonists decrease and increase postsynaptic GABA(A) receptor currents, respectively^73^, and decreased DRD4 responsiveness is associated with decreased cortical hemodynamic activity^74^. In mice, DRD4 knockout causes disrupted GABAergic activity, which facilitates the development of SCHIZ-like symptoms under stress^75^. Moreover, the atypical antipsychotic clozapine has a high affinity for DRD4, and DRD4 knock-out mice are less responsive to its effects^76^, suggesting that decreased DRD4 in BP and SCHIZ might not only contribute to disease etiology but also hinder successful treatment.

During our exploratory analyses, we also observed decreased DRD4 in association with substance use but were unable to find validation in an independent dataset. That said, DRD4 knock-out mice show an enhanced response to ethanol, cocaine, and methamphetamine^76^, and DRD4 genetic polymorphisms have been implicated in substance use and attention deficit hyperactivity disorder^77^. Therefore, similar to HTR2B, decreased DRD4 might promote the common covariation of substance use with BP and SCHIZ.

The enzyme catechol-O-methyltransferase (COMT) has been theorized to play a role in psychiatric disorders^58,78–80^. Termination of monoamine activity in the frontal cortex depends mostly on the activity of COMT instead of the dopamine transporter^81^. COMT is located within a risk locus for SCHIZ implicated by copy number variation^82^ and by meta-analyses of linkage^63,64^ and association studies^61,62^. We observed a nominal decrease in COMT in both of the BA10 microarray datasets that we reanalyzed. This downregulation might increase dopamine levels^58^, with behavioral consequences such as aggression and increased sensitivity to stress^81^. There was not a similar diagnosis effect in our qPCR dataset, but COMT was increased with antipsychotic medication in a manner paralleling previous observations from cell culture and other tissues^37^.

### GABA-related Gene Expression in BP and SCHIZ

Similar to our current work, previous studies in neighboring frontal areas have repeatedly shown large decreases in somatostatin (SST) in BP and SCHIZ (^32,35,83–85^, review:^86^). SST is selectively expressed in GABA-ergic interneurons which represent ∼30% of the total cortical interneuron population^87^. SST neurons synapse on pyramidal cell dendritic shafts and spines, indicating a role in filtering excitatory glutamatergic inputs^88^. Reduced SST interneuron function in SCHIZ is theorized to disrupt cortical information integration processes, such as working memory^86^. That said, decreased SST is not specific to psychotic disorders, and has been observed in Major Depressive Disorder (**Fig S21**) and following chronic stress^89^. SST neurons in the frontal cortex modulate anxiety-like behavior, and SST knock-out mice display elevated anxiety and corticosterone levels^89^. Both post-mortem and preclinical studies link SST to drug-taking behavior^39,89^, which is further supported by the decreased SST with substance use observed in our exploratory analyses. Altogether, SST appears to be a compelling convergence point for stress to aggravate a variety of psychiatric symptoms.

Both BP and SCHIZ also exhibited increased Gephryin (GPHN), a postsynaptic scaffolding protein which is abundant in the human cortex and localized to cell bodies and the apical dendrites of pyramidal neurons^90^. GPHN helps cluster, anchor, and stabilize GABA(A) and glycine receptors at inhibitory synapses^91^. Therefore, the upregulation of GPHN in BP and SCHIZ could be related to the shift in GABA(A) receptor subunit composition that occurs at postsynaptic sites^92,93^, potentially contributing to increased GABA(A) receptor binding in the frontal cortex in SCHIZ^93,94^. This change in GABA(A) receptor binding, as well as the increased GABA(A) Receptor Subunit Beta1 (GABRB1) expression that we observed in SCHIZ, might serve as a compensatory mechanism for disturbed local GABAergic neurotransmission.

### Astrocyte-related Gene Expression in BP and SCHIZ

We observed a large upregulation of the intermediate filament Glial fibrillary acidic protein (GFAP) in SCHIZ, an effect previously observed in neighboring frontal cortex^32^. Increased expression of GFAP is a hallmark of astrocyte activation^95,96^ and follows increased intracellular calcium signaling ^97^. Astrocyte reactivity is found in many pathological states involving excitotoxicity^95,96^, but we found little evidence of diagnosis-related glutamatergic alterations in BA10, with the exception of a nominal elevation of Glutamate metabotropic receptor 5 (GRM5). Instead, we hypothesize that astrocyte activation elicited by GABAergic stimulation might be a feature of BP and SCHIZ.

In general, GABA is considered an inhibitory neurotransmitter, but it can elicit intracellular calcium increases and regional excitation in the glial syncytium^98,99^. This increase in astrocytic calcium can be elicited by GABA(A) or GABA(B) receptor activation^100,101^. Therefore, our results showing increased GABRB1 receptor subunit expression in SCHIZ could suggest a mechanism for increased astrocyte activation. We also found increased Monoamine oxidase B (MAOB) in both disorders. This enzyme synthesizes GABA in astrocytes^102^. GFAP-expressing reactive astrocytes use MAOB to produce and release excess GABA, causing tonic inhibition and altered dopamine function^103^. Therefore, the presence of more reactive astrocytes (GFAP) and MAOB in BP and SCHIZ could not only be caused by GABA signaling, but contribute to further alterations in regional excitatory/inhibitory balance and dopaminergic dysfunction.

There was decreased expression of the GABA transporter GAT-3 (gene: SLC6A11) in SCHIZ. SLC6A11 is abundantly expressed in astrocyte processes surrounding synapses and neuronal bodies^104,105^. GAT-3 activity causes extracellular GABA uptake and sodium ion accumulation in astrocytes. Due to sodium/calcium exchange, this leads to increased intracellular astrocytic calcium^104^. Therefore, decreased GAT-3 function in SCHIZ could contribute to excess GABA, but dampen further elevations in calcium due to GABA signaling.

Finally, astrocytic metabolism of GABA is performed by mitochondrial GABA transaminase (gene: ABAT)^106^. ABAT was upregulated in BP and SCHIZ. Increased expression of this degrading enzyme could shorten the availability of GABA, causing disinhibition. Increased ABAT expression could underlie manic symptoms in BP, as ABAT deficiency has been associated with inconsolable crying, dullness, and lethargy in case studies^107^ and increased ABAT has been associated with aggression in rodents^108^.

Interestingly, signatures of glial activation (upregulation of GFAP) and GABA dysfunction (upregulation of SLC6A11 and GABRB1) were also strongly associated with substance use in our exploratory analyses, as well as “interactions with the legal system” (GABRB1) and “fatigue” (SLC6A11). This raises the question as to whether glial activation and GABA dysfunction in BA10 might be particularly important for the executive dysfunction and cognitive symptoms accompanying BP and SCHIZ. This could complement recent evidence showing elevated GFAP in serum accompanying declines in executive function, memory and attention in older individuals^109^.

#### Limitations and Future Directions

Differential expression is not always predictive of protein levels, but proteins are challenging to quantify in post-mortem tissue due to degradation. Moreover, our qPCR results were derived from gross anatomical samples and therefore specific cellular relationships cannot be determined. Future endeavors will evaluate mRNA expression in BP and SCHIZ within BA10 cryosections for high resolution cortical layer quantitation.

#### Conclusion

SCHIZ and BP are associated with a notable dysregulation of neurotransmission-related gene expression in BA10, a cortical area highly specialized in humans. These effects included alterations in gene expression underlying serotonin, dopamine, GABA, and astrocytic networks. We hypothesize that these disruptions underly specific cognitive and behavioral manifestations common to the two disorders.

## Supporting information

Supplemental Methods and Results

## Data Availability

All data produced will be available online on FigShare following publication.

## Acknowledgements

We thank Jennifer Fitzpatrick and Linda Morgan for their expert technical assistance on the project, and Richard Stein, Ph.D. for his work overseeing the clinical metadata database.

This work was primarily supported by the Pritzker Neuropsychiatric Disorders Research Consortium [to HA and SJW]. This work was further supported by grants from the Hope for Depression Research Foundation (RGA No. DTF Phase II [D]), National Institute on Drug Abuse (Grant #U01-DA043098 [to HA]), Office of Naval Research (Grant # 00014-19-1-2149 [to HA]), and National Institute of Mental Health (Grant # R01-MH-08580 [to MPV]). Research training for LTF was supported by the Michigan Research and Discovery Scholars (MRADS) program, and ER was supported by the Undergraduate Research Opportunity Program (UROP) at the University of Michigan.

## References

1. Ramnani, N. & Owen, A. M. Anterior prefrontal cortex: insights into function from anatomy and neuroimaging. Nat Rev Neurosci 5, 184–194 (2004).

2. Semendeferi, K., Armstrong, E., Schleicher, A., Zilles, K. & Van Hoesen, G. W. Prefrontal cortex in humans and apes: A comparative study of area 10. Am. J. Phys. Anthropol. 114, 224–241 (2001).

3. Semendeferi, K. et al. Spatial organization of neurons in the frontal pole sets humans apart from great apes. Cereb Cortex 21, 1485–1497 (2011).

4. Petrides, M. & Pandya, D. N. Dorsolateral prefrontal cortex: comparative cytoarchitectonic analysis in the human and the macaque brain and corticocortical connection patterns: Dorsolateral prefrontal cortex in human and monkey. European Journal of Neuroscience 11, 1011–1036 (1999).

5. Koch, S. B. J., Mars, R. B., Toni, I. & Roelofs, K. Emotional control, reappraised. Neurosci Biobehav Rev 95, 528–534 (2018).

6. Sydnor, V. J. et al. Neurodevelopment of the association cortices: Patterns, mechanisms, and implications for psychopathology. Neuron 109, 2820–2846 (2021).

7. Bethlehem, R. a. I. et al. Brain charts for the human lifespan. Nature 604, 525–533 (2022).

8. Hibar, D. P. et al. Cortical abnormalities in bipolar disorder: an MRI analysis of 6503 individuals from the ENIGMA Bipolar Disorder Working Group. Mol Psychiatry 23, 932–942 (2018).

9. van Erp, T. G. M. et al. Cortical Brain Abnormalities in 4474 Individuals With Schizophrenia and 5098 Control Subjects via the Enhancing Neuro Imaging Genetics Through Meta Analysis (ENIGMA) Consortium. Biol Psychiatry 84, 644–654 (2018).

10. Ongür, D. et al. Default mode network abnormalities in bipolar disorder and schizophrenia. Psychiatry Res 183, 59–68 (2010).

11. Glahn, D. C. et al. Beyond hypofrontality: a quantitative meta-analysis of functional neuroimaging studies of working memory in schizophrenia. Hum Brain Mapp 25, 60–69 (2005).

12. Tu, P.-C. et al. Identification of common neural substrates with connectomic abnormalities in four major psychiatric disorders: A connectome-wide association study. Eur Psychiatry 64, e8 (2020).

13. Zhou, Y. et al. The selective impairment of resting-state functional connectivity of the lateral subregion of the frontal pole in schizophrenia. PLoS One 10, e0119176 (2015).

14. Alústiza, I., Radua, J., Pla, M., Martin, R. & Ortuño, F. Meta-analysis of functional magnetic resonance imaging studies of timing and cognitive control in schizophrenia and bipolar disorder: Evidence of a primary time deficit. Schizophr Res 188, 21–32 (2017).

15. Jung, H.-Y. et al. White matter correlates of impulsivity in frontal lobe and their associations with treatment response in first-episode schizophrenia. Neurosci Lett 767, 136309 (2022).

16. Hoptman, M. J., Antonius, D., Mauro, C. J., Parker, E. M. & Javitt, D. C. Cortical thinning, functional connectivity, and mood-related impulsivity in schizophrenia: relationship to aggressive attitudes and behavior. Am J Psychiatry 171, 939–948 (2014).

17. Pietraszek, M. Significance of dysfunctional glutamatergic transmission for the development of psychotic symptoms. Pol J Pharmacol 55, 133–154 (2003).

18. Selvaraj, S., Arnone, D., Cappai, A. & Howes, O. Alterations in the serotonin system in schizophrenia: a systematic review and meta-analysis of postmortem and molecular imaging studies. Neurosci Biobehav Rev 45, 233–245 (2014).

19. Zhan, L. et al. Altered expression and coregulation of dopamine signalling genes in schizophrenia and bipolar disorder. Neuropathol Appl Neurobiol 37, 206–219 (2011).

20. Iwamoto, K., Kakiuchi, C., Bundo, M., Ikeda, K. & Kato, T. Molecular characterization of bipolar disorder by comparing gene expression profiles of postmortem brains of major mental disorders. Mol Psychiatry 9, 406–416 (2004).

21. Maycox, P. R. et al. Analysis of gene expression in two large schizophrenia cohorts identifies multiple changes associated with nerve terminal function. Mol Psychiatry 14, 1083–1094 (2009).

22. Ohayon, S., Yitzhaky, A. & Hertzberg, L. Gene expression meta-analysis reveals the up-regulation of CREB1 and CREBBP in Brodmann Area 10 of patients with schizophrenia. Psychiatry Res 292, 113311 (2020).

23. Scarr, E., Udawela, M. & Dean, B. Changed frontal pole gene expression suggest altered interplay between neurotransmitter, developmental, and inflammatory pathways in schizophrenia. NPJ Schizophr 4, 4 (2018).

24. Scarr, E., Udawela, M. & Dean, B. Changed cortical risk gene expression in major depression and shared changes in cortical gene expression between major depression and bipolar disorders. Aust N Z J Psychiatry 53, 1189–1198 (2019).

25. Purves, D. et al. Neuroscience. (Sinauer Associates, 2001).

26. Diagnostic and statistical manual of mental disorders: DSM-5. (American Psychiatric Association, 2013).

27. Atz, M. et al. Methodological considerations for gene expression profiling of human brain. J. Neurosci. Methods 163, 295–309 (2007).

28. Livak, K. J. & Schmittgen, T. D. Analysis of relative gene expression data using real-time quantitative PCR and the 2(-Delta Delta C(T)) Method. Methods 25, 402–408 (2001).

29. Viechtbauer, W. Conducting Meta-Analyses in R with The metafor Package. Journal of Statistical Software 36, (2010).

30. Ritchie, M. E. et al. limma powers differential expression analyses for RNA-sequencing and microarray studies. Nucleic Acids Res. 43, e47 (2015).

31. Pollard, K. S., Dudoit, S. & Laan, M. J. van der. Multiple Testing Procedures: the multtest Package and Applications to Genomics. in Bioinformatics and Computational Biology Solutions Using R and Bioconductor 249–271 (Springer, New York, NY, 2005). doi:10.1007/0-387-29362-0_15.

32. Gandal, M. J. et al. Shared molecular neuropathology across major psychiatric disorders parallels polygenic overlap. Science 359, 693–697 (2018).

33. Narayan, S. et al. Molecular profiles of schizophrenia in the CNS at different stages of illness. Brain Res. 1239, 235–248 (2008).

34. Lanz, T. A. et al. Postmortem transcriptional profiling reveals widespread increase in inflammation in schizophrenia: a comparison of prefrontal cortex, striatum, and hippocampus among matched tetrads of controls with subjects diagnosed with schizophrenia, bipolar or major depressive disorder. Transl Psychiatry 9, 151 (2019).

35. Hagenauer, M. H. et al. Inference of cell type content from human brain transcriptomic datasets illuminates the effects of age, manner of death, dissection, and psychiatric diagnosis. PLoS ONE 13, e0200003 (2018).

36. Martin, M. V., Mirnics, K., Nisenbaum, L. K. & Vawter, M. P. Olanzapine Reversed Brain Gene Expression Changes Induced by Phencyclidine Treatment in Non-Human Primates. Mol Neuropsychiatry 1, 82–93 (2015).

37. Wang, Z. et al. Drug Gene Budger (DGB): an application for ranking drugs to modulate a specific gene based on transcriptomic signatures. Bioinformatics 35, 1247–1248 (2019).

38. Kapoor, M. et al. Analysis of whole genome-transcriptomic organization in brain to identify genes associated with alcoholism. Transl Psychiatry 9, 89 (2019).

39. Seney, M. L. et al. Transcriptional Alterations in Dorsolateral Prefrontal Cortex and Nucleus Accumbens Implicate Neuroinflammation and Synaptic Remodeling in Opioid Use Disorder. Biol Psychiatry 90, 550–562 (2021).

40. Flati, T. et al. A gene expression atlas for different kinds of stress in the mouse brain. Sci Data 7, 437 (2020).

41. Wu, Y., Li, X., Liu, J., Luo, X.-J. & Yao, Y.-G. SZDB2.0: an updated comprehensive resource for schizophrenia research. Hum Genet 139, 1285–1297 (2020).

42. Wu, Y., Yao, Y.-G. & Luo, X.-J. SZDB: A Database for Schizophrenia Genetic Research. Schizophr Bull 43, 459–471 (2017).

43. Fisar, Z., Hroudová, J. & Raboch, J. Inhibition of monoamine oxidase activity by antidepressants and mood stabilizers. Neuro Endocrinol Lett 31, 645–656 (2010).

44. Leonardi, E. T. & Azmitia, E. C. MDMA (ecstasy) inhibition of MAO type A and type B: comparisons with fenfluramine and fluoxetine (Prozac). Neuropsychopharmacology 10, 231–238 (1994).

45. Schmuck, K., Ullmer, C., Kalkman, H. O., Probst, A. & Lubbert, H. Activation of meningeal 5-HT2B receptors: an early step in the generation of migraine headache? Eur J Neurosci 8, 959–967 (1996).

46. Krabbe, G. et al. Activation of serotonin receptors promotes microglial injury-induced motility but attenuates phagocytic activity. Brain, Behavior, and Immunity 26, 419–428 (2012).

47. Ullmer, C., Schmuck, K., Kalkman, H. O. & Lübbert, H. Expression of serotonin receptor mRNAs in blood vessels. FEBS Letters 370, 215–221 (1995).

48. Leysen, J. E. 5-HT2 Receptors. Current Drug Targets - CNS & Neurological Disorders 3, 11–26 (2004).

49. Paolucci, M., Altamura, C. & Vernieri, F. The Role of Endothelial Dysfunction in the Pathophysiology and Cerebrovascular Effects of Migraine: A Narrative Review. J Clin Neurol 17, 164–175 (2021).

50. Kettenmann, H., Kirchhoff, F. & Verkhratsky, A. Microglia: new roles for the synaptic stripper. Neuron 77, 10–18 (2013).

51. Kolodziejczak, M. et al. Serotonin Modulates Developmental Microglia via 5-HT 2B Receptors: Potential Implication during Synaptic Refinement of Retinogeniculate Projections. ACS Chem. Neurosci. 6, 1219–1230 (2015).

52. Genovese, G. et al. Increased burden of ultra-rare protein-altering variants among 4,877 individuals with schizophrenia. Nat Neurosci 19, 1433–1441 (2016).

53. Bevilacqua, L. et al. A population-specific HTR2B stop codon predisposes to severe impulsivity. Nature 468, 1061–1066 (2010).

54. Pitychoutis, P. M., Belmer, A., Moutkine, I., Adrien, J. & Maroteaux, L. Mice Lacking the Serotonin Htr2B Receptor Gene Present an Antipsychotic-Sensitive Schizophrenic-Like Phenotype. Neuropsychopharmacology 40, 2764–2773 (2015).

55. Tikkanen, R. et al. Impulsive alcohol-related risk-behavior and emotional dysregulation among individuals with a serotonin 2B receptor stop codon. Transl Psychiatry 5, e681 (2015).

56. Devroye, C., Cathala, A., Piazza, P. V. & Spampinato, U. The central serotonin2B receptor as a new pharmacological target for the treatment of dopamine-related neuropsychiatric disorders: Rationale and current status of research. Pharmacol Ther 181, 143–155 (2018).

57. Seamans, J. K. & Yang, C. R. The principal features and mechanisms of dopamine modulation in the prefrontal cortex. Progress in Neurobiology 74, 1–58 (2004).

58. Tunbridge, E. M., Harrison, P. J. & Weinberger, D. R. Catechol-o-Methyltransferase, Cognition, and Psychosis: Val158Met and Beyond. Biological Psychiatry 60, 141–151 (2006).

59. Diamond, A. Consequences of Variations in Genes that affect Dopamine in Prefrontal Cortex. Cerebral Cortex 17, i161–i170 (2007).

60. Pardiñas, A. F. et al. Common schizophrenia alleles are enriched in mutation-intolerant genes and in regions under strong background selection. Nat Genet 50, 381–389 (2018).

61. Allen, N. C. et al. Systematic meta-analyses and field synopsis of genetic association studies in schizophrenia: the SzGene database. Nat Genet 40, 827–834 (2008).

62. Sun, J., Kuo, P.-H., Riley, B. P., Kendler, K. S. & Zhao, Z. Candidate genes for schizophrenia: a survey of association studies and gene ranking. Am J Med Genet B Neuropsychiatr Genet 147B, 1173–1181 (2008).

63. Lewis, C. M. et al. Genome scan meta-analysis of schizophrenia and bipolar disorder, part II: Schizophrenia. Am J Hum Genet 73, 34–48 (2003).

64. Ng, M. Y. M. et al. Meta-analysis of 32 genome-wide linkage studies of schizophrenia. Mol Psychiatry 14, 774–785 (2009).

65. Howrigan, D. P. et al. Exome sequencing in schizophrenia-affected parent-offspring trios reveals risk conferred by protein-coding de novo mutations. Nat Neurosci 23, 185–193 (2020).

66. Rothmond, D. A., Weickert, C. S. & Webster, M. J. Developmental changes in human dopamine neurotransmission: cortical receptors and terminators. BMC Neurosci 13, 18 (2012).

67. Roberts, D. A., Balderson, D., Pickering-Brown, S. M., Deakin, J. F. & Owen, F. The relative abundance of dopamine D4 receptor mRNA in post mortem brains of schizophrenics and controls. Schizophr Res 20, 171–174 (1996).

68. Mulcrone, J. & Kerwin, R. W. No difference in the expression of the D4 gene in post-mortem frontal cortex from controls and schizophrenics. Neurosci Lett 219, 163–166 (1996).

69. Stefanis, N. C., Bresnick, J. N., Kerwin, R. W., Schofield, W. N. & McAllister, G. Elevation of D4 dopamine receptor mRNA in postmortem schizophrenic brain. Brain Res Mol Brain Res 53, 112–119 (1998).

70. Meador-Woodruff, J. H. et al. Dopamine receptor mRNA expression in human striatum and neocortex. Neuropsychopharmacology 15, 17–29 (1996).

71. Yizhar, O. et al. Neocortical excitation/inhibition balance in information processing and social dysfunction. Nature 477, 171–178 (2011).

72. Mehta, U. M. et al. A transdiagnostic evaluation of cortical inhibition in severe mental disorders using Transcranial Magnetic Stimulation. Journal of Psychiatric Research 143, 364–369 (2021).

73. Wang, X., Zhong, P. & Yan, Z. Dopamine D 4 Receptors Modulate GABAergic Signaling in Pyramidal Neurons of Prefrontal Cortex. J. Neurosci. 22, 9185–9193 (2002).

74. Mulligan, R. C., Kristjansson, S. D., Reiersen, A. M., Parra, A. S. & Anokhin, A. P. Neural correlates of inhibitory control and functional genetic variation in the dopamine D4 receptor gene. Neuropsychologia 62, 306–318 (2014).

75. Tan, T. et al. Stress Exposure in Dopamine D4 Receptor Knockout Mice Induces Schizophrenia-Like Behaviors via Disruption of GABAergic Transmission. Schizophrenia Bulletin 45, 1012–1023 (2019).

76. Rubinstein, M. et al. Mice lacking dopamine D4 receptors are supersensitive to ethanol, cocaine, and methamphetamine. Cell 90, 991–1001 (1997).

77. Ptáček, R., Kuželová, H. & Stefano, G. B. Dopamine D4 receptor gene DRD4 and its association with psychiatric disorders. Med Sci Monit 17, RA215–RA220 (2011).

78. Tunbridge, E., Burnet, P. W. J., Sodhi, M. S. & Harrison, P. J. Catechol-o-methyltransferase (COMT) and proline dehydrogenase (PRODH) mRNAs in the dorsolateral prefrontal cortex in schizophrenia, bipolar disorder, and major depression. Synapse 51, 112–118 (2004).

79. Walshe, M. et al. The Association between COMT, BDNF, and NRG1 and Premorbid Social Functioning in Patients with Psychosis, Their Relatives, and Controls. Scientifica 2012, 1–6 (2012).

80. Srivastava, K. et al. Effect of Catechol-O-Methyltransferase Genotype Polymorphism on Neurological and Psychiatric Disorders: Progressing Towards Personalized Medicine. Cureus 13, e18311 (2021).

81. Dauvilliers, Y., Tafti, M. & Landolt, H. P. Catechol-O-methyltransferase, dopamine, and sleep-wake regulation. Sleep Medicine Reviews 22, 47–53 (2015).

82. Marshall, C. R. et al. Contribution of copy number variants to schizophrenia from a genome-wide study of 41,321 subjects. Nat Genet 49, 27–35 (2017).

83. Morris, H. M., Hashimoto, T. & Lewis, D. A. Alterations in somatostatin mRNA expression in the dorsolateral prefrontal cortex of subjects with schizophrenia or schizoaffective disorder. Cereb. Cortex 18, 1575–1587 (2008).

84. Hashimoto, T. et al. Conserved regional patterns of GABA-related transcript expression in the neocortex of subjects with schizophrenia. Am J Psychiatry 165, 479–489 (2008).

85. Mellios, N. et al. Molecular determinants of dysregulated GABAergic gene expression in the prefrontal cortex of subjects with schizophrenia. Biol Psychiatry 65, 1006–1014 (2009).

86. Van Derveer, A. B. et al. A Role for Somatostatin-Positive Interneurons in Neuro-Oscillatory and Information Processing Deficits in Schizophrenia. Schizophr Bull 47, 1385–1398 (2021).

87. Rudy, B., Fishell, G., Lee, S. & Hjerling-Leffler, J. Three groups of interneurons account for nearly 100% of neocortical GABAergic neurons. Dev Neurobiol 71, 45–61 (2011).

88. Melchitzky, D. S. & Lewis, D. A. Dendritic-targeting GABA neurons in monkey prefrontal cortex: comparison of somatostatin- and calretinin-immunoreactive axon terminals. Synapse 62, 456–465 (2008).

89. Robinson, S. L. & Thiele, T. E. A role for the neuropeptide somatostatin in the neurobiology of behaviors associated with substances abuse and affective disorders. Neuropharmacology 167, 107983 (2020).

90. Waldvogel, H. J. et al. Distribution of gephyrin in the human brain: an immunohistochemical analysis. Neuroscience 116, 145–156 (2003).

91. Choii, G. & Ko, J. Gephyrin: a central GABAergic synapse organizer. Exp Mol Med 47, e158 (2015).

92. Akbarian, S. et al. Gene expression for glutamic acid decarboxylase is reduced without loss of neurons in prefrontal cortex of schizophrenics. Arch Gen Psychiatry 52, 258–266 (1995).

93. Verdurand, M., Fillman, S. G., Weickert, C. S. & Zavitsanou, K. Increases in [3H]muscimol and [3H]flumazenil binding in the dorsolateral prefrontal cortex in schizophrenia are linked to α4 and γ2S mRNA levels respectively. PLoS One 8, e52724 (2013).

94. Benes, F. M., Vincent, S. L. & Todtenkopf, M. The density of pyramidal and nonpyramidal neurons in anterior cingulate cortex of schizophrenic and bipolar subjects. Biol Psychiatry 50, 395–406 (2001).

95. Pekny, M. & Nilsson, M. Astrocyte activation and reactive gliosis. Glia 50, 427–434 (2005).

96. Pekny, M., Wilhelmsson, U. & Pekna, M. The dual role of astrocyte activation and reactive gliosis. Neuroscience Letters 565, 30–38 (2014).

97. Kanemaru, K. et al. Calcium-dependent N-cadherin up-regulation mediates reactive astrogliosis and neuroprotection after brain injury. Proc. Natl. Acad. Sci. U.S.A. 110, 11612–11617 (2013).

98. Kang, J., Jiang, L., Goldman, S. A. & Nedergaard, M. Astrocyte-mediated potentiation of inhibitory synaptic transmission. Nat Neurosci 1, 683–692 (1998).

99. Deemyad, T., Lüthi, J. & Spruston, N. Astrocytes integrate and drive action potential firing in inhibitory subnetworks. Nat Commun 9, 4336 (2018).

100. Mariotti, L., Losi, G., Sessolo, M., Marcon, I. & Carmignoto, G. The inhibitory neurotransmitter GABA evokes long-lasting Ca^2+^ oscillations in cortical astrocytes. Glia 64, 363–373 (2016).

101. Nilsson, M., Eriksson, P. S., Rönnbäck, L. & Hansson, E. GABA induces Ca2+ transients in astrocytes. Neuroscience 54, 605–614 (1993).

102. Yoon, B. et al. Glial GABA, synthesized by monoamine oxidase B, mediates tonic inhibition. J Physiol 592, 4951–4968 (2014).

103. Heo, J. Y. et al. Aberrant Tonic Inhibition of Dopaminergic Neuronal Activity Causes Motor Symptoms in Animal Models of Parkinson’s Disease. Current Biology 30, 276-291.e9 (2020).

104. Boddum, K. et al. Astrocytic GABA transporter activity modulates excitatory neurotransmission. Nat Commun 7, 13572 (2016).

105. Mederos, S. & Perea, G. GABAergic-astrocyte signaling: A refinement of inhibitory brain networks. Glia 67, 1842–1851 (2019).

106. Besse, A. et al. The GABA transaminase, ABAT, is essential for mitochondrial nucleoside metabolism. Cell Metab 21, 417–427 (2015).

107. Hegde, A. U. et al. GABA Transaminase Deficiency With Survival Into Adulthood. J Child Neurol 34, 216–220 (2019).

108. Jager, A. et al. Cortical control of aggression: GABA signalling in the anterior cingulate cortex. European Neuropsychopharmacology 30, 5–16 (2020).

109. Verberk, I. M. W. et al. Serum markers glial fibrillary acidic protein and neurofilament light for prognosis and monitoring in cognitively normal older people: a prospective memory clinic-based cohort study. The Lancet Healthy Longevity 2, e87–e95 (2021).

