## Supplemental Methods and Results for "Neurotransmission-Related Gene Expression in the Frontal Pole (Brodmann Area 10) is Altered in Subjects with Bipolar Disorder and Schizophrenia"

1 Michigan Neuroscience Institute. University of Michigan, Ann Arbor, MI, United States America,
2 University of California-Irvine, Irvine, CA, United States of America,
3 HudsonAlpha Institute for Biotechnology, Huntsville,
AL, United States of America,
4 Cornell University, New York, NY, United States of America,
5 Stanford University, Palo Alto, CA, United States of America

*Shared first authorship

**Correspondence:**

Dr. Megan Hagenauer
Michigan Neuroscience Institute
University of Michigan
205 Zina Pitcher Pl.
Ann Arbor, MI, 48109, USA


Meta-analysis: 40
Comparisons with Previous Published Results: 41
Comparison with RNA-Seq Results from the Dorsolateral Prefrontal Cortex (DLPFC) and Neighboring Regions 41
Comparison with Microarray Results from DLPFC Grey-Matter Focused Dissections 42
Comparison with Previous Microarray Results from Other Psychiatric Disorders 42
Examining sensitivity of qPCR for measuring low-level expression: 43
Exploring Functional Patterns in the Differential Expression Results: 44
Annotation with General Functional Information: 44
Exploring Overlap Between our Results and Genetic Variants Associated With Diagnosis: 44
Exploring Overlap Between our Results and Stress-Related Differential Gene Expression: 44
Exploring Overlap Between our Results and Drug-Related Differential Gene Expression: 45
Supplementary Results 47
Full statistical reporting for results 47
Sensitivity to low-level expression 47
Similarity to Effects in Previous BA10 Microarray Studies 49
BA10 Microarray Meta-Analysis 52
Exploratory Analyses: Factors Contributing to Diagnosis-Related Gene Expression 53
Overlap Between our Diagnosis Results and Therapeutic-Related Differential Gene Expression 53
Overlap Between our Diagnosis Results and Differential Gene Expression Related to Substances of Abuse 55
Overlap Between our Diagnosis Results and Symptom-Related Differential Gene Expression 59
Overlap Between our Diagnosis Results and Stress-Related Differential Gene Expression 62
Overlap Between our Diagnosis Results and Genetic Variants Associated With Schizophrenia 62
Supplemental Table Legends 64
References 67

**Page Numbers for Supplemental Figures**

Figure S 1. Batch related variables in relationship to diagnosis. 6
Figure S 2. Sample sizes for psychological autopsy variables included in the exploratory qPCR analysis. 18
Figure S 3. Image illustrating the sampling area used in this study. 20

Figure S 15. The effects of co-variates (pH, Age, RNA Integrity) in our BA10 qPCR study strongly replicate the effects of co-variates in our re-analysis of two BA10 microarray studies. 50

#### Supplementary Methods

All analyses were performed in R (Rstudio v1.0.153, R v3.4.1) and the code has been publicly released on Github at (*https://github.com/hagenaue/Adriana_FrontalPole, https://github.com/hagenaue/FrontalPole_Microarray*). All statistical tests were performed using two-sided p-value calculations.

##### qPCR Experiments

***Subject Recruitment and Brain Bank Protocols:*** Brain tissue samples from patients with Bipolar Disorder and Schizophrenia as well as non-psychiatric controls were collected through the University of California-Irvine Pritzker Brain Donor Program.

To obtain brain donations, the University of California-Irvine Pritzker Brain Donor Program embedded brain donation staff within the investigative and autopsy room staff at the local Coroner's Office. The brain donor coordinator screened deaths daily prior to autopsy. Decedents were screened for inclusion/exclusion criteria that included (but was not limited to) history of psychiatric disorder, infectious disease, manner of death, time since death, as well as other medical and logistical factors. The severity and duration of physiological stress at the time of death was represented by an agonal factor score for each subject (ranging from 0±4, with 4 representing severe physiological stress [1–3]). All subjects included in the current study were deemed to have a fast death (<1 hr) with minimal physiological stress (agonal factor 0). Subjects' gender, race, or ethnicity did not play a factor in who was approached for donation. Age range was restricted to between 18 years old and 85 years old (older subjects were occasionally procured if there was no clinical evidence of dementia and evidence that they were high functioning).

Informed consent and medical information for each subject were obtained through next of kin. Subjects were recruited by cold-calling legal next-of-kin within hours after death. Although we did not keep exact records of declination, we estimate our procurement rates at 30-45%. There was no measurable way to determine factors for declined participation because it was impossible to follow up declines with a survey. Informal conversation with next-of-kin suggested that the decision to participate was sometimes based upon the decedent's expressed pro-donation sentiments.

The brains were then extracted during the autopsy, sliced on the coronal plane into 0.75 cm thick slabs, then snap-frozen for storage at -80 ℃ until microdissection. We calculated the interval between the estimated time of death and the freezing of the brain tissue (the postmortem interval or PMI) using coroner records. A neuropathology report was generated from examination of digital images of fresh brain slices immediately after autopsy to rule out neurological illness. We measured the pH of cerebellar tissue to indicate the extent of oxygen deprivation experienced around the time of death [1–3]. For inclusion in the current study, pH was intended to be >6.5, but one SCHIZ subject was missing brain pH data but otherwise had high quality tissue and RNA metrics. This missing pH datapoint was replaced by average pH for analysis purposes. Subject recruitment for the current study took place in nine cohorts between 1993-2013 (**Table S 2, Figure S 1).**

**A. Brain Bank Cohort**

| | Control | BP | Schiz |
|---|---|---|---|
| Dep Cohort 1 | 2 | 2 | 0 |
| Schiz Cohort 2 | 4 | 0 | 5 |
| Dep Cohort 5 | 1 | 4 | 0 |
| Dep Cohort 6 | 0 | 2 | 2 |
| Cohort 7 | 0 | 6 | 6 |
| Cohort 8 | 0 | 2 | 4 |
| Cohort 11 | 6 | 1 | 3 |
| Cohort 12 | 9 | 4 | 2 |
| Cohort 13 | 5 | 0 | 2 |

**B. Dissection/Extraction Group**

| | Dissection/Extraction Group | | | | | | | | | | | | |
|---|---|---|---|---|---|---|---|---|---|---|---|---|---|
| | 1 | 2 | 3 | 4 | 5 | 6 | 7 | 8 | 9 | 10 | 11 | 12 | 6v2 |
| Control | 3 | 1 | 2 | 2 | 2 | 2 | 3 | 2 | 2 | 2 | 2 | 3 | 0 |
| BP | 1 | 2 | 2 | 2 | 2 | 0 | 1 | 2 | 2 | 2 | 2 | 2 | 1 |
| Schiz | 2 | 1 | 2 | 2 | 2 | 2 | 2 | 1 | 1 | 2 | 2 | 2 | 1 |

**C. GABA-GLU Dataset: qPCR Card**

| | Card 1.eds | Card 2.eds | Card 3.eds | Card 4.eds | Card 5.eds | Card 6.eds | Card 7.eds | Card 8.eds | Card 9.eds | Card 10.eds | Card 11.eds | Card 12.eds | Card 13.eds | Card 14.eds | Card 15.eds | Card 16.eds | Card 17.eds | Card 18.eds |
|---|---|---|---|---|---|---|---|---|---|---|---|---|---|---|---|---|---|---|
| Control | 1 | 2 | 1 | 1 | 1 | 1 | 1 | 2 | 0 | 0 | 2 | 2 | 1 | 1 | 2 | 2 | 1 | 1 |
| BP | 1 | 1 | 1 | 1 | 1 | 1 | 1 | 1 | 2 | 2 | 1 | 1 | 2 | 2 | 0 | 0 | 1 | 1 |
| Schiz | 1 | 1 | 2 | 2 | 1 | 1 | 1 | 1 | 2 | 2 | 1 | 1 | 1 | 0 | 2 | 2 | 2 | 2 |

| | Card 19.eds | Card 20.eds | Card 21.eds | Card 22.eds | Card 23.eds | Card 24.eds | Card 25.eds | Card 26.eds | Card 27.eds | Card 28.eds | Card 29.eds | Card 30.eds | Card 31.eds | Card 32.eds | Card 33.eds | Card 34.eds | Card 35.eds | Card 36.eds |
|---|---|---|---|---|---|---|---|---|---|---|---|---|---|---|---|---|---|---|
| Control | 1 | 2 | 2 | 2 | 1 | 1 | 2 | 2 | 0 | 0 | 2 | 2 | 2 | 2 | 1 | 1 | 2 | 2 |
| BP | 1 | 0 | 0 | 0 | 2 | 2 | 1 | 1 | 2 | 2 | 1 | 1 | 1 | 1 | 2 | 2 | 1 | 1 |
| Schiz | 1 | 1 | 2 | 2 | 1 | 1 | 0 | 0 | 2 | 2 | 1 | 1 | 1 | 1 | 1 | 1 | 1 | 1 |

**D. DA5HT Dataset: qPCR Card**

| | Card 39.eds | Card 40.eds | Card 41.eds | Card 42.eds | Card 43.eds | Card 44.eds | Card 45.eds | Card 46.eds | Card 47.eds | Card 48.eds | Card 49.eds | Card 50.eds | Card 51.eds | Card 52.eds | Card 53.eds | Card 54.eds | Card 55.eds | Card 56.eds | Card 57.eds | Card 58.eds |
|---|---|---|---|---|---|---|---|---|---|---|---|---|---|---|---|---|---|---|---|---|
| Control | 3 | 3 | 3 | 3 | 3 | 3 | 2 | 2 | 4 | 4 | 3 | 3 | 2 | 2 | 4 | 4 | 2 | 2 | 4 | 4 |
| BP | 2 | 2 | 3 | 3 | 2 | 2 | 2 | 2 | 1 | 1 | 3 | 3 | 3 | 3 | 2 | 2 | 3 | 3 | 2 | 2 |
| Schiz | 3 | 3 | 1 | 1 | 3 | 3 | 4 | 4 | 3 | 3 | 1 | 1 | 3 | 3 | 2 | 2 | 3 | 3 | 2 | 2 |

***Figure S 1. Batch related variables in relationship to diagnosis.***
***A)** Brain bank cohorts, defined by the brain bank at UC-Irvine. Subject recruitment, primary (slab) dissection, and clinical information gathering took place loosely around the same time for subjects in each cohort. Since our study included a secondary dissection that was balanced across diagnosis and cohort, brain bank cohorts are unlikely to influence gene expression. **B)** Diagnosis was balanced as much as possible across dissection/extraction group: These groups of subjects received their secondary (Brodmann Area 10, BA10) dissection and RNA extraction in tandem. These groups were carefully balanced by diagnosis, age, pH, PMI, and cohort, although two samples from one group (6) later needed re-extraction (6v2). **C)** Diagnosis was balanced as much as possible across qPCR card. Each qPCR Card in the GABA-GLU dataset included 4 samples, with replicate samples included on numerically adjacent cards (e.g., Card 1 & Card 2). **D)** Each qPCR Card in the DA5HT dataset included 8 samples, with replicate samples included on numerically adjacent cards (e.g., Card 39 & Card 40). Eight subjects were run in quadruplicate, with their extra samples included on Card 57 and 58.*

***Psychological Autopsy:*** Once subjects were procured, there was an in-depth psychological autopsy process before inclusion into a study cohort that was completed by a trained clinician (David Walsh, Psy.D.). The psychological autopsy included reviewing coroner or medical examiner records, medical and psychiatric records, and a detailed interview with the next-of-kin for symptom signs, duration, and severity that might not be detailed in medical records **(Appendix 1).** Schizophrenia and Bipolar Disorder cases were confirmed to meet diagnostic criteria from the Diagnostics and Statistical Manual of Mental Disorders [4]. In the control group, there was no evidence of neurological or psychiatric disorders nor was there a history of said illnesses in their first-degree relatives.

***Appendix 1.***

### Psychological Autopsy Protocol

**Identifying Information:**

Name: ______________________________________________________HSB#: __________________

**Next of Kin:**

*Name:* ___________________________________________ *Phone:* ______________________________

*Address:*______________________________________________________________________________

*Primary Informant-*

*Information provided by:* _____________________________ *Last contact with decedent:* __________________

*Frequency of interaction: daily / weekly / monthly / quarterly / semiannually / annually / less than annually*

*Interactions were: in person / by phone*

*Secondary Informant-*

*Information provided by:* _____________________________ *Last contact with decedent:* __________________

*Frequency of interaction: daily / weekly / monthly / quarterly / semiannually / annually / less than annually*

*Interactions were: in person / by phone*

**Consented by (tissue source):** UCI (OC Coroner) / DRCC (Transplant line) / Orange County Eye Bank / Other

**Demographic Information:**

*Gender: M / F Date of birth:*____________ *Place of birth:*_________________________

*Where was his/her mother born?* ________________

*Where was his/her maternal grandmother born (mother's mother)?* ________________

*Where was his/her maternal grandfather born (mother's father)?* ________________

*Where was his/her father born?* ________________

*Where was his/her paternal grandmother born (father's mother)?* ________________

*Where was his/her paternal grandfather born (father's father)?* ________________

*Education (highest level attained):* _______ *years*

*Race:*

*African American / Asian / Caucasian / Hispanic / Pacific Islander / Other:* _________

*Age at time of death:* _____

*Occupational status at death:*

*Employed / Unemployed / Retired / Student / Disabled / Unknown*

*Marital status at time of death:*

*Married / Separated / Divorced / Widowed / Never married / Significant Other*

**Location of Death and Processing Times:**

*Hospitalized (or ER) at time of death* *Yes / No*

Hospital lab reports available Yes / No

*Location:* ___________________________________________________________________________

Attending physician:_____________________________________________________________________

Psychological Autopsy Protocol (Walsh, Bunney, et al 2005)


Coroner / Investigator assigned to case:________________________________ O.C. Case #: ____________________

Date of death: _______________ Time of death: _______________ Date of Autopsy: _________________

Time of Autopsy: ____________ Hours to cold: ________________ Hours to ice: _____________________

Final PMI:_________________

#### Autopsy Report:

Was a toxicology screen completed by coroner or emergency room personnel Yes / No

Blood Alcohol Level: ___________

Toxicology Results: ________________________________________________________________________

#### Mode of Death (copy of death certificate in file):

Source (circle one): Orange County Sheriff-Coroner / Primary Medical Physician / Private Autopsy

Cause: Suicide / Accident / Sudden medical condition / Long-term medical illness / Undetermined or Accidental Overdose / Undetermined

Details: ________________________________________________________________________________

__________________________________________________________________________________________

__________________________________________________________________________________________

#### Agonal Duration (Hardy et al, 1985)

| | AFS |
|---|---|
| (0) Violent fast death: Death in these cases were shootings, blunt force trauma, or asphyxia (accidental, homicidal, or suicidal). Death occurred in 0 – 10 minutes. | 0 |
| (1) Fast death of natural causes: Sudden, unexpected death of people who had apparently been reasonably healthy. The most frequent c.o.d. in this group was myocardial infarction. Most of these cases died at home or in ER. Death occurred in more than 10 minutes but less than 1 hour. | 0 |
| (2) Intermediate death: Patients who were ill but death was unexpected. They could neither be classified as sudden or slow deaths. Most died in hospital. Death occurred in more than 1 hour but less than 24 hours. | 1 |
| (3) Slow death: Death following a long illness with prolonged terminal phase. Typically patients died from cancers, CVD, bronchopneumonia. Most died in hospital or on hospice. Death occurred in more than 24 hours. | 1 |

#### Agonal Risk Factors (Johnston et al, 1997; Harrison et al, 1991)

| | No = 0 / Yes = 1 |
|---|---|
| Medical records and/or cause of death indicate patient was in a coma immediately prior to death (note duration ________). | 0 / 1 |
| Medical records and/or cause of death indicate patient experienced medical conditions (such as infection, sepsis) resulting in temperature above 104 F immediately prior to death. | 0 / 1 |
| Medical records and/or cause of death indicate patient experienced renal failure and/or multi-organ failure immediately prior to death. | 0 / 1 |
| Medical records and/or cause of death indicate patient experienced severe head injury (open fracture) immediately prior to death. | 0 / 1 |
| Medical records and/or cause of death indicate patient experienced hypoxia immediately prior to death. | 0 / 1 |
| Medical records and/or cause of death indicate patient was diagnosed as brain dead prior to death. | 0 / 1 |

Medical records and/or cause of death indicate patient was on a respirator immediately prior to death (note duration ______). Respirator should be regarded differently than CPR efforts (bagged). 0 / 1

Medical records and/or cause of death indicate a seizure immediately prior to death. 0 / 1

**Agonal Factor Score (AFS; Tomita, Vawter, Walsh, et al, 2004) =** ________

***Prescription Medications:*** *None / Yes, medications as listed:*

| Medication | *t.o.d. | Dosage | Date Rx | Medication | *t.o.d. | Dosage | Date Rx |
|---|---|---|---|---|---|---|---|

*Records indicate decedent on medication at time of death. All other notations indicate medication history.

**Medical History (major / chronic illnesses, major surgeries, etc):**

*Do you know if he/she had any medical conditions?*

______________________________________________________________________

______________________________________________________________________

______________________________________________________________________

______________________________________________________________________

*Patient suffered from any communicable diseases (ie: hepatitis, tuberculosis, HIV)* *Yes / No*

*Patient suffered from any of the following medical conditions: cancer, head injury, brain tumor, seizures, epilepsy, stroke, memory problems, other (specify):______________________* *Yes / No*

*Patient ever diagnosed with a learning disability, developmental disorder, mental retardation* *Yes / No*

*Patient ever receive a brain scan (MRI, CAT, PET)* *Yes / No*

***Prior Medical Hospitalizations:***

| *Date* | *Facility* | *Reason for Admission* | *Diagnosis* | *LOS* |
|---|---|---|---|---|

Total hospitalizations _______

Records indicate any medical hospitalizations secondary to suicide attempts Yes / No

Details: ______________________________________________________________________________
______________________________________________________________________________
______________________________________________________________________________
______________________________________________________________________________

##### *Psychiatric History:*

*Age of onset of psychiatric symptoms (circle one):*

*Not applicable / 0-12 / 13-19 / 20-30 / 31-40 / 41-50 / 51-60 / 61-70 / 71+*

*Premorbid social adjustment (How was he/she functioning prior to onset of his illness?)*

*Not applicable / Appropriate / Poor*

*History of suicide attempts?* *Yes / No*

##### *Psychiatric Hospitalizations:*

*Was he/she ever in a psychiatric facility?*

| *Date* | *Facility* | *Physician* | *Reason for Admission* | *Diagnosis* | *LOS* |
|---|---|---|---|---|---|

*Total psychiatric hospitalizations* _______

*Details:* ______________________________________________________________________________
______________________________________________________________________________
______________________________________________________________________________

##### *Outpatient Mental Health Treatment:*

| *Date* | *Provider* | *Reason for Treatment* | *Frequency / Duration* |
|---|---|---|---|

*Details:* ______________________________________________________________________________
______________________________________________________________________________
______________________________________________________________________________

***Substance Use:***

| ***Substance*** | ***Most recent use*** | ***Duration*** | ***Notes*** | ***Pattern*** |
|---|---|---|---|---|
| *Alcohol* | | | | *Not indicated*<br>*Use*<br>*Abuse @ t.o.d.*<br>*Hx of abuse*<br>*Hx Use* |
| *Stimulants (methamphetamine, methylphenidate)* | | | | *Not indicated*<br>*Use*<br>*Abuse @ t.o.d.*<br>*Hx of abuse*<br>*Hx Use* |
| *Sedatives (barbiturates, benzodiazapines)* | | | | *Not indicated*<br>*Use*<br>*Abuse @ t.o.d.*<br>*Hx of abuse*<br>*Hx Use* |
| *Cannabis*<br>*(marijuana, hashish)* | | | | *Not indicated*<br>*Use*<br>*Abuse @ t.o.d.*<br>*Hx of abuse*<br>*Hx Use* |
| *Cocaine* | | | | *Not indicated*<br>*Use*<br>*Abuse @ t.o.d.*<br>*Hx of abuse*<br>*Hx Use* |
| *Hallucinogens (Ecstasy, PCP, LSD, mescaline)* | | | | *Not indicated*<br>*Use*<br>*Abuse @ t.o.d.*<br>*Hx of abuse*<br>*Hx Use* |
| *Opiods*<br>*(heroin)* | | | | *Not indicated*<br>*Use*<br>*Abuse @ t.o.d.*<br>*Hx of abuse*<br>*Hx Use* |
| *Inhalants*<br>*(Paint, glue,*<br>*propellants)* | | | | *Not indicated*<br>*Use*<br>*Abuse @ t.o.d.*<br>*Hx of abuse*<br>*Hx Use* |
| *Tobacco* | | | | *Not indicated*<br>*Use @ t.o.d.*<br>Hx of use |
| *Other substances* | | | | *Not indicated*<br>*Use*<br>*Abuse @ t.o.d.*<br>*Hx of abuse*<br>*Hx Use* |

***Queries for alcohol and drug use*** *(Ask informant about decedent's use of all substances listed above)***:**

| | |
|---|---|
| *Was there ever a time when you thought he/she was drinking too much* | *Yes / No* |
| *Was there ever a time when you were concerned about his/her drug use* | *Yes / No* |
| *Did his/her alcohol or drug use ever interfere with work* | *Yes / No* |
| *Did his/her alcohol or drug use ever interfere with relationships* | *Yes / No* |
| *Did he/she ever try to stop drinking and/or using drugs* | *Yes / No* |
| *Do you know if he/she was ever hospitalized for chemical dependency* | *Yes / No* |

***Queries for psychotic disorder*** *(Based on SCID; First, et al, 1995)***:**

| | |
|---|---|
| **Did he/she think people talked about him/her or took special notice of him/her* | *Yes / No* |
| *Did he/she ever report that he/she received special messages from the television, radio or newspaper* | *Yes / No* |
| **Did he/she believe that people went out of their way to make life difficult for him/her* | *Yes / No* |
| **Did he/she believe that anyone was trying to hurt him/her* | *Yes / No* |
| **Did he/she feel especially important in some way* | *Yes / No* |
| *Did he/she ever complain that his/her body had changed (other than normal weight changes, etc.)* | *Yes / No* |
| *Did he/she ever report that someone was controlling his/her actions or thoughts* | *Yes / No* |
| *Did he/she ever report that others could hear his/her thoughts* | *Yes / No* |
| **Did he/she ever report hearing things that others could not hear* | *Yes / No* |
| **Did he/she ever report seeing things that others could not see* | *Yes / No* |
| *Did he/she ever report smelling things that others could not smell* | *Yes / No* |
| *Did he/she report feeling strange sensations from his/her body* | *Yes / No* |
| *Was there ever an instance when he/she appeared rigid or would stay in a fixed position for a long time* | *Yes / No* |
| *Did his/her emotional expressions seem flat (no facial expressions, monotone voice)* | *Yes / No* |
| *Were his/her emotional expressions incongruent with what was going on* | *Yes / No* |
| *Was there ever an instance when his/her speech was not understandable or incoherent* | *Yes / No* |
| *In conversations, did he/she abnormally shift from one unrelated topic to another* | *Yes / No* |
| **Did he/she rapidly shift from one emotional state to another* | *Yes / No* |
| **Did he/she have trouble at work, home or with friends because of any of these symptoms* | *Yes / No* |
| **Do you think that alcohol or drugs accounted for these psychotic symptoms (duration / pattern)* | *Yes / No* |
| *Did the decedent ever receive mental health treatment for these symptoms* | *Yes / No* |

*Symptom pattern*

*Age symptoms first experienced (circle one):*

*Not applicable 0-12 / 13-19 / 20-30 / 31-40 / 41-50 / 51-60 / 61-70 / 71+*

*Frequency (circle one):* *Not applicable / Associated with substance abuse / Associated with GMC / Single episode / Cyclical / Chronic*

***Queries for affective disorders*** *(Based on SCID; First, et al, 1995)*:

| | |
|---|---|
| **Was there ever a time when he/she seemed depressed or down* | *Yes / No* |
| **Did you notice that he/she had a significant change in weight* | *Yes / No* |
| **Were there times when his/her appetite significantly decreased / increased* | *Yes / No* |
| **Did he/she report feeling tired all the time or having no energy* | *Yes / No* |
| **Did he/she report having trouble sleeping* | *Yes / No* |
| **Did he/she appear restless, fidgety or unable to sit still* | *Yes / No* |
| **Did he/she ever express feelings of worthlessness* | *Yes / No* |
| **Did he/she ever report that life was hopeless* | *Yes / No* |
| **Did he/she report or appear to have trouble concentrating* | *Yes / No* |
| **Did he/she ever express thoughts of suicide or that he/she would be better off dead* | *Yes / No* |
| **Did he/she ever attempt suicide (check C.O.D.)* | *Yes / No* |
| **Did he/she recently experience any significant loss like a relationship ending or family member dying* | *Yes / No* |
| **Did he/she seemed to be on an emotional high* | *Yes / No* |
| **Was he/she ever so hyper that he/she got into trouble* | *Yes / No* |
| **Was there times when he/she seemed overly irritable (shout at others, start fights)* | *Yes / No* |
| *Did he/she ever say that he/she had any special powers* | *Yes / No* |
| **Was there ever a time when he/she seemed to need less sleep than usual* | *Yes / No* |
| **Were there times when he/she seemed more talkative than usual* | *Yes / No* |
| **Did he/she say that his/her thoughts were racing* | *Yes / No* |
| **Did he/she ever do reckless things like buying unnecessary things, driving recklessly* | *Yes / No* |
| **Did he/she have trouble at work, home or with friends because of any of these symptoms* | *Yes / No* |
| **Do you think that alcohol or drugs accounted for all these mood symptoms (duration / pattern)* | *Yes / No* |
| *Was he/she suffering from any serious medical conditions* | *Yes / No* |
| *Were his/her mood symptoms and psychotic symptoms present simultaneously* | *Yes / No* |
| *Was there any seasonal pattern to the occurrence of his/her mood symptoms (ie: onset winter / spring)* | *Yes / No* |
| *Did he/she ever receive any mental health treatment for these symptoms (query ECT)* | *Yes / No* |

*Symptom Pattern*

*Age symptoms first experienced (circle one):*

*Not applicable / 0-12 / 13-19 / 20-30 / 31-40 / 41-50 / 51-60 / 61-70 / 71+*

*Frequency (circle one):* *Not applicable / Associated with substance abuse / Associated with GMC / Single episode / Cyclical / Chronic*

***Queries for anxiety disorders*** *(Based on SCID; First, et al, 1995)***:**

| | |
|---|---|
| **Did he/she worry about things more than others* | *Yes / No* |
| **Was he/she restless or "on edge" much of the time* | *Yes / No* |
| **Was he/she easily fatigued* | *Yes / No* |
| **Did he/she complain of difficulty concentrating or his/he mind going blank* | *Yes / No* |
| **Did he/she appear irritable* | *Yes / No* |
| **Did he/she experience sleep disturbances* | *Yes / No* |
| *If anxiety symptoms present, did these anxiety symptoms effect him/her suddenly* | *Yes / No* |
| *Did he/she complain of racing / pounding heart / chest pain* | *Yes / No* |
| *Did he/she appear to be trembling or shaking* | *Yes / No* |
| *Did he/she experience sudden chills, hot flushes, sweating* | *Yes / No* |
| *Did he/she complain of shortness of breath, feeling smothered, feeling of choking* | *Yes / No* |
| *Did he/she complain of nausea or other abdominal discomfort* | *Yes / No* |
| *Did he/she complain of numbness / tingling, dizziness, light-headedness, feeling faint* | *Yes / No* |
| *Did he/she express fear of dying, losing control, going crazy as a result of these anxiety episodes* | *Yes / No* |
| *Did he/she express anxiety or discomfort about being in certain places* | *Yes / No* |
| *Did he/she avoid certain activities due to anxiety* | *Yes / No* |
| *Did he/she express persistent, excessive or unreasonable fear about specific objects (ie: insects, snakes) or situations (ie: flying, social activities)* | *Yes / No* |
| *If anxiety symptoms were present, were these anxiety symptoms preceded by any form of traumatic event?* | *Yes / No* |

*(If yes, query for: intrusive thoughts, recurrent dreams, flashback episodes, psychological distress and/or physiological reactivity to reminders of the trauma, avoidance, restricted affect, sleep disturbances, irritability, difficulty concentrating, hypervigilance, exaggerated startle response.)*

*Symptom Pattern*

*Age symptoms first experienced (circle one):*

*Not applicable 0-12 / 13-19 / 20-30 / 31-40 / 41-50 / 51-60 / 61-70 / 71+*

*Frequency (circle one):* *Not applicable / Associated with substance abuse / Associated with GMC /* Single episode / Cyclical / Chronic

**Functional Impairment:**

Medical records, coroner's investigation, or family informant(s) indicate patient's psychiatric symptoms likely caused impairment in (circle all that apply):

Relational / Social Environment / Education / Occupation / Housing / Economic / Access to Health Care / Interactions with Legal System / Other Problems:________

*Did the patient's illness result in interactions with law enforcement: yes / no /*

Details: ________________________________________________________________________________

***Queries for Family History of Mental Illness or Substance Abuse:***

*How many siblings he/she have from the same biological mother and father?* _____

*How many biological children did he /she have?* _____

*How many of his/her parents, siblings or children had problems with major depression, that is, periods lasting two or more weeks when they felt sad, blue or depressed?* _____

*How many of his/her parents, siblings or children had episodes of bipolar depression, that is, symptoms of major depression (above) in addition to symptoms of mania (periods of four days or longer when they became so happy or excited and irritable that it was clearly not normal)?* _____

*How many of his/her parents, siblings or children had problems from the use of alcohol?* _____

*How many of his/her parents, siblings or children had problems from the use of drugs?* _____

*How many of his/her parents, siblings or children had a problem like schizophrenia, such as hearing voices that other people could not hear, or having false beliefs that people were plotting against them?* _____

*How many of his/her parents, siblings or children ever expressed thoughts of suicide?* _____

*How many of his/her parents, siblings or children actually committed suicide?* _____

*How many of his/her parents, siblings or children showed signs of any other emotional problems?* _____

*How many of his/her parents, siblings or children received treatment, including medication, for a mental health problem?* _____

*How many of his/her parents, siblings or children were ever hospitalized due to a psychiatric condition?* _____

*How many of his/her grandparents, aunts or uncles suffered from mental illness?* _____

*How many of his/her grandparents, aunts or uncles were ever hospitalized due to a psychiatric condition?* _____

*Details:* ____________________________________________________________________________________

____________________________________________________________________________________________

**Symptom Summary** [circle all that apply; denote history (Hx) or time of death (t.o.d.)]**:**

Schizophrenia- delusions / hallucinations / disorganized speech / grossly disorganized behavior / catatonic behavior / negative symptoms

Major Depression- depressed mood / decreased interest or pleasure in activities / significant weight fluctuations / appetite changes / sleep disturbances / psychomotor agitation or retardation / fatigue / worthlessness / difficulty concentrating / suicidal ideations / suicide attempt

Mania- elevated or expansive mood / inflated self esteem / decreased sleep / increased talkativeness / flight or ideas or racing thoughts / easily distracted / increased goal-directed activity or psychomotor agitation / reckless behavior

Anxiety- excessive worry / restlessness / fatigue / difficulty concentrating / irritable / sleep disturbances / racing heart / trembling / chills or sweating / shortness breath / nausea / dizziness or faint / avoidance

**Syndrome at time of death** (circle all that apply)**:** Depressed / Manic / Psychotic / Intoxicated / None

**Additional Comments:**

______________________________________________________________________________

______________________________________________________________________________

______________________________________________________________________________

**Provisional Diagnosis based on available medical records, and/or family report:**

Axis I: ____________________________________________________________

____________________________________________________________

Axis II: ____________________________________________________________

Axis III: ____________________________________________________________

____________________________________________________________

Axis IV: Relational / Social Environment / Education / Occupation / Housing / Economic / Access to Health Care / Interactions with Legal System / Other Problems:________

Axis V: Not established.

**Conclusions based on information from the following sources:**

Sources of Information (circle all that apply):

Informant(s) / Coroner - Medical Examiner / Medical Records / Psychiatric Records / Other __________

**Completed by: ____________________________________________________**

**Date completed: __________________________________________________**

Verified by: ____________________________________________________

Date completed: ________________________________________________

Entered by: _____________________________________________________

Date completed: ________________________________________________

***Exploratory Clinical Variables:*** For the sake of exploratory analysis, the results of the psychological autopsy were further summarized as a database of 46 binary variables (True/False, **Figure S 2**, **Table S 2)** overviewing: 1) diagnosis-related symptoms and related behaviors, 2) medication, 3) exposure to alcohol or drugs of abuse. To produce this database, the medication and substance-related variables were compiled from multiple sources, with the likelihood of current usage being defined by one of the following: 1) a positive toxicology result for the substance (only available for some subjects), 2) prescribed medications at time of death (only available for 5 subjects), 3) current substance abuse and/or history of abuse with possible relapse (intoxicated at time of death, suicide), or 4) substances indicated in the detailed manner of death description derived from the medical examiner or coroner's report. This drug information was then converted to simple yes/no binary variables for common classes of drugs: antipsychotics, antidepressants, mood-stabilizers, cannabinoids, hallucinogens, opioids, stimulants, depressants, alcohol and tobacco. Using this operationalization, it is worth noting that the quality of reporting was best for common recreational substances for which usage was likely to be recognized by family members (tobacco, alcohol, stimulants), whereas other substances were more likely to be present in the coroner's report or toxicology only in situations where substances were suspected as contributing to cause of death (overdose, accident, suicide, drug interaction). Therefore, for many substances the documented use of a substance by the deceased individual should be taken as stronger evidence than a lack of evidence of use of a substance. This absence of evidence is most notable in the case of antipsychotic medication, for which only 9 individuals had documented usage. To assess how much lack of certainty might contribute to the effects that we were observing, we also created three ordinal variables based on documented presence (coded as 1), absence (coded as -1), or lack of information (coded as 0) as derived from toxicology reports for Antipsychotics, Antidepressants, and Opioids. These toxicology-based variables ended up producing similar results to those found using our broader definition.

| | Sample Size | | | Percent of Group | | | | |
|---|---|---|---|---|---|---|---|---|
| Exploratory Variable | CTRL | SCHIZ | BP | CTRL | SCHIZ | BP | Total n | Small n |
| Full Sample | 26 | 22 | 21 | 100% | 100% | 100% | 69 | |
| Hallucinogens | 0 | 1 | 2 | 0% | 5% | 10% | 3 | ! |
| Dizziness | 0 | 1 | 3 | 0% | 5% | 14% | 4 | ! |
| Depressants | 0 | 2 | 3 | 0% | 9% | 14% | 5 | ! |
| Shortness_breath | 0 | 1 | 4 | 0% | 5% | 19% | 5 | ! |
| Disorganized_speech | 0 | 6 | 0 | 0% | 27% | 0% | 6 | ! |
| Chest_discomfort | 0 | 2 | 4 | 0% | 9% | 19% | 6 | ! |
| Increased_behavior | 0 | 0 | 6 | 0% | 0% | 29% | 6 | ! |
| Antidepressants | 0 | 4 | 4 | 0% | 18% | 19% | 8 | |
| Avoidance | 0 | 2 | 6 | 0% | 9% | 29% | 8 | |
| Opioids | 0 | 2 | 6 | 0% | 9% | 29% | 8 | |
| Disorganized_beh_or_catatonic | 0 | 9 | 0 | 0% | 41% | 0% | 9 | |
| Antipsychotics | 0 | 5 | 4 | 0% | 23% | 19% | 9 | |
| Cannabinoids | 0 | 4 | 7 | 0% | 18% | 33% | 11 | |
| Expansive_or_grandiose | 0 | 0 | 12 | 0% | 0% | 57% | 12 | |
| Stimulants | 0 | 3 | 10 | 0% | 14% | 48% | 13 | |
| Restless | 0 | 8 | 6 | 0% | 36% | 29% | 14 | |
| Negative_sxs | 0 | 15 | 0 | 0% | 68% | 0% | 15 | |
| Overdose | 0 | 7 | 8 | 0% | 32% | 38% | 15 | |
| Reckless | 0 | 4 | 11 | 0% | 18% | 52% | 15 | |
| Worry | 3 | 6 | 7 | 12% | 27% | 33% | 16 | |
| Interactions_with_legal_system | 1 | 6 | 9 | 4% | 27% | 43% | 16 | |
| Difficulty_concentrating | 0 | 6 | 10 | 0% | 27% | 48% | 16 | |
| Decreased_sleep | 1 | 5 | 10 | 4% | 23% | 48% | 16 | |
| Suicide | 0 | 5 | 11 | 0% | 23% | 52% | 16 | |
| Appetite | 0 | 5 | 11 | 0% | 23% | 52% | 16 | |
| Racing_thoughts | 0 | 4 | 12 | 0% | 18% | 57% | 16 | |
| Agitated | 0 | 8 | 9 | 0% | 36% | 43% | 17 | |
| Tobacco | 3 | 10 | 7 | 12% | 45% | 33% | 20 | |
| Irritable | 1 | 6 | 13 | 4% | 27% | 62% | 20 | |
| Worth_or_helpless | 0 | 5 | 15 | 0% | 23% | 71% | 20 | |
| Talkativeness | 1 | 5 | 15 | 4% | 23% | 71% | 21 | |
| Easily_fatigued | 2 | 10 | 10 | 8% | 45% | 48% | 22 | |
| Alcohol | 1 | 8 | 13 | 4% | 36% | 62% | 22 | |
| Trouble_concentrating | 0 | 8 | 14 | 0% | 36% | 67% | 22 | |
| Weight | 2 | 6 | 15 | 8% | 27% | 71% | 23 | |
| Hallucinations | 0 | 19 | 5 | 0% | 86% | 24% | 24 | |
| Anxiety | 0 | 13 | 11 | 0% | 59% | 52% | 24 | |
| Sleep_irregularities | 2 | 7 | 15 | 8% | 32% | 71% | 24 | |
| Fatigue | 3 | 9 | 15 | 12% | 41% | 71% | 27 | |
| Sleep_disturbances | 2 | 7 | 18 | 8% | 32% | 86% | 27 | |
| Suicidal_ideations | 1 | 7 | 19 | 4% | 32% | 90% | 27 | |
| Delusions | 0 | 20 | 11 | 0% | 91% | 52% | 31 | |
| Psychosis | 0 | 22 | 12 | 0% | 100% | 57% | 34 | |
| Affective | 0 | 18 | 20 | 0% | 82% | 95% | 38 | ! |
| SubstancesOrNot | 5 | 18 | 18 | 19% | 82% | 86% | 41 | ! |
| Depressed_mood | 7 | 16 | 20 | 27% | 73% | 95% | 43 | ! |

***Figure S 2. Sample sizes for psychological autopsy variables included in the exploratory qPCR analysis.***

*The results of the psychological autopsy (**Appendix 1)** were summarized as a database of 46 binary variables (True/False, **Table S 2)** overviewing 1) diagnosis-related symptoms and related behaviors, 2) medication, 3) exposure to alcohol or drugs of abuse. To produce this database, the medication and substance-related variables were compiled from multiple sources, with the likelihood of current usage being defined by one of the following: 1) a positive toxicology result for the substance (only available for some subjects), 2) prescribed medications at time of death (only available for 5 subjects), 3) current substance abuse and/or history of abuse with possible relapse (intoxicated at time of death, suicide), or 4) substances indicated in the detailed manner* *of death description or coroner's report. This drug information was then converted to simple yes/no binary variables for common classes of drugs: antipsychotics, antidepressants, mood-stabilizers, cannabinoids, hallucinogens, opioids, stimulants, depressants, alcohol and tobacco.*

***Representativeness of Sample:*** Overall, in general the samples collected by the UC-Irvine Pritzker brain bank were representative of the available deaths that were brought in for autopsy, but not representative of the general population. These sources of bias were further amplified by our exclusion criteria. For example, in our brain bank, the non-psychiatric (control) females tended to have a higher average age of death than control males, posing greater risk for dementia (an exclusion criteria). Females often died of cancer and had a slow death (>1 hr, an exclusion criteria) whereas more males died at an earlier age of heart disease and were more likely to experience a fast death (<1 hr). Furthermore, within our brain bank, a large percentage of the depressed subjects were suicide completers. More men committed suicide by gunshot wounds to the head (an exclusion criteria due to compromised brain tissue) whereas females were more likely to overdose. These factors and others contributed to the skew present in the demographics in the sample for our current study, which is predominantly Caucasian (91%) and male (88%).

Another potential source of bias worth noting was our exclusion criteria based on brain pH (pH>6.5). We measured the pH of cerebellar tissue as an indication of the extent of oxygen deprivation experienced around the time of death, which is repeatedly implicated as one of the largest sources of variation in post-mortem brain gene expression data [1–3]. However, pH can indicate other biological changes besides hypoxia. For example, there are small consistent decreases in pH associated with BP even in live subjects [5–7] and metabolic changes associated with pH are theorized to play an important role in SCHIZ [8]. Therefore, our exclusion criteria may have ruled out some subjects with decreased pH due to the disorders themselves. That said, the variation in pH associated with hypoxia is a magnitude larger than the variation in pH due to diagnosis, and therefore we felt that this exclusion criteria was necessary to reduce large-magnitude noise in our data.

Altogether, our sample was well-designed to control for the largest sources of noise in human post-mortem gene expression data (manner of death, hypoxia, and other perimortem factors), but not optimal for the analysis of more subtle influences on diagnosis-related gene expression (sex, race).

***Frontal Pole Dissection and RNA Extraction:*** Frontal Pole tissue was collected from 72 subjects (CTRL: *n*=27, BP: *n*=21, SCHIZ: *n*=24). This sample size was determined by the amount of tissue available that met strict quality metrics (agonal factor 0, pH>6.5). Tissue slabs were sent to University of Michigan. Following the receipt of all tissue, subjects were divided into 12 batches for the dissection of frontal pole tissue, RNA extraction and RNA purification procedures, which took place in 2018. These batches were counterbalanced by diagnosis (n=1-3 per diagnosis per group, Fisher's exact test: *p*>0.99)*,* as well as for a variety of other demographic variables (age, pH, PMI, cohort). Following counterbalancing, the procedures themselves were performed by an experimenter blinded to the diagnosis associated with the samples. The extraction for two subjects (from batch 6) failed and was redone (referred to as batch 6v2, **Figure S 1** ).

To dissect the frontal pole, the foremost rostral slab from the left hemisphere was placed on dry ice. The lateral end of the slab was sub-dissected to obtain blocks averaging 500 µg that contained Brodmann area 10 (BA10, **Figure S 3**). The blocks were then wrapped in foil, placed on dry ice, and then stored at −80°C until further processing.

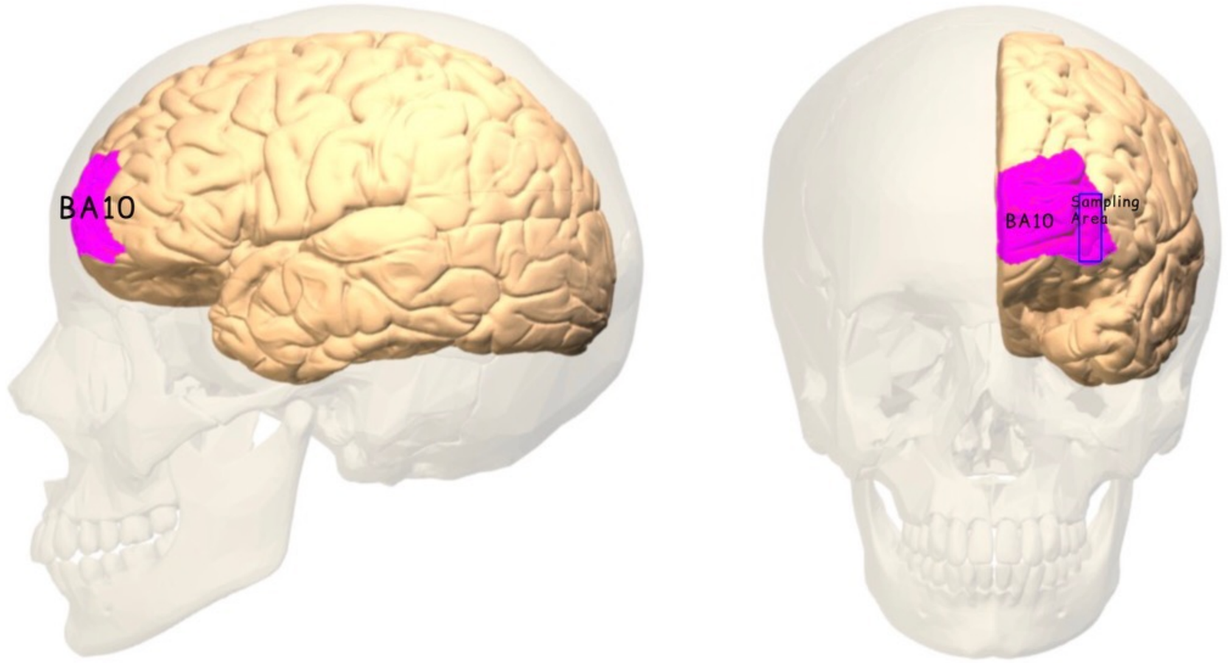

***Figure S 3. Image illustrating the sampling area used in this study.***
*Samples were taken from the first coronal slab of each brain by dissecting a piece of the lateral end of BA10 from the left hemisphere. Image sourced via Wikipedia from BodyParts3D, © The Database Center for Life Science licensed under CC Attribution-Share Alike 2.1 Japan. Modified to change color and add labels.*

Total RNA was extracted using TRIzol™ reagent (ThermoFisher Scientific Cat#15596026/15596018) following the protocol from the manufacturer, with DNA and protein removed by centrifugation. Total RNA was resuspended in DEPC $H_20$ in a volume proportional to the weight of the dissected tissue. The concentration and purity (260/280 and 260/230) of the total RNA was then assessed using a Nanodrop within two separate laboratories (Molecular Behavioral Neuroscience Institute (MBNI, now called Michigan Neuroscience Institute) and DNA Sequencing Core (DSC, now called the Advanced Genomics Core)), and the values averaged. RNA Integrity of the total RNA (RNA Integrity Number or RIN, 28s/18s rRNA ratio) was also assessed using an Agilent Bioanalyzer. Total RNA (100 µL) was then purified using RNeasy® Mini Kit (Qiagen Cat#74106) and the concentration, purity, and integrity of purified RNA was assessed again.

During later quality control steps, the data from three subjects were excluded due to poor RNA quality metrics: two subjects had purified RNA with poor RNA integrity (RIN<5.5) and one subject had low RNA purity and concentration (average 260/230<1, average RNA concentration <120 ng/µL). The RNA metrics from two subjects (batch 6v2) were deemed satisfactory during initial review, but their RNA metric data was lost prior to later analysis. Their RNA metrics were replaced with average values for analysis purposes.

***Assessing final sample characteristics***: The variability and distribution of each of the subject variables in our final sample - including demographics, pre- and post-mortem biological variables, and technical variables - can be seen in **Figure S 4**. The variation of these subject variables across diagnosis groups can be seen in **Figure S 5.** The potential for a confounding relationship with diagnosis (p<0.10) was assessed for each subject variable using one-way

ANOVA (for continuous variables) or Fisher's exact test (for categorical variables). The full descriptive and inferential statistics for each diagnosis relationship can be found in **Table S 4.**

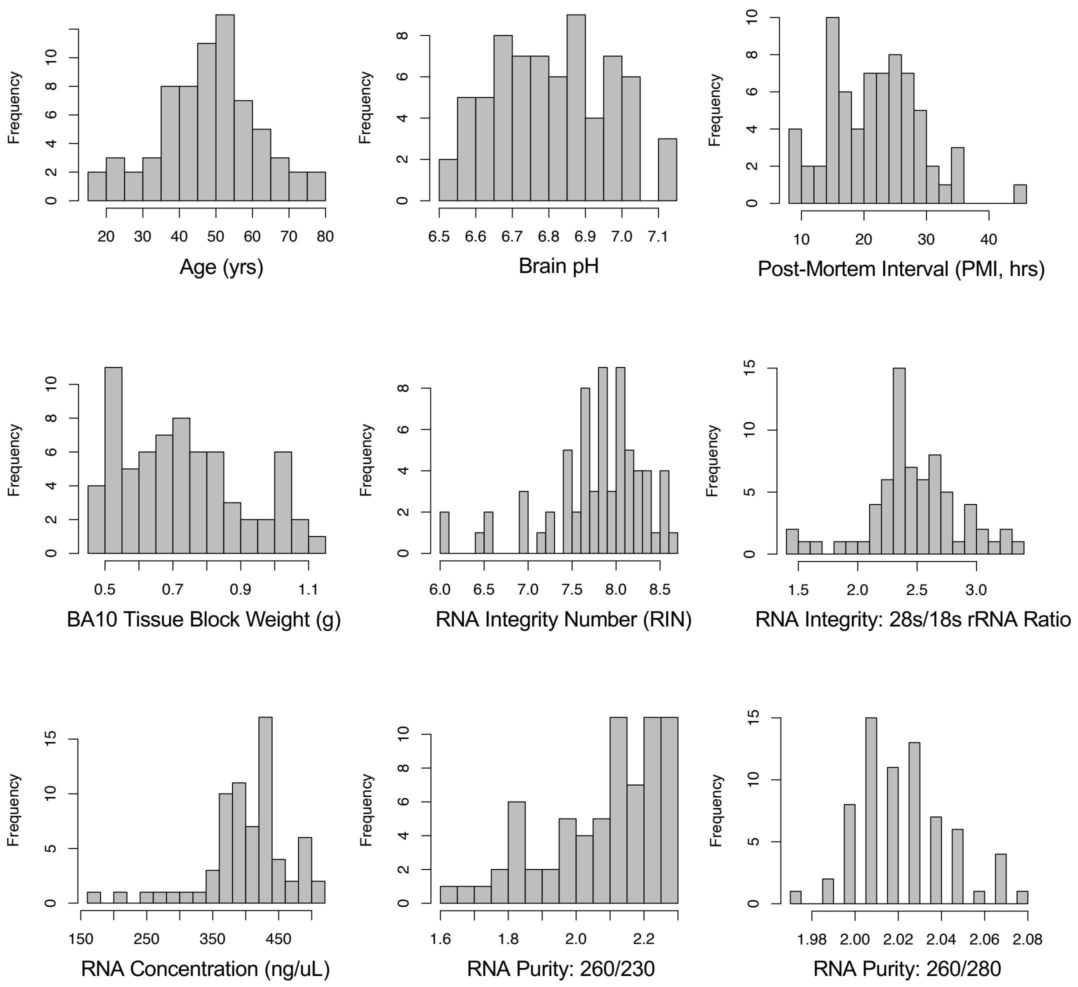

***Figure S 4. Histograms illustrating the distribution of each subject variable.***
*Histograms represent the distribution for each subject variable (demographic variables, tissue and RNA-quality metrics) following the exclusion of 3 subjects from the sample based on poor RNA quality metrics (two subjects had purified RNA with poor RNA integrity (RIN<5.5) and one subject had low RNA purity and concentration (average 260/230<1, average RNA concentration <120 ng/uL)). One subject was missing pH data and the RNA metrics from two subjects (batch 6v2) were lost prior to data analysis – these values were replaced with average values during data analysis, but were excluded during histogram creation.*

| Diagnosis | M | F |
|---|---|---|
| Control | 25 | 1 |
| BP | 15 | 6 |
| Schiz | 21 | 1 |

| | CTRL | BP | SCHIZ |
|---|---|---|---|
| African American | 1 | 0 | 0 |
| Asian | 1 | 0 | 0 |
| Caucasian | 22 | 21 | 20 |
| Latino | 2 | 0 | 1 |
| Pacific Islander | 0 | 0 | 1 |

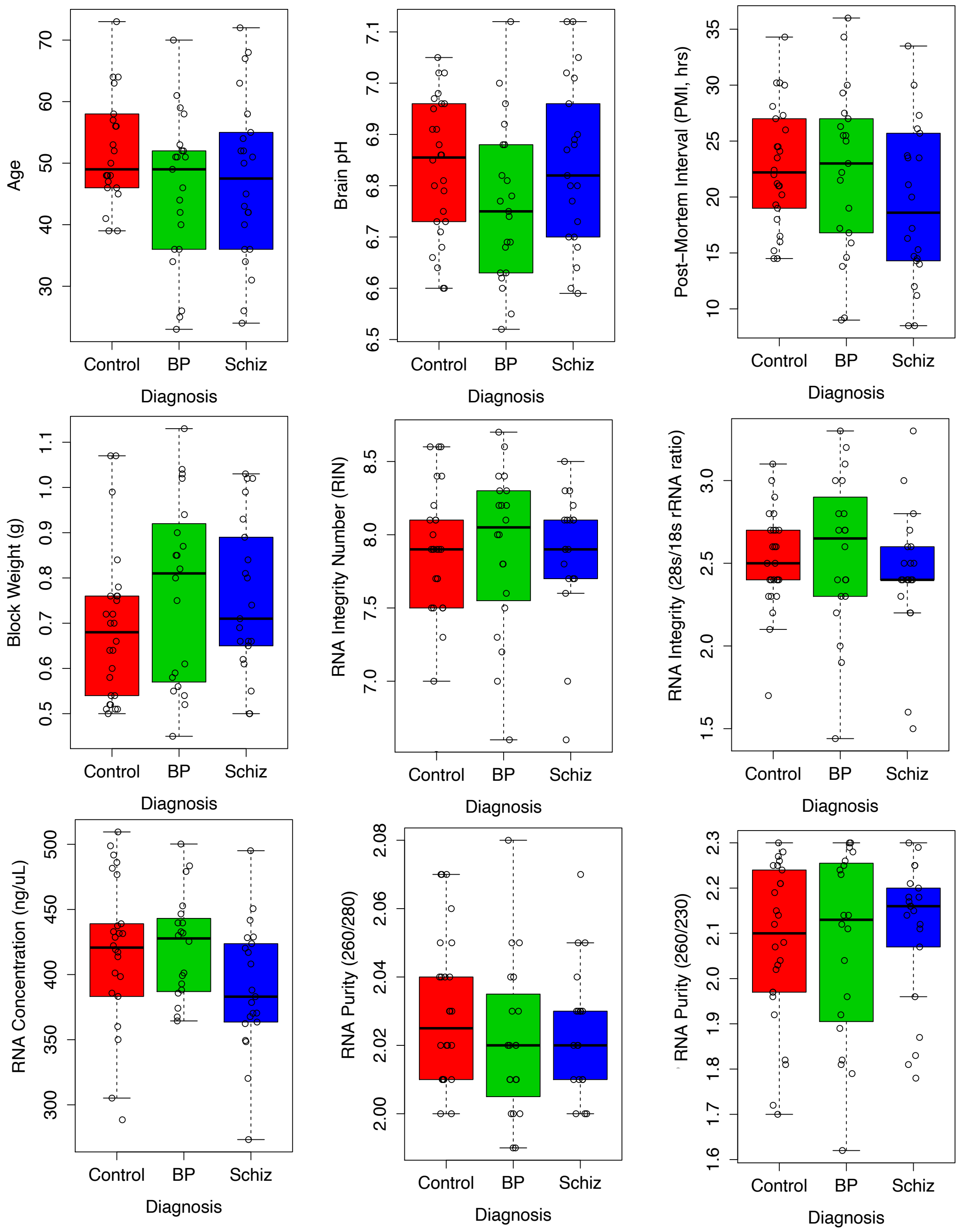

***Figure S 5. The relationship between each subject variable and diagnosis.***

*Following quality control, the final sample size included 69 subjects (CTRL: n=26, BPD: n=21, SCHIZ: n=22). Each boxplot represents the relationship between each subject variable (demographic variables, tissue and RNA-quality metrics) and diagnosis following the exclusion of 3 subjects from the sample based on poor RNA quality metrics (boxes=first quartile, median, and third quartile, whiskers = range and/or 1.5x the interquartile range if there are outlying data points). The full descriptive and inferential statistics for each relationship can be found in* ***Table S 4.*** *Gender (M=male, F=female) was the only subject variable that was significantly related to diagnosis, with the BP group including more female subjects (Fisher's Exact Test: p=0.0174). The diagnosis groups were otherwise balanced in regard to critical biological variables, including age (mean=48.3 yrs, range: 18-79 yrs), brain pH (mean=6.81, range=6.52-7.12), and post-mortem interval (PMI: mean=21.8, range=8.5-44.2 hrs), as well as tissue and RNA quality metrics, including the weight of the dissected BA10 blocks (mean=0.734 g, range=0.45-1.13), and the concentration (mean=403, range=179-510 ng/uL), purity (260/280: mean=2.02, range=1.97-2.08; 260/230: mean=2.08, range=1.62-2.3), and integrity of the purified RNA (RIN: mean=7.83, range=6-8.7, 28s/18s rRNA: mean=2.52, range=1.44-3.4) (p>0.10 for all variables).*

***General qPCR Methods:*** Procedures for cDNA synthesis and qPCR were performed by an experimenter blinded to the diagnosis associated with the samples. To produce cDNA for all samples, a reverse transcription reaction was performed within a single batch (plate) using the iScript Reverse Transcription Supermix kit (Bio-Rad REF#1708841). cDNA samples were then analyzed in duplicate via qPCR performed using the Applied Biosystems ViiA 7 real time PCR system and two sets of Taqman Gene Expression Custom Array qPCR cards (*described separately below*).

##### *GABA-GLU qPCR Experiment*

***Experimental Design:*** For this experiment, samples were analyzed using Taqman Human GABA Glutamate Gene Expression Array qPCR cards (ThermoFisher Scientific REF#4342259). Each qPCR card contained 4 samples, and each sample had 96 measurements, each representing a single gene (the full list of included genes can be found in **Table S 3)**. For each sample, 84 of these measurements were targeted genes and 12 measurements were "housekeeping" genes that were intended for usage as reference (18S Ribosomal RNA (18S), Hypoxanthine Phosphoribosyltransferase 1 (HPRT1), Glucuronidase Beta (GUSB), Actin Beta (ACTB), Beta-2-Microglobulin (B2M), Hydroxymethylbilane Synthase (HMBS), Importin 8 (IPO8), Phosphoglycerate Kinase 1 (PGK1), Ribosomal Protein Lateral Stalk Subunit P0 (RPLP0), TATA-Box Binding Protein (TBP), Transferrin Receptor (TFRC), and Ubiquitin C (UBC)). Each of the 72 subjects had two replicate samples that were run on adjacent cards (*e.g.,* Card 1 and Card 2), for 144 samples total.

***Quality Control and Normalization:*** A summary of quality control steps and metrics can be found in **Figure S 6** and **Figure S 7.** In general, the data were very high quality. Replicate Cq measurements were in strong agreement: there was a median correlation between replicate samples of R=0.995 (IQ range: 0.992-0.996). Two subjects had a single replicate sample for which the whole stem of wells did not amplify properly (>95% of measurements were missing values ("No Amp" or "Undetermined")). All remaining subjects had only 0-4 missing values across the Cq measurements for their two replicate samples (132 total, or less than 1% of the remaining dataset of 13,632 measurements (0.97%)). 99.6% of the measurements showed less than a 20% difference with their replicate and were deemed high quality. The 48 Cq measurements that were deemed low quality because they showed greater than 20% difference with their replicate were removed from the dataset.

| Subject-Level Quality Control | Both Datasets |
|---|---|
| Original subjects (n): | 72 subjects (n=27, BPD: n=21, SCHIZ: n=24) |
| Subjects removed during QC: | 2 subjects w/ low RIN (<3), 1 subject w/ low RNA purity/concentration |
| Subjects with missing metadata (imputed): | 2 subjects missing RNA metrics, 1 subject missing pH |
| Final subjects following QC (n): | 69 subjects (CTRL: n=26, BPD: n=21, SCHIZ: n=22) |

| Measurement-Level Quality Control: | GabaGlu Dataset | DA5HT Dataset |
|---|---|---|
| % that failed amplification (NA): | 2.30% | 0.69% |
| % w/ >20% difference with replicate: | 0.36% | 0.00% |
| % of measurements still bad following QC: | 0.13% | 0.00% |

| Sample-Level Quality Control: | | |
|---|---|---|
| Original samples (n): | 144 | 160 |
| Median replicate sample-sample correlation (R): | 0.995 | 0.996 |
| After removing subjects that failed QC (n): | 138 | 154 |
| Samples that failed amplification (>95% NA): | 2 | 0 |
| Extreme outlier samples identified by PCA: | 3 | 0 |
| Final samples (n): | 133 | 154 |

| Probe-Level Quality Control: | | |
|---|---|---|
| Original target genes (#): | 84 | 31 |
| Original reference genes (#): | 12 | 17 |
| Target genes w/ >20 bad measurements (#): | 2 (GABRR1, GRM6) | 2 (SLC18A1 and SLC6A3) |
| Reference genes w/ >20 bad measurements (#): | 1 (18S) | 0 |
| Final target genes following QC (#): | 82 | 29 |
| Final reference genes following QC (#): | 11 | 17 |

***Figure S 6. Quality Control Metrics Overview.***
*Abbreviations: GABA-GLU: The experiment that used the custom qPCR cards focused on GABA and glutamate related targets, DA5HT: The experiment that used the custom qPCR cards focused on dopamine and serotonin related targets, QC=quality control, n=total sample size, NA="Not Applicable", used to denote an empty measurement, PCA=principal components analysis.* ***Figure S 7*** *illustrates each of the quality control steps further.*

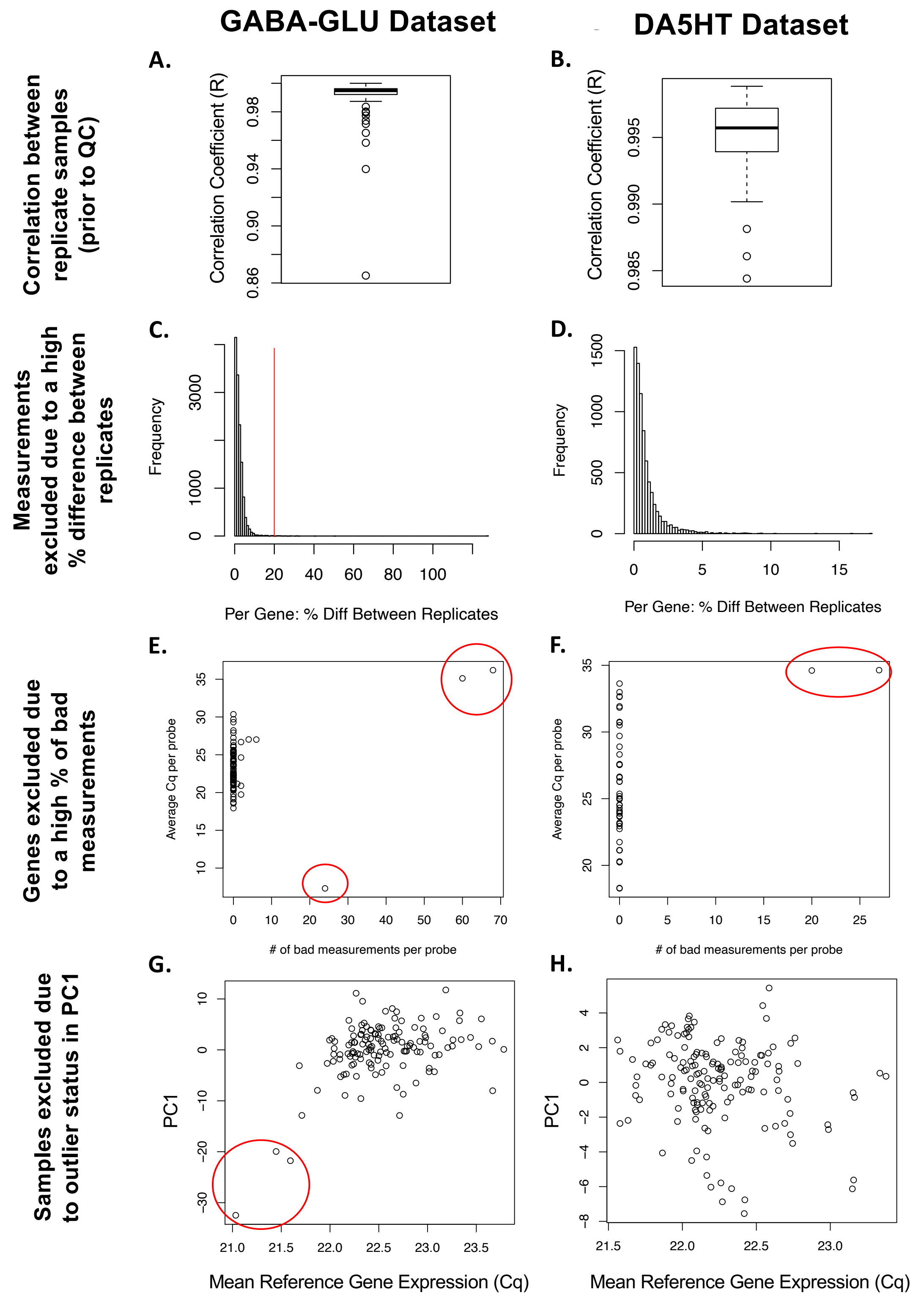

***Figure S 7. Quality control and data exclusion.***

***A-B)*** *A boxplot illustrating the correlation between replicate samples (R) in each dataset (box=first quartile, median, and third quartile, whiskers = range and/or 1.5x the interquartile* *range if there are outlying data points) prior to subject-level, sample-level, or probe-level quality control.* ***C-D)*** *A histogram illustrating the percentage difference between replicate measurements for each subject for each gene. Within the GABA-GLU dataset, replicate measurements that differed by >20% were considered low-quality and excluded from the dataset. Within the DA5HT dataset, no measurements met this criteria for exclusion.* ***E-F)*** *Following the exclusion of three subjects with low quality RNA metrics and two samples within the GABA-GLU dataset which had >95% of the measurements fail amplification, several genes still had an unusually high number (>15) of bad measurements (measurements that either failed amplification or that that differed by >20% from their replicate). All of these genes had either very high expression (average Cq<10) or very low expression (average Cq>34.5) and were excluded from the final dataset.* ***G-H)*** *Following basic quality control and normalization, principal components analysis was used to identify the principal components of variation in the normalized data (-ΔCq). The top principal component of variation in the GABA-GLU dataset (PC1) contained three outlier datapoints that also had particularly high average reference gene expression (Cq). These three samples did not resemble their replicates, suggesting technical artifact, and were excluded from the final dataset. The DA5HT dataset did not have similar outlier datapoints.*

The six samples from the three subjects that had poor RNA quality metrics (*discussed earlier*) were excluded from analysis and additional data cleaning was performed. In particular, three genes were found to have a high rate of missing and/or low quality (>20% difference between replicates) Cq measurements: one reference gene with unusually low Cq values on average (*i.e.,* particularly high expression, 18S: average Cq of 7.31, 24/136 or 18% of measurements NA), and two target genes with particularly high Cq values on average (*i.e.,* particularly low expression, average Cq>35: GABRR1: 68/136 or 50% of measurements NA, GRM6: 60/136 or 44% of measurements NA). The data for these genes was thrown out. All of the other 93 genes had only 0-6 missing and/or low quality Cq measurements total (mode of 0; **Table S 3**).

In general, our results indicated that the data could benefit from the traditional method of normalizing qPCR data using housekeeping gene expression as reference (-ΔCq, [9,10], **Figure S 8** and **Figure S 9).** There were notable differences in overall Cq (Cq z-score distribution) across samples that seemed to correlate with technical variation (qPCR card, RNA integrity, and RNA concentration). This pattern was mirrored in the data from just the reference (housekeeping) genes. The mean of Cq z-scores for target genes in the samples was tightly correlated with the mean of Cq z-scores for the reference genes (R=0.89) and the principal component of variation in the dataset (PC1), which accounted for more than 60% of the variation in the dataset, strongly correlated with average reference gene z-score (R=0.90).

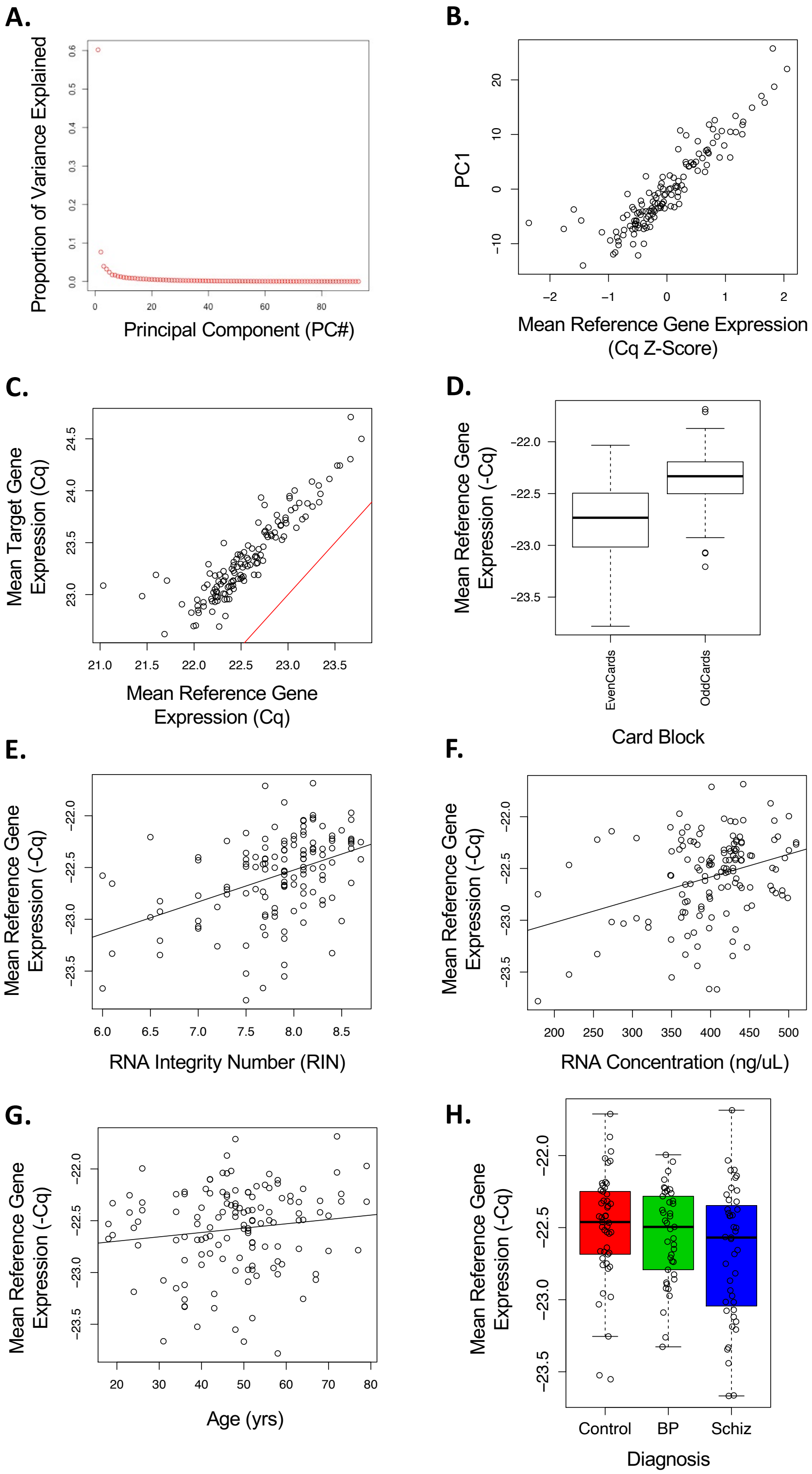

***Figure S 8. Reference gene expression in the GABA-GLU dataset.***
*Our results indicated that the data could benefit from the traditional method of normalizing qPCR data using housekeeping gene expression as reference (-ΔCq,* [9,10]*. **A)** A scree plot*

*illustrating the proportion of variance explained by each principal component of variation shows that the first principal component (PC1) accounted for more than 60% of the variation in the dataset prior to normalization (proportion>0.6). This analysis was performed using the Cq data (z-scores) following subject-level and probe-level quality control, but prior to eliminating the three outlier samples that were later identified using principal components analysis performed on the normalized (-ΔCq) data.* ***B)*** *PC1 strongly correlated with average reference gene (Cq) z-score (R=0.90).* ***C)*** *The mean of Cq z-scores for target genes in the samples tightly correlated with the mean of Cq z-scores for the reference genes (R=0.89),* ***D-F)*** *Following full quality control, mean reference gene expression (-Cq) continued to strongly correlate with technical variation in the data including* ***D)*** *qPCR card (not shown, Model=Eq.3, $\chi^2(35, n=133)=239.0453$, $p<2.2e-16$) or card "block" (even-numbered vs. odd-numbered cards, Model=Eq.5, $\chi^2(1, n=133)=98.8612$, $p<2.2e-16$),* ***E)*** *RNA integrity number (RIN) of the purified RNA (Model=Eq.3, $\chi^2(1, n=133)=42.0402$, $p=8.942e-11$),* ***F)*** *RNA concentration (ng/uL) of the purified RNA (Model=Eq.3, $\chi^2(1, n=133)=13.3676$, $p=0.000256$),* ***G)*** *Mean reference gene expression (-Cq) also correlated positively with subject age (yrs, Model=Eq.3, $\chi^2(1, n=133)=7.6693$, $p=0.005617$),* ***H)*** *Mean reference gene expression (-Cq) did not differ by diagnosis (Model=Eq.3, $\chi^2(2, n=133)=2.6325$, $p=0.2681$).*

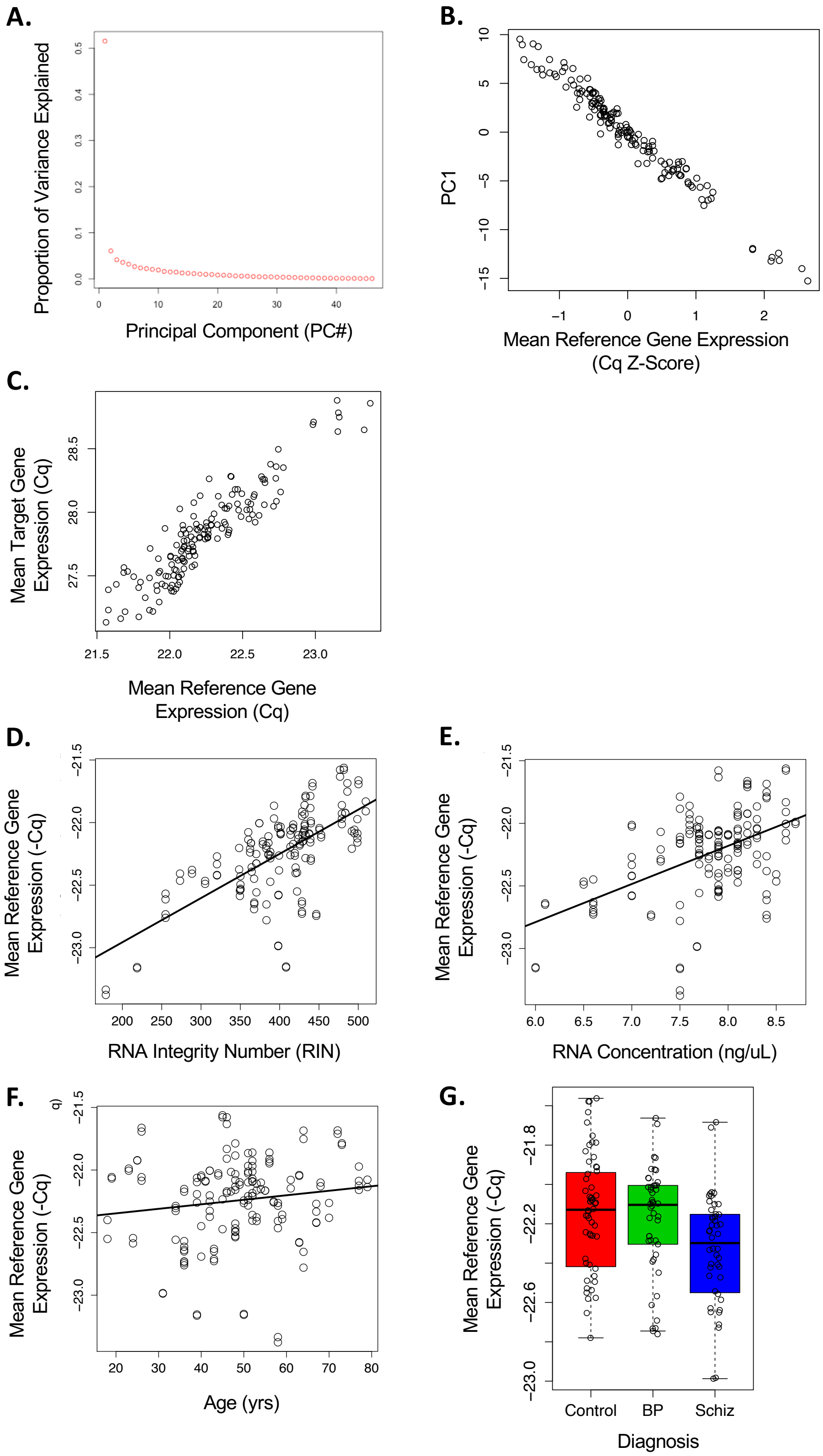

***Figure S 9. Reference gene expression in the DA5HT dataset.***
*Our results indicated that the data could benefit from the traditional method of normalizing qPCR data using housekeeping gene expression as reference (-ΔCq, [9,10]). **A)** A scree plot*

*illustrating the proportion of variance explained by each principal component of variation shows that the first principal component (PC1) accounted for more than 50% of the variation in the dataset prior to normalization (proportion>0.5). This analysis was performed using the Cq data (z-scores) following subject-level and probe-level quality control.* ***B)*** *PC1 strongly correlated with average reference gene (Cq) z-score (R=-0.98).* ***C)*** *The mean of Cq z-scores for target genes in the samples tightly correlated with the mean of Cq z-scores for the reference genes (R=0.94),* ***D-E)*** *Following full quality control, mean reference gene expression (-Cq) continued to strongly correlate with technical variation in the data including qPCR Card (not shown, Model=Eq.3, $\chi^2$(19, n=154)=240.3334, p<2.2e-16)* ***D)*** *RNA integrity number (RIN) of the purified RNA (Model=Eq.3, $\chi^2$(1, n=154)=27.0860, p=1.946e-07),* ***E)*** *RNA concentration (ng/uL) of the purified RNA (Model=Eq.3, $\chi^2$(1, n=154)=61.8920, p=3.628e-15),* ***F)*** *Mean reference gene expression (-Cq) also correlated positively with subject age (yrs, Model=Eq.3, $\chi^2$(1, n=154)=4.4269, p= 0.03538),* ***G)*** *Mean reference gene expression (-Cq) did not differ by diagnosis (Model=Eq.3, $\chi^2$(2, n=154)=1.0600, p=0.58859).*

To perform traditional qPCR normalization using reference (housekeeping) gene expression [9,10], we averaged the reference gene measurements (Cq) for each sample. Despite our data cleaning, 6 samples still had at least one missing or low quality measurement (NA) amongst the data from their remaining 11 reference genes. Therefore, before averaging the reference gene measurements (Cq) in each sample, these missing values were replaced with the average Cq for that gene from across all subjects. Then the average reference gene Cq measurements were subtracted from the Cq values for each target gene and inverted (multiplied by -1) to produce normalized target gene expression measurements (-ΔCq) for each sample, with each unit representing a doubling of expression (log2). For intuitive plotting, these units were then centered on the average -ΔCq from the control subjects to produce -ΔΔCq units (often called log(2) fold change). In our results we more formally defined log(2) fold change (-ΔΔCq) as the difference between the normalized gene expression measurements (-ΔCq) of case and control subjects as calculated within a regression model that controlled for important biological and technical co-variates (*see detailed discussion below)* [10].

Following normalization of the Cq values for the target genes using average "housekeeping" gene expression as the reference (-ΔCq), variability between samples was reduced. Principal components analysis was run again using the centered and scaled -ΔCq data (z-scores), with missing -ΔCq z-scores replaced with 0. PC1 now explained a smaller portion of the variance in the dataset (35%). Within PC1, there were three outlier samples. These three samples were all from control subjects. The replicates for these three outlier samples did not show similarly extreme values, and the three outlier samples also all had notably elevated average reference gene expression (mean Cq<21.6), suggesting technical artifact. These three outlier samples were excluded from the dataset, leaving a final sample size of *n*=133 samples representing 69 subjects. Principal components analysis was then run one more time on this final dataset and the results were used to guide the selection of biological and technical co-variates to include in the differential expression model (**Figure S 10*,*** *discussed below*).

**A.** Scree Plots: PCA for -ΔCq

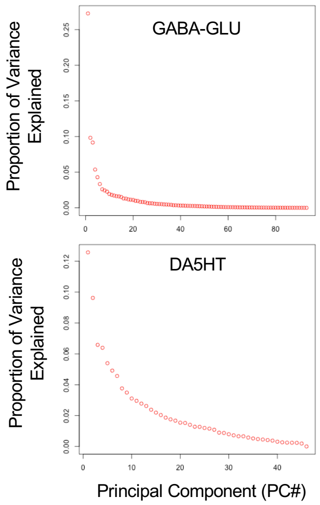

**B.** Potential Covariates vs. Principal Components of Variation: P-values

| | GABA-GLU Dataset | | | DA5HT Dataset | | |
|---|---|---|---|---|---|---|
| **Variable** | **PC1** | **PC2** | **PC3** | **PC1** | **PC2** | **PC3** |
| **Diagnosis** | 6.30E-01 | 3.05E-01 | 9.15E-01 | 1.88E-01 | 9.80E-02 | 3.59E-01 |
| **Gender** | 9.21E-01 | 9.47E-01 | 7.05E-01 | 8.02E-01 | 3.25E-01 | 9.20E-02 |
| **Age (yrs)** | **3.39E-04** | 1.26E-01 | **9.16E-06** | 6.28E-01 | 7.13E-02 | 1.55E-01 |
| **Brain pH** | **1.94E-02** | **3.56E-03** | **7.81E-03** | **4.29E-03** | 9.12E-01 | 5.67E-01 |
| **PMI (hrs)** | **4.84E-02** | 1.05E-01 | 1.95E-01 | **8.62E-03** | 6.83E-01 | 1.87E-01 |
| **Block Weight (g)** | 6.96E-01 | 7.77E-01 | 7.61E-01 | 9.89E-01 | 3.02E-01 | 7.02E-01 |
| **Concentration (ng/uL)** | 1.17E-01 | 1.41E-01 | 6.64E-01 | 6.18E-01 | 2.65E-01 | **3.41E-04** |
| **Integrity (RIN)** | 1.48E-01 | 5.34E-02 | 2.26E-01 | **6.01E-03** | **1.10E-03** | 3.88E-01 |
| **Integrity (28s/18s rRNA)** | **1.47E-02** | 7.22E-01 | **9.16E-03** | **1.34E-03** | 6.14E-01 | **4.69E-02** |
| **Purity (260/280)** | 3.83E-01 | 1.89E-01 | 1.01E-01 | 9.65E-01 | 5.19E-01 | **2.56E-02** |
| **Purity (260/230)** | 1.88E-01 | 7.67E-01 | 6.84E-01 | 4.75E-01 | 5.00E-01 | 7.71E-01 |
| **Dissecton/Extraction Group** | 8.43E-01 | **4.55E-02** | 7.93E-01 | 6.12E-01 | 8.62E-01 | **3.96E-03** |
| **qPCR Card** | **5.82E-31** | **3.67E-12** | **2.20E-54** | **2.78E-23** | **4.21E-26** | **1.19E-16** |
| **Card Block (Odd vs. Even)** | **4.36E-17** | **8.97E-03** | **2.08E-49** | | | |

**C.** Potential Covariates vs. Principal Components of Variation: Variables Surviving Automated Model Selection

| | Diagnosis | Gender | Age (yrs) | Brain pH | PMI (hrs) | Conc. (ng/uL) | Integrity (RIN) | Integrity (28s/18s rRNA) | Purity (260/280) | Purity (260/230) | qPCR Card | Card Block (Odd vs. Even) | Dissecton / Extraction Group |
|---|---|---|---|---|---|---|---|---|---|---|---|---|---|
| **GABAGLU - V1** | | | | | | | | | | | | | |
| PC1 | Default | Default | Y | Y | Y | Y | Y | N | N | N | NA | Y | N |
| PC2 | Default | Default | N | Y | Y | Y | Y | N | N | N | NA | Y | Y |
| PC3 | Default | Default | Y | N | N | Y | Y | Y | Y | N | NA | Y | N |
| **GABAGLU - V2** | | | | | | | | | | | | | |
| PC1 | Default | Default | Y | Y | Y | Y | Y | N | N | Y | Y | NA | NA |
| PC2 | Default | Default | Y | Y | Y | N | Y | N | Y | Y | Y | NA | NA |
| PC3 | Default | Default | Y | N | N | Y | Y | Y | Y | N | Y | NA | NA |
| **DA5HT - V1** | | | | | | | | | | | | | |
| PC1 | Default | Default | N | Y | Y | N | Y | Y | N | N | Y | NA | N |
| PC2 | Default | Default | N | N | N | N | Y | N | N | N | Y | NA | Y |
| PC3 | Default | Default | Y | Y | Y | N | N | N | N | N | Y | NA | Y |
| **DA5HT - V2** | | | | | | | | | | | | | |
| PC1 | Default | Default | N | Y | Y | N | Y | Y | N | N | Y | NA | NA |
| PC2 | Default | Default | N | N | N | N | Y | N | N | N | Y | NA | NA |
| PC3 | Default | Default | Y | N | Y | Y | N | N | N | N | Y | NA | NA |

***Figure S 10. The biological and technical variables that had the strongest relationships with the top three principal components of variation (PC1-3) were chosen as co-variates in our final model.***

*Principal components analysis was run on the final, normalized dataset following all quality control (-ΔCq) in both datasets.* ***A)*** *Scree plots illustrating the proportion of variance explained by each principal component of variation in the two datasets shows that PC1 now accounts for a much lower proportion of the variance following full quality control and normalization (vs.* ***Figure S 8A*** *and* ***Figure S 9A****),* ***B)*** *Nominal p-values for the relationship between each potential co-variate and each of the top principal components of variation (PC1-3) in the two datasets, as derived from a simple multilevel model containing only the variable of interest and subject ID as a random effect variable (Eq.1),* ***C)*** *Variables that survived two versions of an automated model selection procedure for each of the top principal components of variation (PC1-3). The automated model selection procedure used the step() function (lmertest package v.3.1-3, [11]), which conducted backward elimination (threshold alpha=0.1) from a full multi-level model containing all potentially relevant biological and technical co-variates in addition to the required terms of Diagnosis and Gender (Eq.2). We ran two versions of the automated model selection procedure because there were two potential batch-related technical co-variates (Dissection/extraction Group and qPCR Card) that required a large number of degrees of freedom in the model and were highly multi-collinear (especially within the GABAGLU dataset, which only had 4 samples per qPCR card). Therefore, during our automated model selection* *procedure we ran separate models including batch as defined as either **V1)** the two variables of Dissection/extraction batch and the simplified variable of CardBlock (odd vs. even-numbered cards, for GABAGLU) or Dissection/extraction batch and Card (for DA5HT), **V2**) qPCR Card.*

##### *DA-5HT qPCR Experiment*

***Experimental Design:*** For this experiment, samples were analyzed using Taqman Dopamine Serotonin Gene Expression Array qPCR Cards (ThermoFisher Scientific REF#4342253). Each qPCR card contained 8 samples, and each sample had 48 measurements, each representing a single gene (the full list of included genes can be found in **Table S 3**). For each sample, 31 of these measurements were target genes and 17 measurements were “housekeeping” genes that were intended for usage as reference (HPRT1, GUSB, B2M, IPO8, TFRC, Glyceraldehyde 3-phosphate dehydrogenase (GAPDH), Tyrosine 3-Monooxygenase/Tryptophan 5-Monooxygenase Activation Protein Zeta (YWHAZ), Peptidylprolyl Isomerase A (PPIA), RNA Polymerase II Subunit A (POLR2A), CASC3 Exon Junction Complex Subunit (CASC3), Growth Arrest And DNA Damage Inducible Alpha (GADD45A), Pumilio RNA Binding Family Member 1 (PUM1), Proteasome 26S Subunit, ATPase 4 (PSMC4), ER Membrane Protein Complex Subunit 7 (EMC7), Glucose-6-Phosphate Isomerase (GPI), RAB7A, Member RAS Oncogene Family (RAB7A), and Receptor Accessory Protein 5 (REEP5)). Five of these reference genes (HPRT1, GUSB, B2M, IPO8, and TFRC) had been previously included on the GABA-GLU qPCR cards. Every subject had at least two replicate samples, 8 subjects were run in quadruplicate.

***Quality Control and Normalization:*** Similar to the GABA-GLU experiment, the data were again of very high quality (**Figure S 6, Figure S 7**). Replicate Cq measurements were in strong agreement: there was a median correlation between replicate samples of R=0.9957 (IQ range: 0.9940-0.9972). There were fewer missing Cq measurements in the DA-5HT dataset than in the GABA-GLU dataset (53/7680 Cq values or 0.69%) and no samples had high numbers of missing measurements (range: 0-3 NA values). All measurements showed less than a 20% difference with their replicate.

The samples from the three subjects that had poor RNA quality metrics (*discussed earlier*) were excluded from analysis and additional data cleaning was performed. In particular, two target genes with particularly high Cq values on average (*i.e.,* particularly low expression, average Cq>34.6) were found to have a high rate of missing and/or low quality (“NA”) Cq values (SLC18A1: 27/154 or 18% of measurements NA, SLC6A3: 20/154 or 13% of measurements NA). The data for these genes was thrown out. Following this cleaning, there was only one other gene with a single NA measurement (**Table S 3**). There was also a strong correlation between the Cq values (averaged by subject) for the five genes represented on both the GABA-GLU and DA-5HT cards (HPRT1: R=0.69, GUSB: R=0.62, B2M: R=0.89, IPO8: R=0.66, TFRC: R=0.84, **Figure S 11**), indicating that the sample-level technical variation in the datasets represented a smaller percentage of the variation than the subject-level variation in the datasets, even prior to normalization.

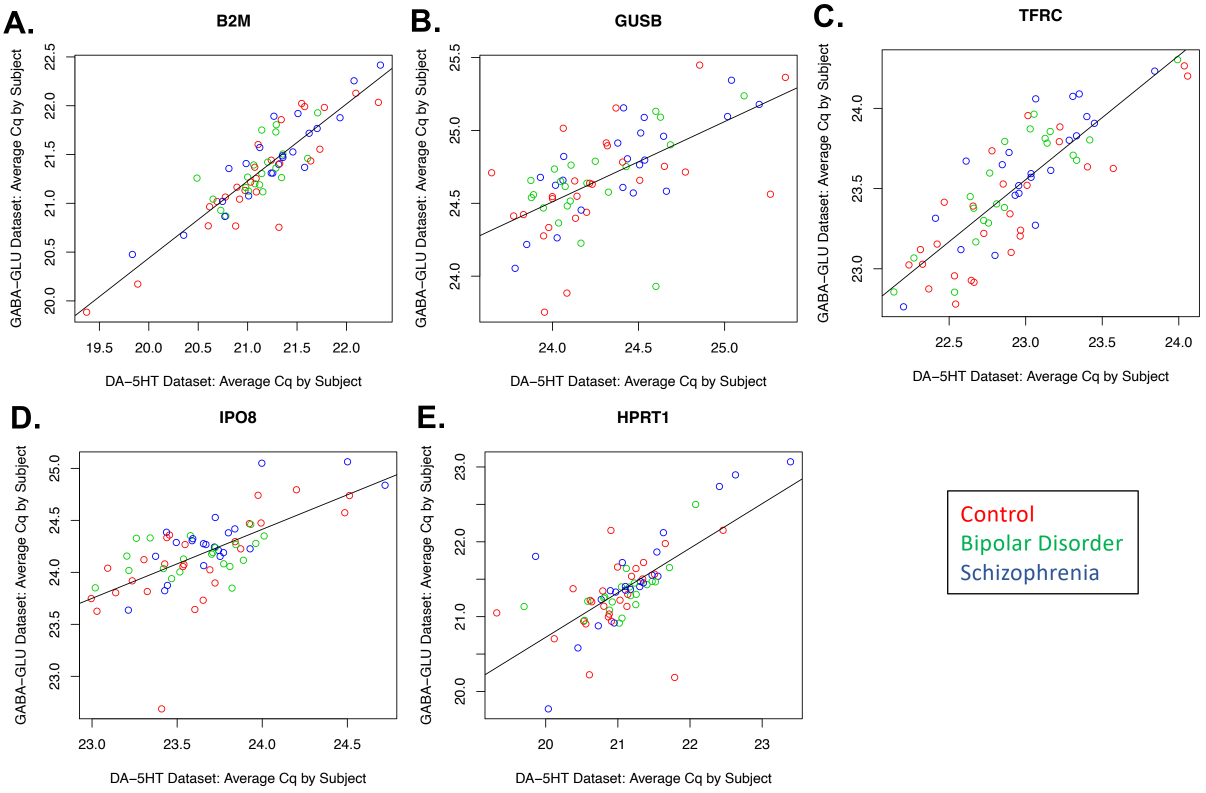

***Figure S 11. There was a strong correlation between the Cq values for the five reference genes represented on both the GABA-GLU and DA-5HT cards, indicating that both datasets contain high quality data prior to quality control and normalization.***
*To assess replication across experiments the Cq values were averaged by subject within each experiment (scatterplots: y-axis=average Cq per subject in the GABA-GLU dataset, x-axis=average Cq per subject in the DA-5HT dataset). Color indicates the diagnosis for each subject (red=CTRL, green=BP, blue=SCHIZ).* ***A)*** *Beta-2-Microglobulin (B2M, R=0.89),* ***B)*** *Glucuronidase Beta (GUSB, R=0.62),* ***C)*** *Transferrin Receptor (TFRC, R=0.84),* ***D)*** *Importin 8 (IPO8, R=0.66),* ***E)*** *Hypoxanthine Phosphoribosyltransferase 1 (HPRT1, R=0.69).*

Similar to the data from the GABA-GLU cards, in general our results still indicated that the data could benefit from the traditional method of normalizing qPCR data using housekeeping gene expression as reference (-ΔCq, [9,10], **Figure S 8** and **Figure S 9**). There were notable differences in overall Cq (Cq z-score distribution) across samples that seemed to at least partially correlate with technical variables (qPCR card, RNA integrity, and RNA concentration). This pattern was mirrored in the data from just the reference (housekeeping) genes. In general, the mean Cq z-scores for target genes in the samples was tightly correlated with the mean Cq z-scores for the reference genes (R=0.937) and the principal component of variation in the dataset (PC1), which accounted for more than 50% of the variation in the dataset, strongly correlated with average reference gene z-score (R= -0.984).

Similar to the GABA-GLU dataset, we performed traditional qPCR normalization using reference gene expression [9,10] and then ran principal components analysis again on centered and scaled -ΔCq data (z-scores), with missing -ΔCq z-scores replaced with 0. PC1 now explained a smaller portion of the variance (12%, **Figure S 10**). No samples had extreme PC1 values and no outlier samples were removed, leaving a final sample size of $n$=154 samples representing 69 subjects.

##### *Differential Expression Analysis*

***Model Selection:*** Our previous experience analyzing gene expression in human post-mortem brains has repeatedly demonstrated that the effects of psychiatric diagnosis in the brain are often masked by other biological (age, agonal factor, brain pH, post-mortem interval) and technical variables (dissection, batch effects, RNA integrity, RNA concentration) that add confounding variability or large magnitude noise within our gene expression measurements [1–3,12,13]. Within the current qPCR dataset, there was only one variable that was correlated with diagnosis (gender, **Figure S 5**), but there was variation across quite a few impactful demographic and technical variables that could influence our results (**Figure S 4)**.

To identify the variables that were most likely to introduce noise in our results, we first examined the relationship between each of these potential co-variates and the top principal components of variation (PC1-PC3) identified within the normalized (-ΔCq) final GABA-GLU and DA5HT datasets (**Figure S 10**). Since each subject was represented by 1-4 samples, subject ID was included as a random effect variable in all models (*Eq.1*). These multi-level models were then evaluated using function lmer (*lme4* package v1.1-13 [14]), with estimates optimized using log-likelihood criteria, and p-values assigned using a Type III Wald test comparing the full and reduced model for each variable using the *anova()* function (*car* package v2.1-5 [15]).

*Equation 1:* $PC\# \sim \beta_0+\beta_x(Variable),\ random=\sim 1|ID$

For each of the top principal components of variation (PC1-PC3), we also determined which variables survived an automated model selection procedure using the *step()* function (*lmertest* package v.3.1-3, [11]), which conducted backward elimination (threshold alpha=0.1) from a full multi-level model containing all potentially relevant biological and technical co-variates in addition to the required terms of Diagnosis and Gender (*Eq.2*). Two batch-related technical co-variates had been implicated within our initial analysis comparing PC1-3 to each of the potential co-variates individually (dissection/extraction batch and qPCR card) but required a large number of degrees of freedom in the model and were highly multi-collinear (especially within the GABAGLU dataset, which only had 4 samples per qPCR card). Attempts to reduce the dimensions for Card using hierarchical clustering indicated that part of the effect of Card within the GABA-GLU dataset was caused by a pattern of differential expression across odd and even-numbered cards ("CardBlock"). Therefore, during our automated model selection procedure we ran separate models including batch as defined as either 1) qPCR Card, or 2) the two variables of Dissection/extraction batch and CardBlock (for GABAGLU) or Dissection/extraction batch and Card (for DA5HT).

*Equation 2:* $PC\# \sim \beta_0 + \beta_{1\text{-}2}(Diagnosis) + \beta_3(Gender) + \beta_4(Age) + \beta_5(pH) + \beta_6(PMI) + \beta_7(Concentration) + \beta_8(Integrity\_RIN) + \beta_9(Integrity\_28S/18S\_rRNA) + \beta_{10}(Purity\_260/280) + \beta_{11}(Purity\_260/230) + \beta_X(Batch^*),\ random=\sim 1|ID$

The results from these automated model selection analyses indicated that three biological co-variates (Age, pH, and PMI), two traditional RNA metrics (RNA Concentration and Integrity (RIN)), and batch (defined in both manners) might be particularly important sources of noise in the dataset (**Figure S 10**). Out of the batch-related variables, qPCR card was particularly strongly associated with all three of the top principal components of variation in both datasets, whereas models including dissection group often showed only borderline significant improvement when compared using *anova(),* and worse performance when evaluated using other metrics *(AIC, BIC)*, therefore moving forward we decided to focus on models containing Card instead of Dissection/extraction batch. The three less-traditional RNA metrics (Integrity (28s/18s) and Purity (260/280 or 260/230)) were occasionally related to the top principal components of variation (**Figure S 10**), and therefore were considered in later exploratory analyses.

***Differential Expression Analysis:*** We used the results from the principal components analysis as a guide for determining the most important co-variates to include in the differential expression analysis. The effect of diagnosis (log2 fold change, or -ΔΔCq) was evaluated using the normalized gene expression measurements (-ΔCq) for each target gene within a multilevel (mixed effects) model (*Equation 3*), with subject ID included as a random effect variable.

*Equation 3:* $y \sim \beta_0 + \beta_{1\text{-}2}(Diagnosis) + \beta_3(Gender) + \beta_4(Age) + \beta_5(pH) + \beta_6(PMI) + \beta_7(Concentration) + \beta_8(Integrity\_RIN) + \beta_x(Card^*)$*, random=~1|ID*

As before, this multi-level model was evaluated using function *lmer* (*lme4* package v1.1-13 [14]), with estimates optimized using log-likelihood criteria, and p-values assigned using a Type III Wald test comparing the full and reduced model for each variable using the *Anova* function (*car* package v2.1-5 [15]). To be conservative, the p-values were corrected for false discovery rate ("FDR") following concatenation of the results from the target genes included in both the GABA-GLU and DA-5HT experiments (*discussed below*) using the Benjamini-Hochberg method within the function *mt.rawp2adjp* (package *multtest* v2.32.0 [16]).

As a post-hoc analysis to characterize the magnitude and direction of effect of each diagnosis on gene expression (-ΔCq), each multilevel model was re-evaluated using the function *lme* (package *nlme* v3.1-131 [17]), with estimates similarly optimized using maximum log-likelihood, and summarized to produce individual coefficients, approximate standard errors and respective p-values for each level of the fixed effects variables using the function *summary().* We double-checked that average reference gene expression prior to normalization (-Cq) was not associated with diagnosis using the same statistical methods ($p>0.17$) to ensure that diagnosis effects were not being artificially introduced into the results by normalization procedures (**Figure S 8, Figure S 9**).

Finally, out of concern that the variable of Card might produce overfitting, the results from our chosen model were compared to the results from two other models with reduced complexity (*Equations 4 & 5*):

*Equation 4:* $y \sim \beta_0 + \beta_{1\text{-}2}(Diagnosis)$*, random=~1|ID*

*Equation 5:* $y \sim \beta_0 + \beta_{1\text{-}2}(Diagnosis) + \beta_3(Gender) + \beta_4(Age) + \beta_5(pH) + \beta_6(PMI) + \beta_7(Concentration) + \beta_8(Integrity\_RIN) + \beta_9(CardBlock, \text{when applicable})$*, random=~1|ID*

Later, other exploratory analyses were similarly performed to determine the sensitivity of our results to model specification, including the inclusion of less-traditional RNA metrics (***Figure S 12***). Exploratory analyses to determine the sensitivity of our results to model specification revealed remarkable stability in effect size estimation. Notably, the effect of diagnosis on HTR2B expression was so large that it survived false discovery rate correction even when using a highly-reduced model that only included diagnosis (*i.e.,* no control for other sources of noise and confounding variability in the dataset).

| | | Beta / Log2FC: | | | | | | | | | | Nominal P-value | | | | | | | | | |
|---|---|---|---|---|---|---|---|---|---|---|---|---|---|---|---|---|---|---|---|---|---|
| | GeneSymbol | Eq3_BiolCovariatesRINConc_Card | Eq4_JustDiagnosis | Eq5_BiolCovariatesRINConc_CardBlock | Eq6_DiagnosisRNAMetrics_CardBlock | Eq7_DiagnosisRNAMetrics_Card | Eq8_BiolCovariates_CardBlock2 | Eq9_BiolCovariates_Card | Eq10_BiolCovariatesRNAMetrics_CardBloc | Log2FC_MIN | Log2FC_MAX | Eq3_BiolCovariatesRINConc_Card | Eq4_JustDiagnosis | Eq5_BiolCovariatesRINConc_CardBlock | Eq6_DiagnosisRNAMetrics_CardBlock | Eq7_DiagnosisRNAMetrics_Card | Eq8_BiolCovariates_CardBlock2 | Eq9_BiolCovariates_Card | Eq10_BiolCovariatesRNAMetrics_CardBloc | PVAL_MIN | PVAL_MAX |
| BP: | | | | | | | | | | | | | | | | | | | | | |
| | ABAT | 0.16 | 0.16 | 0.18 | 0.16 | 0.15 | 0.18 | 0.16 | 0.16 | 0.15 | 0.18 | 0.0055 | 0.0065 | 0.0065 | 0.0056 | 0.0023 | 0.0059 | 0.0044 | 0.0101 | 0.0023 | 0.0101 |
| | SST | -0.31 | -0.41 | -0.33 | -0.44 | -0.35 | -0.32 | -0.31 | -0.36 | -0.44 | -0.31 | 0.0935 | 0.0329 | 0.0596 | 0.0162 | 0.0334 | 0.0682 | 0.1040 | 0.0428 | 0.0162 | 0.1040 |
| | GPHN | 0.16 | 0.09 | 0.12 | 0.10 | 0.14 | 0.12 | 0.16 | 0.12 | 0.09 | 0.16 | 0.0026 | 0.1181 | 0.0424 | 0.0994 | 0.0031 | 0.0459 | 0.0037 | 0.0436 | 0.0026 | 0.1181 |
| | MAPK1 | 0.14 | 0.16 | 0.15 | 0.17 | 0.14 | 0.15 | 0.14 | 0.15 | 0.14 | 0.17 | 0.0113 | 0.0021 | 0.0042 | 0.0016 | 0.0103 | 0.0048 | 0.0109 | 0.0041 | 0.0016 | 0.0113 |
| | DRD4 | -0.47 | -0.41 | -0.51 | -0.41 | -0.42 | -0.56 | -0.52 | -0.52 | -0.56 | -0.41 | 0.0051 | 0.0407 | 0.0085 | 0.0267 | 0.0067 | 0.0089 | 0.0089 | 0.0059 | 0.0051 | 0.0407 |
| | HTR2B | -0.68 | -0.83 | -0.61 | -0.85 | -0.90 | -0.60 | -0.68 | -0.65 | -0.90 | -0.60 | 0.0270 | 0.0029 | 0.0336 | 0.0017 | 0.0021 | 0.0351 | 0.0266 | 0.0226 | 0.0017 | 0.0351 |
| SCHIZ: | | | | | | | | | | | | | | | | | | | | | |
| | ABAT | 0.08 | 0.10 | 0.12 | 0.11 | 0.08 | 0.11 | 0.08 | 0.12 | 0.08 | 0.12 | 0.0993 | 0.0881 | 0.0363 | 0.0583 | 0.0892 | 0.0478 | 0.1194 | 0.0387 | 0.0363 | 0.1194 |
| | SST | -0.47 | -0.39 | -0.43 | -0.38 | -0.38 | -0.47 | -0.52 | -0.43 | -0.52 | -0.38 | 0.0069 | 0.0385 | 0.0099 | 0.0384 | 0.0185 | 0.0044 | 0.0032 | 0.0100 | 0.0032 | 0.0385 |
| | GPHN | 0.10 | 0.05 | 0.10 | 0.07 | 0.10 | 0.08 | 0.08 | 0.10 | 0.05 | 0.10 | 0.0295 | 0.3521 | 0.0825 | 0.1995 | 0.0324 | 0.1776 | 0.1112 | 0.0617 | 0.0295 | 0.3521 |
| | MAPK1 | 0.09 | 0.07 | 0.08 | 0.09 | 0.10 | 0.07 | 0.07 | 0.09 | 0.07 | 0.10 | 0.0730 | 0.1622 | 0.0780 | 0.0780 | 0.0646 | 0.1443 | 0.1265 | 0.0655 | 0.0646 | 0.1622 |
| | DRD4 | -0.35 | -0.35 | -0.35 | -0.35 | -0.36 | -0.38 | -0.36 | -0.35 | -0.38 | -0.35 | 0.0244 | 0.0797 | 0.0483 | 0.0573 | 0.0220 | 0.0517 | 0.0435 | 0.0433 | 0.0220 | 0.0797 |
| | HTR2B | -0.98 | -1.00 | -0.97 | -1.00 | -1.01 | -0.99 | -1.00 | -0.97 | -1.01 | -0.97 | 0.0009 | 0.0004 | 0.0005 | 0.0003 | 0.0006 | 0.0003 | 0.0005 | 0.0004 | 0.0003 | 0.0009 |

***Figure S 12. The results from an analysis exploring the sensitivity of the estimation of the effect of diagnosis on gene expression to model specification: An illustration of the results for just the top diagnosis-related genes.***

*The statistical output for each diagnosis (SCHIZ or BP) for the six genes that showed a significant relationship with diagnosis using our specified model (FDR<0.10) when using other model specifications. The specifications for each of the statistical models that were compared as well as the full results for all other genes can be found in* ***Table S 6.*** *The effect of each diagnosis (Beta, Log(2) Fold Change, or -ΔΔCq) on the normalized gene expression measurements (-ΔCq) for each gene as derived from each model is color-coded so that an increase in expression is illustrated in pink and a decrease is illustrated in blue. Log2FC_MIN and Log2FC_MAX provide the minimum and maximum Beta (Log(2) Fold Change) across all models, respectively. The nominal p-values associated with those effects are color-coded so that green signals a lower ("more significant") p-value and orange signals a higher p-value. PVAL_MIN and PVAL_MAX provide the minimum and maximum nominal p-value across all models, respectively. Genes that have diagnosis effects that are sensitive to model specification (e.g., GPHN) should be interpreted carefully. Notably, the effect of diagnosis on HTR2B expression was so large that it survived false discovery rate correction even when using a highly-reduced model that only included diagnosis (i.e., no control for other sources of noise and confounding variability in the dataset).*

***Post-hoc Exploratory Analysis:*** One strength of our dataset was the richness of the subject metadata available to accompany the results due to in-depth psychological autopsy (**Appendix 1).** We summarized this metadata as a database of 46 binary variables (True/False, **Figure S 2**) overviewing 1) diagnosis-related symptoms and related behaviors, 2) medication, 3) exposure to alcohol or drugs of abuse (*discussed earlier)*. We also created three ordinal variables based on documented presence (coded as 1), absence (coded as -1), or lack of information (coded as 0) as derived from toxicology reports for Antipsychotics, Antidepressants, and Opioids.

To explore the relationship between each of these 49 exploratory variables and the expression of our top diagnosis-related genes (n=20, **Figure 3**), we added each variable of interest to our previous qPCR differential expression model:

*Equation 6:* $y \sim \beta_0 + \beta_{1\text{-}2}(Diagnosis) + \beta_3(\underline{Exploratory\ Variable}) + \beta_4(Gender) + \beta_5(Age) + \beta_6(pH) + \beta_7(PMI) + \beta_8(Concentration) + \beta_9(Integrity\_RIN) + \beta_x(Card^*)$*, random=~1|ID*

The output for all 49 variables from all genes was then concatenated, and a false discovery rate correction was run on the p-values for the exploratory variables using methods discussed above. To further reduce probability of false discovery, the putative relationships between target gene expression and the exploratory variables were examined visually for small subgroup sample size (n<6) and substance-related results were cross-referenced with previous evidence indicating an association between the substance and expression of the target gene (*substance abuse:* human post-mortem studies [18–20]*, antipsychotics:* primate prefrontal cortex [21], Drug Gene Budger database: https://maayanlab.cloud/DGB/, [22], *described below*).

#### Meta-Analysis of BA10 Microarray Datasets:

We compared our results to previously-published transcriptional profiling studies examining gene expression in BA10 in relationship to Schizophrenia and Bipolar Disorder [23–26]. Then, to increase consistency, we re-annotated and re-analyzed the data from the two BA10 microarray studies that had publicly released their data (Maycox et al. 2009; Iwamoto et al. 2004). Of these studies, two had available data [23,24], which allowed us to perform a more detailed comparison by re-annotating, re-analyzing, and then meta-analyzing their results.

##### *Iwamoto et al. 2004:*

***Study Overview:*** Iwamoto et al. [23] used microarray (Affymetrix HG-U95A) to characterize gene expression in post-mortem BA10 tissue from subjects with Schizophrenia, Bipolar Disorder, and Major Depressive Disorder, and matched controls. Donor recruitment and the collection of BA10 tissue was conducted by the Stanley Foundation ("Neuropathology Collection"). The dissection method emphasized grey matter, and was likely to include pia mater on the cortical surface but a minimal amount of subcortical white matter (Dr. Maree Webster, *personal communication*). The subjects had been diagnosed using DSM-IV criteria, and their medical history was screened to rule out neurological illness. Most subjects had experienced sudden death, but there were a few subjects on a respirator (n=4) or with possible anoxia (n=3). The experimental design was well-balanced across diagnosis groups in regards to many variables of interest, including age, gender, hypoxia (pH), post-mortem interval, rate of death, lifetime alcohol, lifetime drugs, and smoking. In general, the brain pH was lower (interquartile range between 6.0-6.5) indicating greater hypoxia prior to death, the post-mortem interval was similar (interquartile range between 18-38 hrs), and the age was younger (interquartile range between 30-55 yrs) for the samples in this dataset in comparison to our qPCR experiment.

***Preprocessing:*** We imported the microarray .CEL files and metadata from GEO (https://www.ncbi.nlm.nih.gov/geo/, Accession# GSE12654) using the "GEOquery" R package (v.2.56.0, [27]). We summarized these data using Robust Multiarray Average (RMA [28]) using a custom up-to-date chip definition file (.cdf) to define probe-to-transcript correspondence ("hgu95av2hsentrezgcdf_25.0.0" downloaded from http://brainarray.mbni.med.umich.edu/Brainarray/Database/CustomCDF/CDF_download.asp [29]).

This process included background subtraction, log(2)-transformation, and quantile normalization. Gene Symbol annotation for probeset Entrez gene ids were provided by the R package *org.Hs.eg.db.* (v.3.11.4). To control for technical variation, the sample processing batches were estimated using the microarray chip scan dates extracted from the .CEL files (using the function *protocolData* in the GEOquery package [27]). RNA degradation was estimated computationally using the 3′/5′ intensity ratios for the probes for each gene (R package AffyRNADegradation, v.1.34.0, [30]).

***Quality Control:*** The original study had a sample size of *n*=60 (n=15/diagnosis), but 10 samples were removed during the quality control that accompanied the original analysis, leaving a final sample size of *n*=50 (CTRL: n=15, BPD: n=11, SCHIZ: n=13, MDD=11, [23]). Only data from these 50 samples was released on Gene Expression Omnibus (GEO). In these remaining samples, the overall median sample-sample correlation was high (median=0.988), implying good technical consistency. There was no evidence of sample mismatch or sample mixing as indicated by agreement between subject gender and the strongly bimodal distributions for sex chromosome gene expression (XIST, RPS4Y1, DDX3Y, [31]).

***Differential Expression Analysis:*** All available demographic and technical variables of interest were confirmed to be balanced across diagnosis groups ($p$>0.13, Age, pH, PMI, RNADegradation, Gender, RateOfDeath, SmokingAtTOD, LifetimeDrugs, LifetimeAlcohol) using either a chi-square test (categorical vs. categorical) or ANOVA (continuous vs. categorical). There were too many processing batches as estimated using scan date to be considered as a meaningful covariate (11 scan dates).

To identify sources of large-scale noise in the data, each of the demographic and technical variables was examined in relationship to the top principal components of variation using either ANOVA or linear regression. PC1 was related to RateOfDeath (p=0.0153) and RNADegradation (p=4.77e-06). PC2 was related to BrainPH (p=0.000516), RateOfDeath (p=0.0412), and SmokingAtTOD (p=0.0223). PC3 was related to pH (p=4.95e-06) and PMI (p=0.0469).

Based on these results, we chose to examine results from three models:

Basic Model (“Model 1”)
*Equation 7:* $y \sim \beta_0 + \beta_{1\text{-}3}(Diagnosis)$

Model with largest implicated sources of noise (“Model 2”):
*Equation 8:*
$\beta_0 + \beta_{1\text{-}3}(Diagnosis) + \beta_4(pH) + \beta_5(RNADegradation) + \beta_{6\text{-}7}(RateOfDeath)$

Model with implicated sources of noise and common reviewer demands (“Model 3”):
*Equation 9:*
$y \sim \beta_0 + \beta_{1\text{-}3}(Diagnosis) + \beta_4(Gender) + \beta_5(Age) + \beta_6(pH) + \beta_7(PMI) + \beta_{8\text{-}9}(RateOfDeath) + \beta_{10}(RNADegradation)$

Differential expression analysis was conducted using the limma pipeline [32], using an empirical Bayes moderation of the standard errors towards a common value to reduce the influence of gene-specific outliers and noise. P-values were corrected for multiple comparisons following the Benjamini-Hochberg method (FDR or q-value) within the function *mt.rawp2adjp* (package *multtest* v2.32.0 [16]).

***Maycox et al. 2009:***

***Study overview:*** Maycox et al. [24] used microarray (Affymetrix HG-U133Plus2.0 GeneChips) to characterize gene expression in post-mortem BA10 tissue from subjects with Schizophrenia and non-psychiatric control subjects. Donor recruitment and the collection of BA10 tissue was conducted by Charing Cross Hospital (CCHPC). The subjects with Schizophrenia were elderly residents in long-stay nursing facilities who had become ill before the introduction of antipsychotic medication. The typical duration between illness onset and death was 5 decades, with 3 of those decades eventually spent on neuroleptic drugs (one subject was neuroleptic naïve). The subjects were diagnosed prospectively using DSM-III criteria, and their medical history was screened to rule out neurological illness, with additional histological confirmation. Control subjects came from the community, CCHPC, and local nursing homes, and were reported to have died due to similar causes ([24], information about manner/rate of death was not included in the metadata). Tissue from both schizophrenia and control subjects was collected and stored in parallel. In general, the brain pH was lower (interquartile range between 6.0-6.7) indicating greater hypoxia prior to death, the post-mortem interval was shorter (interquartile range between 3-13 hrs), and the age was older (interquartile range between 55-90 yrs) for the samples in this dataset in comparison to our qPCR experiment. Following screening for RNA quality (RIN>6), the remaining samples were processed in four batches balanced for diagnosis for target generation and hybridization procedures (*n*=57).

***Preprocessing:*** We imported the microarray .CEL files and metadata from Gene Expression Omnibus (https://www.ncbi.nlm.nih.gov/geo/, Accession # GSE17612) using the "GEOquery" R package (v.2.56.0, [27]). We summarized these data using Robust Multiarray Average (RMA [28]) using a custom up-to-date chip definition file (.cdf) to define probe-to-transcript correspondence ("hgu133plus2hsentrezgcdf_25.0.0" downloaded from http://brainarray.mbni.med.umich.edu/Brainarray/Database/CustomCDF/CDF_download.asp [29]). This process included background subtraction, log(2)-transformation, and quantile normalization. Gene Symbol annotation for probeset Entrez gene ids were provided by the R package *org.Hs.eg.db.* (v.3.11.4). To control for technical variation, the sample processing batches were estimated using the microarray chip scan dates extracted from the .CEL files (using the function *protocolData* in the GEOquery package [27]). RNA degradation was estimated computationally using the 3′/5′ intensity ratios for the probes for each gene (R package AffyRNADegradation, v.1.34.0, [30]).

***Quality Control:*** The original study had a sample size of *n*=57, but 6 samples were removed during the quality control that accompanied the original analysis, leaving a final sample size of *n*=51 (CTRL: n=23, SCHIZ: n=28, [24]). Only data from these 51 samples was released publicly on GEO by the original authors. In these remaining samples, the overall median sample-sample correlation was high (median=0.988), implying good technical consistency. Principal components analysis and the visualization of sample-sample correlations revealed an additional two outlier samples, which were removed from the dataset. There was no evidence of sample mismatch or sample mixing as indicated by agreement between subject gender and the strongly bimodal distributions for sex chromosome gene expression (XIST, RPS4Y1, DDX3Y, [31]). The final sample size following quality control was *n*=55 (CTRL: *n*=23, SCHIZ: *n*=26).

***Differential Expression Analysis:*** Brain pH was lower in the Schizophrenia subjects (*pH vs. Diagnosis*: F(1,48)=25.04, p=7.95e-06). All other available demographic and technical variables (Age, PMI, RNADegradation, Gender, ScanDate) were confirmed to be balanced across diagnosis groups ($p$>0.32) using either a chi-square test (categorical vs. categorical) or ANOVA (continuous vs. categorical).

To identify sources of large-scale noise in the data, each of the demographic and technical variables was examined in relationship to the top principal components of variation using either ANOVA or linear regression. PC1 was related to pH (p=0.00117), age (p=0.00848), and RNA degradation (p=3.89e-06). PC2 was related to pH (p=0.0274), PMI (p= 0.0496), and Age (p=7.36e-05). PC3 was not significantly related to any of the demographic or technical variables. PC4 was related to ScanDate (p=7.53e-05), RNA degradation (p= 0.00275), and gender (p=0.0407)

Based on these results, we chose to examine results from three models:

Basic model controlling for confound ("Model 1")
*Equation 10:* $y \sim \beta_0 + \beta_1(Diagnosis) + \beta_2(pH)$

Model with confound and largest implicated sources of noise ("Model 2"):
*Equation 11:*
$\beta_0 + \beta_1(Diagnosis) + \beta_2(pH) + \beta_3(RNADegradation) + \beta_4(Age) + \beta_5(PMI)$

Model with implicated sources of noise and common reviewer demands ("Model 3"):
*Equation 12:* $y \sim \beta_0 + \beta_1(Diagnosis) + \beta_2(Gender) + \beta_3(Age) + \beta_4(pH) + \beta_5(PMI) + \beta_{7\text{-}8}(ScanDate) + \beta_9(RNADegradation)$

Differential expression analysis was conducted using the limma pipeline [32], using an empirical Bayes moderation of the standard errors towards a common value to reduce the influence of gene-specific outliers and noise. P-values were corrected for multiple comparisons following the Benjamini-Hochberg method (FDR or q-value) within the function *mt.rawp2adjp* (package *multtest* v2.32.0 [16]).

***Meta-analysis of Schizophrenia Effects in Iwamoto and Maycox:***

***Comparison of BA10 differential expression results:*** The differential expression results (Models 1-3) associated with each diagnosis and each co-variate (Age, PMI, pH, Gender, RNADegradation) in the two microarray datasets were aligned by EntrezID. The differential expression results from the two datasets (Log2FC and Tstats) were then compared using scatterplots and a correlation matrix. The differential expression results from the microarray experiments were then similarly compared to the results from our qPCR experiments by aligning the results by official gene symbol.

***Meta-analysis:*** We performed a meta-analysis of the effects of Schizophrenia across the two microarray datasets. To do this, we applied random effects modeling to the respective beta coefficients (log2FC) and accompanying sampling variance (SE^2) derived from the differential expression output from each dataset using the *rma()* function within the *metafor* package [33]. P-values were corrected for multiple comparisons following the Benjamini-Hochberg method (FDR or q-value) within the function *mt.rawp2adjp* (package *multtest* v2.32.0 [16]).

When designing our differential expression analyses, we had originally assumed that that the results from Model 1 would be unbiased but poorly control for noise, that Model 3 would be overfitted but strongly control for noise, and that the results of Model 2 would be the most robust. However, after running the meta-analysis using the output from all three models, we discovered that the top differentially expressed genes produced by Models 1 & 2 suspiciously included many sex-chromosome genes, which no longer showed a relationship with diagnosis after controlling for gender (Model 3). Therefore, Model 3 was chosen as the final preferred model for both the findings from each individual study and for use in the meta-analysis.

**Comparisons with Previous Published Results:**

***Comparison with RNA-Seq Results from the Dorsolateral Prefrontal Cortex (DLPFC) and Neighboring Regions***

To gain insight into the similarities and differences between the effects of Schizophrenia and Bipolar Disorder on gene expression in BA10 in comparison to better-studied neighboring cortical areas, we compared the results from our two BA10 qPCR experiments and microarray meta-analysis to two large previously-published meta-analyses of Dorsolateral Prefrontal Cortex (DLPFC) or frontal cortex RNA-Seq data (Table S1 in [18], data from CommonMind Consortium: https://www.synapse.org/cmc and PsychEncode Consortium: https://www.synapse.org/pec). The Schizophrenia meta-analysis included PsychEncode (“GVEX”) RNA-Seq data collected as part of the “Array Collection” and the “New Collection” from the Stanley Medical Research Institute (SMRI) as well as a subset of matched case/control CommonMind Consortium RNA-Seq data from DLPFC samples (BA9/BA46) from brain banks at the Mount Sinai Icahn School of Medicine, University of Pittsburgh, University of Pennsylvania and (final sample sizes from personal communication with Dr. Michael Gandal (2021-09-09): SCHIZ meta-analysis n=384: 181 SCHIZ, 203 CTRL). The Bipolar Disorder meta-analysis included GVEX RNA-Seq samples and a subset of matched case/control samples from two brain banks within the CommonMind Consortium data (University of Pittsburgh, Mount Sinai; final sample sizes from personal communication with Dr. Michael Gandal (2021-09-09): Bipolar Disorder meta-analysis n=171: 70 BP, 101 CTRL).

We chose to emphasize the comparison of our BA10 results with the results from the two RNA-Seq meta-analyses by [18] for several reasons: 1) The sample sizes included in the RNA-Seq meta-analyses were large and likely to decrease noise in the results, 2) The samples used in the RNA-Seq meta-analyses were from high quality brain banks, and had a similar distribution of sample quality characteristics and demographics to our BA10 samples (with the exception of slightly lower average brain pH), 3) Both RNA-Seq meta-analyses were enriched with a large portion of tissue derived from grey-matter focused dissections. The samples from both University of Pittsburgh and Stanley Medical Research Institute were derived from grey-matter focused dissections ([13], Maree J. Webster, PhD personal communication 2-24-2021), whereas the samples from the Mount Sinai and University of Pennsylvania brain banks were derived from dissections that included both white matter and grey matter [13], 4) Both RNA-Seq meta-analyses carefully controlled for potential confounds and sources of noise in the datasets in a manner comparable to our BA10 analyses, including matching case/control samples to reduce confounding variation, applying batch corrections based on library preparation date, and using differential expression models that further controlled for important sources of variation in the dataset (CMC dataset: age, sex, RIN, RIN2, ethnicity, PMI, pH, PC1 & PC2 from sequencing characteristic matrix; GVEX dataset: age, sex, RIN, RIN2, ancestry (via PCA), PMI, institution, PC1 & PC2 from sequencing characteristic matrix).

To allow comparison with our BA10 results, we mapped the Ensembl ID annotation provided with the RNA-Seq meta-analysis results to Entrez ID and official human gene symbol using the org.HS.eg.db package in R [34]. Out of the original 15823 Ensembl IDs in the dataset, there were 1614 that could not be mapped to Entrez ID or gene symbol, 365 instances of mapping to more than one gene symbol (mostly MIR and LOC gene families), and 36 instances of mapping to the same symbol as another Ensembl ID, producing a mapped dataset of 16188 rows, 14209 of which contained both a unique Ensembl identifier (no multimapping) and Entrez and Gene Symbol annotation.

***Comparison with Microarray Results from DLPFC Grey-Matter Focused Dissections***

The comparison of our BA10 results with the DLPFC/frontal cortex RNA-Seq meta-analysis results revealed a strong overall similarity between the effects of diagnosis in the two regions, but also highlighted several interesting points of divergence. To gain insight into how much of this divergence might be driven by differences in dissection, we also ran comparisons between our BA10 results and results from a previous re-analysis (Table S11 and Table S12 in [13]) of two publicly-available DLPFC microarray datasets from experiments performed using grey matter dissected tissue (GEO Accession#: GSE53987 [35], n=66: 18 CTRL, 17 BP, 14 SCHIZ, 17 MDD; GEO Accession#: GSE21138 [36], n=54: 27 CTRL, 27 SCHIZ). These datasets included samples from BA46 (DLPFC) from the University of Pittsburgh [35,37] and Mental Health Research Institute Australia [36] brain banks. Both of these datasets had much smaller sample sizes than the Gandal et al. RNA-Seq meta-analyses, and the Lanz et al. dataset was derived from tissue with a greater incidence of hypoxia than what is found in our BA10 datasets (average pH 6.3). Despite these weaknesses, the results between these two grey matter-focused datasets showed a remarkable degree of convergence with each other (correlation between Lanz et al. and Narayan et al. SCHIZ Log2FC: R=0.457), suggesting that the results were high quality. Also, similar to our BA10 analyses, the reanalysis of these two datasets [13] controlled for potential confounds and sources of noise in the datasets using differential expression models similar to those used in our current experiments (Age, Sex, RNA Degradation, PMI, pH, batch (scan date, when applicable)). When considering the results, it is important to acknowledge that samples from the University of Pittsburgh brain bank were also used in the CommonMind Consortium data that was included in the RNA-Seq meta-analysis in [18], therefore it is possible that tissue from some of the same subjects may have been profiled in the two studies. Thus the results derived from the Lanz et al. dataset and Gandal et al. RNA-Seq meta-analysis may offer separate insight but not be fully independent.

The Lanz et al. and Narayan et al. datasets were collected using the Affymetrix U133Plus2 microarray, and probes were reannotated using Entrez ID during the original re-analysis to produce gene expression summaries for 19,764 unique Entrez IDs [13]. For our current analysis, these Entrez IDs were provided with more up-to-date gene symbol and Ensembl annotation using the using the org.HS.eg.db package in R (v.3.4.1, [34]), mapping 18,480 Entrez-annotated genes to unique (not multi-mapped) Gene Symbol and Ensembl annotation.

***Comparison with Previous Microarray Results from Other Psychiatric Disorders***

As a more exploratory analysis, we also compared our BA10 Schizophrenia and Bipolar Disorder results to previous studies examining the relationship between other psychiatric diagnoses on cortical gene expression. To do this, we used a large previously-published meta-analysis of psychiatric effects in cortical microarray data (SCHIZ: n=159, BP: n=94, Major Depressive Disorder (MDD): n=87, Autism Spectrum Disorder (ASD): n=50, Alcoholism (AAD): n=17, matched CTRLs: n=293, from Table S1 in [18]), which included results from two diagnoses with overlapping symptoms and life experiences to Bipolar Disorder and Schizophrenia: Major Depressive Disorder (MDD) and Alcohol Abuse Disorder (AAD). The MDD results were derived from samples from BA46 (DLPFC) from the University of Pittsburgh brain bank (GEO Accession#: GSE53987 [35,37]) and from BA9 (prefrontal cortex, GEO Accession#: GSE54568, GSE54567) and BA25 (anterior cingulate cortex, GEO Accession#: GSE54572, GSE54571) from the University of Pittsburgh brain bank [38]. Within all of these datasets, gene expression was profiled using the Affymetrix Human Genome U133 Plus 2.0 microarray chips. The AAD results were derived from a small Illumina HumanHT-12 V3 microarray study using superior frontal cortex tissue from the New South Wales Tissue Resource Centre at the University of Sydney (GEO Accession #: GSE29555, [39]). The meta-analysis included both reannotation and statistical control for potential confounds and sources of noise in the datasets in a manner comparable to our BA10 analyses, including matching case/control samples to reduce confounding variation, applying batch corrections based on chip scan date (when available), and regressing out potential sources of confounding variation or noise in the dataset (Lanz et al. dataset: RNA quality (5'/3' bias), Chang et al. datasets: Brain Region, Sex, Age, Race, PMI, pH RIN, RNA quality (5'/3' bias), Ponomarev et al. dataset: Age, Sex, PMI, pH, and RIN). We did not use the Schizophrenia or Bipolar results from this meta-analysis as a point of comparison due to circularity (they included the BA10 microarray dataset from Maycox et al. [24]). We mapped the Ensembl ID annotation provided with these results to more up-to-date Entrez ID and official human gene symbol using the org.HS.eg.db package in R (v.3.4.1, [34]). This mapping produced 18,981 Ensembl-annotated genes that mapped to unique (not multi-mapped) Gene Symbol and Entrez annotation in the MDD meta-analysis results, and 18,844 in the AAD results.

***Examining sensitivity of qPCR for measuring low-level expression:***

Several of the top genes associated with diagnosis in our qPCR experiments (HTR2B, DRD4, GPHN) had strikingly large or clean effects in our dataset but had either much smaller or non-existent effects in both BA10 microarray or DLPFC comparison datasets. To determine whether this discrepancy might be due to the greater sensitivity of qPCR to low-level expressed genes than microarray or RNA-Seq methods, we first determined how sensitive our qPCR measurements were by examining the relationship between the average Cq and the % difference between replicate measurements for all transcripts quantified in the GABAGLU and DA5HT datasets (144 target and reference transcripts, with 5 measured transcripts in each experiment representing the same reference genes). To perform this analysis, we used the data following removal of a replicate sample from two subjects for which the whole stem of wells did not amplify properly (>95% of measurements were missing values ("No Amp" or "Undetermined")), but prior to any additional QC. We binned the values for % difference between replicate measurements by 1 unit intervals of average Cq (per replicate pair), and then calculated both the median % difference between replicate measurements and upper 95$^{th}$ percentile. Bins with only 1 set of replicate measurements were excluded (Cq 9 and 10).

Next, to compare the sensitivity of the gene expression measurements in our qPCR experiment to RNA-Seq, we compared our results to results from our previous re-analysis of a large RNA-Seq study from a neighboring cortical region (Table S9 in [13], data from the CommonMind Consortium: https://www.synapse.org/cmc, n=603 subjects (514 subjects following QC): 285 CTRL, 263 SCZ, 47 BP, 8 Affective Disorder). This dataset included DLPFC samples from BA9/BA46 from brain banks at the Mount Sinai Icahn School of Medicine, University of Pittsburgh, University of Pennsylvania. The samples taken from University of Pittsburgh were from grey-matter focused dissections, whereas the samples from the Mount Sinai and University of Pennsylvania brain banks were derived from dissections that included both white matter and grey matter [13]. Our previous reanalysis of this dataset [13] controlled for potential confounds and sources of noise in the datasets using differential expression models similar to those used in our current experiments (pH, PMI, sex, age, RIN, institution, manner of death). Importantly, the results were filtered using liberal criteria for minimal expression threshold (CPM>1 in at least 50 individuals (roughly 10% of subjects)), leaving expression data from 22,053 Ensembl-annotated genes available for comparison [13]. To perform our current comparison with the qPCR results, we mapped the Ensembl ID annotation provided with the RNA-Seq results to more up-to-date Entrez ID and official human gene symbol using the org.HS.eg.db package in R [34]. This mapping produced 16,997 Ensembl-annotated genes that mapped to unique (not multi-mapped) Gene Symbol and Entrez ID. We then aligned these results with our qPCR results using official gene symbol and compared the average Cq for each transcript quantified in our two qPCR experiments to 1) the average Log2CPM gene level summary output within the RNA-Seq study, 2) whether the transcript was quantifiable in the RNA-Seq dataset (numeric value vs. NA).

***Exploring Functional Patterns in the Differential Expression Results:***

***Annotation with General Functional Information:*** The differential expression results were annotated with general functional information from three different sources: 1) functional categories related to neurotransmission assigned *a priori* by the lead author based on published literature (used for designing the custom qPCR cards), 2) information about cortical cell type specific gene expression from the BrainInABlender database (Suppl. Table 1 in [13], referencing both official Human and Mouse Gene Symbol to capture broadest evidence base), and 3) Predicted cell type specificity of expression (correlation between gene expression and BrainInABlender cell type index within a large Pritzker DLPFC microarray dataset, Suppl. Table 8 in [13]). We also cross-referenced our list of top differentially expressed genes (six genes with FDR<0.10 in our qPCR experiment) with the list of mammalian phenotypes created by perturbation in the expression of these genes within the Mouse Genome Informatics database (MGI, http://www.informatics.jax.org/phenotypes.shtml, [40]).

***Exploring Overlap Between our Results and Genetic Variants Associated With Diagnosis:*** We also examined overlap between our differential expression results and genetic variants that have previously been associated with Schizophrenia using the SZDB 2.0 database (http://szdb.org, [41,42]). This database includes 571 unique genes implicated by overlap with variants identified in large genome wide association studies (GWAS: CLOZUK, PGC2), 7 of which were included as qPCR targets in our study, and 408 unique genes implicated by CNV studies, 1 of which was included as a qPCR target in our study. The SZDB 2.0 database also includes noisier evidence: 5830 unique genes implicated by Exome Sequencing studies, 45 of which were included as qPCR targets, and 8578 unique genes that show correlated expression with the variants identified in large GWAS studies (CLOZUK, PGC2) as identified using brain-related eQTL databases. Finally, 30 of our qPCR targets were implicated by linkage association studies, and 6 were implicated by differential methylation studies, but this evidence is more difficult to interpret because the overall number of associations present in the SZDB 2.0 database was not available.

***Exploring Overlap Between our Results and Stress-Related Differential Gene Expression:*** We also examined the overlap between our BA10 qPCR results and genes repeatedly implicated within RNA-Seq studies examining the effect of stress on brain gene expression in mice as documented through the Stress Mice Portal (http://hpc-bioinformatics.cineca.it/stress_mice/ , [43]). Specifically, the Stress Mice Portal re-analyzed the data from 18 SRA Bioprojects (RNA-Seq) examining the effects of different stressors on the mouse brain (n=751 samples, 101 stress vs. control comparisons; brain regions: hippocampus, nucleus accumbens, prefrontal cortex, amygdala, bed nucleus of the stria terminalis, ventral tegmental area, hypothalamus). Following re-analysis, they compiled lists of commonly differentially expressed genes (no formal meta-analysis; threshold of absolute value of log2 fold change >= 0.38, adjusted p-value <= 0.05). Within the database, 1925 unique genes were found to be differentially expressed in at least 2 stress RNA-Seq Bioprojects (out of 18, from Table 3, Table S1, Table S2 in [43]) and 27 out of our 111 qPCR targets were included on this list (24%).

***Exploring Overlap Between our Results and Drug-Related Differential Gene Expression:*** We also examined the similarity between our top differential expression results and differential expression associated with exposure to substances of abuse in the post-mortem cortex (alcohol abuse disorder: [18,19], opioid use disorder [20]) and prescription drugs in the prefrontal cortex of primates [21] and in cell culture and other publicly-available transcriptional profiling datasets (https://maayanlab.cloud/DGB/, [22]). To be included in this comparison, a gene needed to show strong evidence of differential expression in BA10: either FDR<0.10 in our qPCR experiment (gene symbol with grey shading) or p<0.05 (Log2FC in bold text) and consistent direction of effect in two independent datasets from BA10 or BA10 and DLPFC/frontal cortex (20 genes total, **Fig 3**).

The Kapoor et al. [19] study examined the effects of alcoholism (subjects with alcoholism: n=65, control subjects: n=73) in the post-mortem prefrontal cortex using RNA-Seq. We accessed the differential expression statistics for each of our 20 genes of interest using gene symbol as the identifier within their interactive webapp (https://lcad.shinyapps.io/coga-inia/, access code: coga-inia, accessed 5/11/2022).

The Seney et al. [44] study examined the effects of opioid use disorder (subjects with OUD: n=20, control subjects: n=20) in the post-mortem dorsolateral prefrontal cortex and nucleus accumbens using RNA-Seq. This study used a cortical dissection method that minimized the inclusion of white matter. Notably, all of the subjects with OUD also died of overdose. We extracted the differential expression statistics for each of our 20 gene of interest using gene symbol as the identifier in supplementary Data File S2.

The Martin et al. [21] study examined the effect of acute and chronic (4 week) exposure to typical and atypical antipsychotics (haloperidol and olanzapine, respectively) and chronic (6 week) exposure to the hallucinogen phencyclidine (PCP) on gene expression in the prefrontal cortex of Cynomolgus monkeys. Within this study, two separate transcriptional profiling experiments were performed using the same tissue using the Affymetrix HG6800 and Affymetrix HGU95A/HGU95Av2 microarray platforms, respectively. We referenced the results from chronic exposure to each of the three drugs from both microarray platforms (Supplemental tables 1&2 in [21]) using official gene symbol, and considered a gene to be potentially differentially expressed in association with the drug if it had a nominal p<0.05 in either experiment.

The Drug Gene Budger database (https://maayanlab.cloud/DGB/, [22]) compiles differential expression results in relationship to exposure to small molecules/perturbagens into an easily-searchable web-interface. These differential expression results are derived from gene expression signatures extracted from publicly-available datasets, including the LINCS L1000 dataset and the original Connectivity Map (CMap) dataset (derived from cell lines) and datasets on Gene Expression Omnibus (GEO, derived from a variety of tissues and experiments). For each of our top differentially expressed genes, we downloaded the full list of perturbagens that have been shown to elicit differential expression (q-value/FDR<0.05), and cross-referenced this list with both a list of drugs/drug categories that were present in the metadata for our subjects with SCHIZ and BP, as well as with lists of commonly-prescribed therapeutics. The therapeutic drugs included typical antipsychotics (thiothixene, thioridazine, trifluoperazine, fluphenazine, haloperidol, chlorpromazine), atypical antipsychotics (clozapine, quetiapine), antidepressants (amitriptyline, sertraline, trimipramine, paroxetine, nortriptyline, fluoxetine), and anticonvulsants (valproic acid, carbamazepine). To provide potential insight into the effects of stress on gene expression, we also extracted results for corticosteroids (dexamethasone, cortisone, triamcinolone, betamethasone, prednisolone, hydrocortisone, corticosterone, fludrocortisone, budesonide). We did not find many results related to common drugs of abuse (alcohol/ethanol, tobacco/nicotine, cannabinoids, stimulants, opiates), but it is worth noting that an *absence* of evidence linking a gene to a drug in this database may not be meaningful, since we lack information regarding the full list of drugs examined and the presence of measurable gene expression for each gene within the full body of treatments and experiments surveyed. We then examined the results from each category of drug for direction of effect irrespective of the dataset of origin or treatment regimen.

#### Supplementary Results

##### Full statistical reporting for results

The complete statistical reporting for the full concatenated qPCR results for diagnosis and all co-variates can be found in **Table S 5**. The complete statistical reporting for the full results from the BA10 microarray re-analysis and meta-analysis can be found in **Table S 8**. The full statistical reporting for the correlation and regression analyses comparing the pattern of gene expression (Log2FC) associated with different diagnoses, variables, and datasets can be found in **Table S 9.**

##### Sensitivity to low-level expression

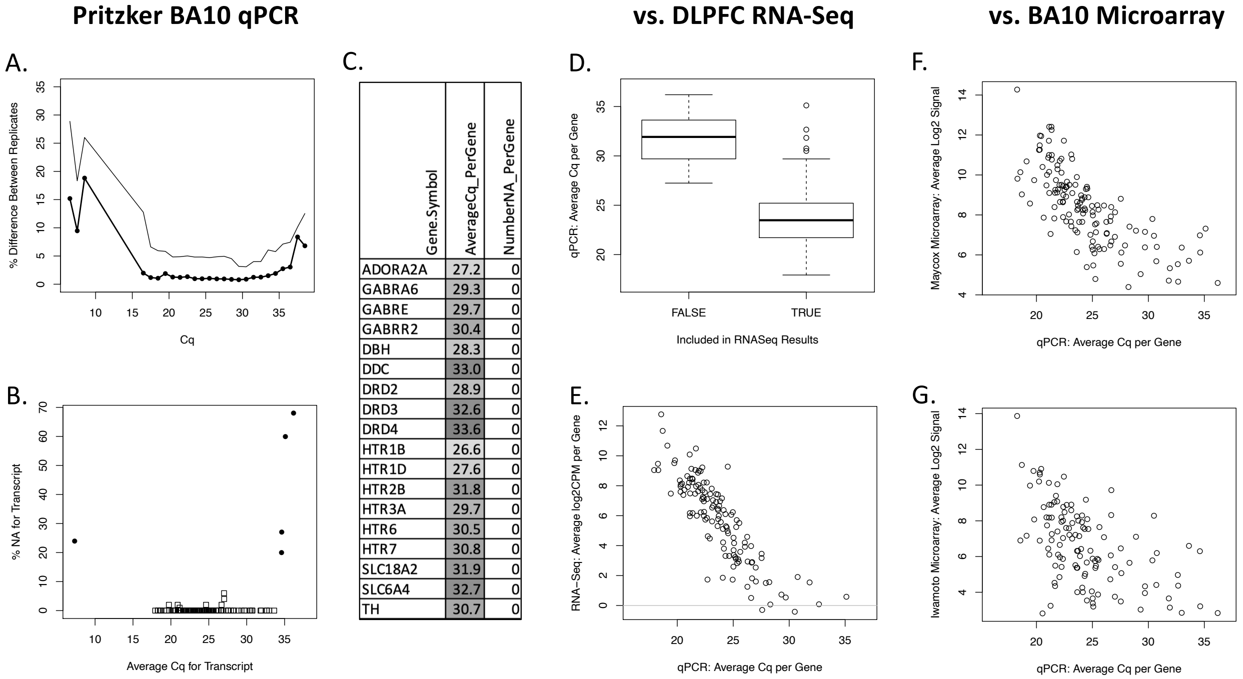

| Gene.Symbol | AverageCq_PerGene | NumberNA_PerGene |
|---|---|---|
| ADORA2A | 27.2 | 0 |
| GABRA6 | 29.3 | 0 |
| GABRE | 29.7 | 0 |
| GABRR2 | 30.4 | 0 |
| DBH | 28.3 | 0 |
| DDC | 33.0 | 0 |
| DRD2 | 28.9 | 0 |
| DRD3 | 32.6 | 0 |
| DRD4 | 33.6 | 0 |
| HTR1B | 26.6 | 0 |
| HTR1D | 27.6 | 0 |
| HTR2B | 31.8 | 0 |
| HTR3A | 29.7 | 0 |
| HTR6 | 30.5 | 0 |
| HTR7 | 30.8 | 0 |
| SLC18A2 | 31.9 | 0 |
| SLC6A4 | 32.7 | 0 |
| TH | 30.7 | 0 |

***Figure S 13. Our qPCR methodology can reliably and accurately measure low-level expression.***

*Using qPCR, we were able to reliably quantify the expression of genes that have too little expression to be reliably measured using RNA-Seq or microarray. This included neurotransmitter receptors that are important pharmaceutical targets in the treatment of SCHIZ and BP.* ***A.*** *qPCR cycle threshold (Cq) measurements were highly reliable when measurements fell in the range of the majority of our target genes (between Cq 18-35). The figure shows the percent difference between duplicate samples for the Cq measurements for each target and reference gene binned by the average Cq measurement for those duplicates. Black dots indicate the median % difference between duplicates, the thinner line represents the 95th percentile of % difference between duplicates.* ***B.*** *There is minimal missing (unmeasurable) Cq data ("NA") when measurements fell in the range of the majority of our target genes (between Cq 18-34.5). The figure shows the number of "NA" measurements per gene (y-axis, both target and reference) in relationship to the average Cq measurement for that gene (x-axis).* ***C.*** *We* *were able to characterize the relationship between diagnosis and the expression of many neurotransmitter signaling-related genes that have particularly low levels of cortical expression (Cq>27).* ***D.*** *Average Cq measurements (y-axis) for genes that had too low of expression to be considered quantifiable in a large RNA-Seq study in the neighboring DLPFC (log(2)CPM<0 in >90% of the large CommonMind Consortium sample [13,45]). This included Tyrosine Hydroxylase (TH), Dopa Decarboxylase (DDC), Solute Carrier Family 18 Member A2 (SLC18A2), Dopamine Receptors D2, D3, and D4 (DRD2, DRD3, DRD4), 5-Hydroxytryptamine Receptor 3A (HTR3A), Adenosine A2a Receptor (ADORA2A), and Gamma-Aminobutyric Acid Type B Receptor Subunit 2 (GABBR2).* ***E.*** *We found additional indications that RNA-Seq might be less well-suited to studying low level expression than qPCR. At higher levels of expression (Cq<30), we observed a clear linear relationship between RNA-Seq average log2CPM observed in the large CommonMind Consortium sample [13,45]) from the neighboring DLPFC and the average Cq per gene observed in our qPCR study. At low levels of expression, this linearity was lost, showing a plateau for genes with an average Cq >30 in our study.* ***F-G.*** *We saw similar indications that microarray might be less well-suited to studying low level expression than qPCR. At higher levels of expression (Cq<27), we observed a clear linear relationship between average microarray log2 signal observed in previous BA10 studies (**F:** [24], **G:** [23]) and the average qPCR cycle threshold (Cq) per gene observed in our qPCR study. At low levels of expression, this linearity was lost, showing a clear plateau for genes with an average Cq>27 in our study.*

#### Similarity to Effects in Previous BA10 Microarray Studies

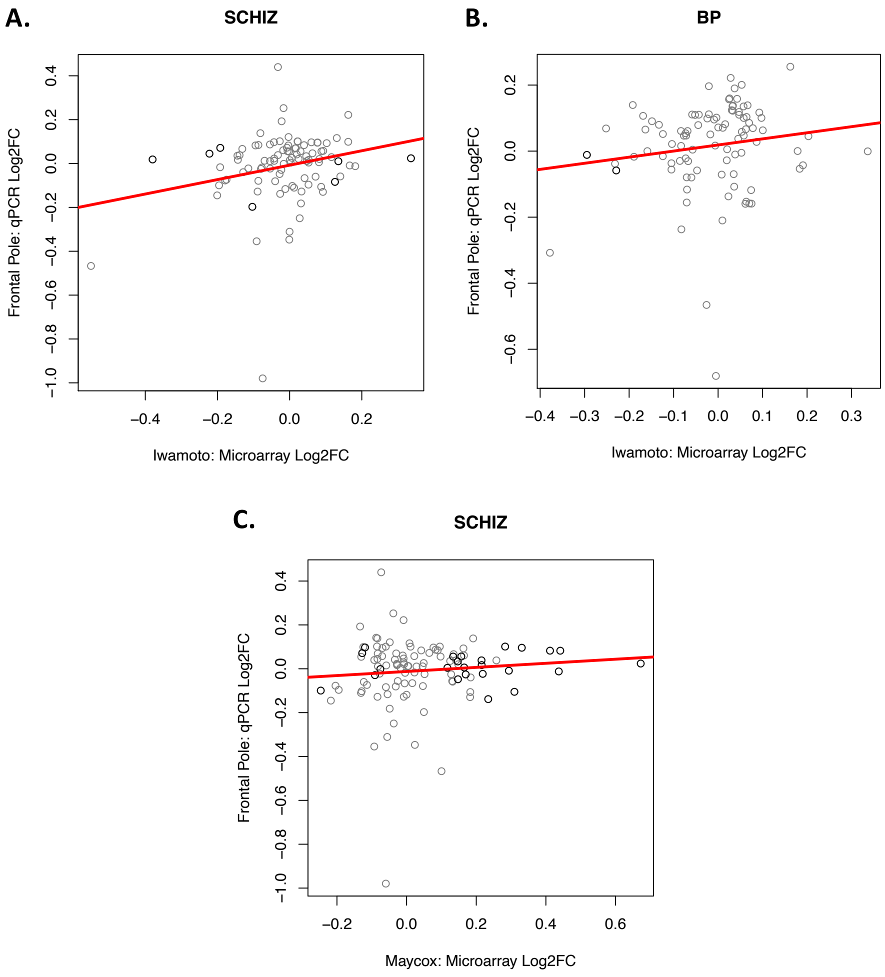

***Figure S 14. The effects of diagnosis in our BA10 qPCR study partially replicate the effects of diagnosis in our re-analysis of two BA10 microarray studies.***
***A**) There was a weak positive correlation between the effects of SCHIZ (Log2FC) measured in our BA10 qPCR results and the effect of SCHIZ (Log2FC) in our re-analysis of the BA10 microarray data from Iwamoto et al.* [23] *(R=0.242, Beta ± SE = 0.329 ± 0.136, T(93) = 2.41, p = 0.0179). **B**) There was a weak positive correlation between the effects of BP(Log2FC) measured in our BA10 qPCR results and the effect of BP (Log2FC) in our re-analysis of the BA10 microarray data from Iwamoto et al.* [23] *but it did not reach significance (R=0.148, Beta ± SE = 0.185 ± 0.128, T(93) = 1.45, p = 0.151*not sig). C) There was a weak positive correlation between the effects of SCHIZ (Log2FC) measured in our BA10 qPCR results and the effect of SCHIZ (Log2FC) in our re-analysis of the BA10 microarray data from Maycox et al.* [24] *but it did not reach significance (R=0.0921, Beta ± SE = 0.0932 ± 0.0969, T(108) = 0.962, p = 0.338*not sig).*

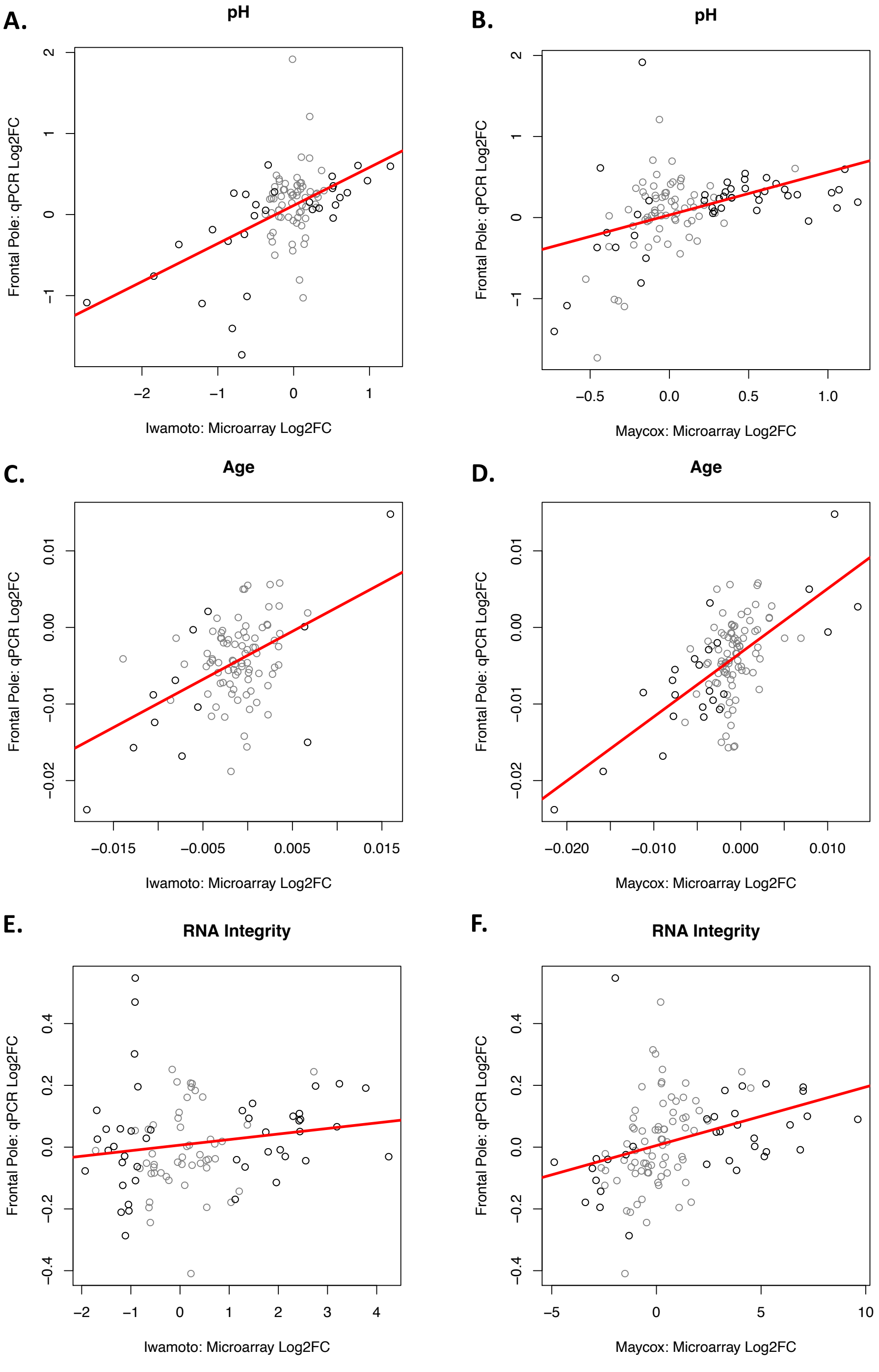

***Figure S 15. The effects of co-variates (pH, Age, RNA Integrity) in our BA10 qPCR study strongly replicate the effects of co-variates in our re-analysis of two BA10 microarray studies.***

*We saw a strong correlation between the differential expression observed in our qPCR study (Log2FC) and the differential expression observed within our re-analysis of two BA10 microarray studies (Log2FC, [23,24]) for variables with larger effect sizes than diagnosis, such as hypoxia (brain pH), RNA integrity, and Age. This suggested that the weakness of the correlation* *between the effects of diagnosis in our qPCR dataset and the BA10 microarray datasets might simply be due to the challenge of measuring small effect sizes in noisy data with a small sample size and not due to some inherent difference arising from comparing results derived from different transcriptional profiling platforms.* ***A)*** *The effects of hypoxia (brain pH) in our qPCR dataset strongly correlate with the effects of pH in Iwamoto et al. (R=0.515, Beta ± SE = 0.470 ± 0.0812, T(93) = 5.78, p = 9.78e-08),* ***B)*** *and with the effects of pH in Maycox et al. (R=0.431, Beta ± SE = 0.529 ± 0.107, T(108) = 4.97, p = 2.57e-06).* ***C)*** *The effects of age in our qPCR dataset strongly correlate with effects of age in Iwamoto et al. (R=0.486, Beta ± SE = 0.628 ± 0.117, T(93) = 5.36, p = 6.00e-07),* ***D)*** *and with the effects of age in Maycox et al. (R=0.620, Beta ± SE = 0.836 ± 0.102, T(108) = 8.21, p = 5.04e-13).* ***E)*** *The effect of RNA Integrity (RIN) in our qPCR dataset weakly correlates with effect of RNA Integrity in Iwamoto et al. (R=0.165, Beta ± SE = 0.0180 ± 0.0111, T(93) = 1.61, p = 0.110*not sig) and does not reach significance,* ***F)*** *but strongly correlates with the effects of RNA Integrity in Maycox et al. (R=0.341, Beta ± SE = 0.0190 ± 0.00504, T(108) = 3.76, p = 0.000273).*

**BA10 Microarray Meta-Analysis**

| | BP | | SCHIZ | | | | MDD |
|---|---|---|---|---|---|---|---|
| | BA10 | | BA10 | | | | BA10 |
| Gene Symbol | Pritzker_qPCR | Iwamoto_Microarray | Pritzker_qPCR | Pritzker_Microarray_MetaAnalysi | Iwamoto_Microarray | Maycox_Microarray | Iwamoto_Microarray |
| TUBB7P | NA | -0.16 | NA | **-0.25** | **-0.23** | **-0.28** | -0.11 |
| TIE1 | NA | **-0.19** | NA | **-0.23** | **-0.21** | **-0.22** | -0.01 |
| HSD17B8 | NA | -0.07 | NA | **-0.15** | **-0.19** | **-0.13** | -0.02 |
| URM1 | NA | -0.08 | NA | **-0.12** | **-0.19** | **-0.11** | -0.06 |
| CACYBP | NA | 0.04 | NA | **0.17** | 0.15 | **0.19** | 0.03 |
| GIT2 | NA | **0.35** | NA | | **0.16** | -0.01 | 0.12 |

***Figure S 16. Differentially expressed genes identified in BA10 via microarray that were not included as targets in our qPCR experiment.***

*Our re-analysis of publicly available BA10 data [23,24] and subsequent meta-analysis identified significant diagnosis effects (FDR<0.10) for 6 genes that were not related to neurotransmission or included as targets in our qPCR study: a downregulation of TUBB7P, TIE1, HSD17B8, URM1 and upregulation of CACYBP, GIT2 in association with SCHIZ. These effects were mirrored in the data for BP and Major Depressive Disorder (MDD). The table illustrates the Log2FC associated with diagnosis from our re-analysis of the Iwamoto et al. [23] and Maycox et al. [24] BA10 microarray studies, as well as our meta-analysis of the two studies. Formatting conventions are similar to* ***Figure 3****, blue=decreased in the diagnosis group, red=increased in the diagnosis group, bold=p<0.05, bold+underline=FDR<0.10.*

#### Exploratory Analyses: Factors Contributing to Diagnosis-Related Gene Expression

##### *Overlap Between our Diagnosis Results and Therapeutic-Related Differential Gene Expression*

**A. Exploratory: Effect of therapeutics on gene expression**

| Gene Symbol | BA10: Diagnosis: Pritzker_qPCR_Bipolar | BA10: Diagnosis: Pritzker_qPCR_Schiz | BA10: Diagnosis: Microarray_MetaAnalysis_Schiz | Antipsychotics: Pritzker_Antipsychotics | Antipsychotics: DrugBudger_Antipsychotics | Antipsychotics: Martin_Haloperidol | Antipsychotics: Martin_Olanzapine | Antidepressants: Pritzker qPCR_Antidepressants | Antidepressants: DrugBudger_Antidepressants |
|---|---|---|---|---|---|---|---|---|---|
| HTR2B | **-0.68** | **-0.98** | -0.05 | 0.06 | Up (6+/2-) | 0.08 | 0.01 | -0.46 | N |
| SST | -0.31 | **-0.47** | -0.22 | 0.37 | Up (4+) | **0.22** | 0.10 | 0.01 | Up (1+) |
| DRD4 | **-0.47** | **-0.35** | -0.04 | 0.23 | Mixed (1-/1+) | 0.03 | 0.00 | 0.19 | Up (1+) |
| BDNF | -0.24 | **-0.35** | -0.10 | **0.45** | Up (6+/4-) | 0.08 | 0.08 | -0.35 | N |
| DRD2 | -0.14 | **-0.25** | -0.04 | 0.09 | Up (10+/4-) | 0.02 | -0.02 | 0.06 | Up (2+) |
| DRD3 | -0.12 | **-0.31** | -0.04 | 0.17 | Up (7+/3-) | **0.06** | -0.01 | -0.01 | Up (2+) |
| HOMER1 | -0.03 | **-0.13** | 0.04 | -0.08 | Down (11-/9+) | | | 0.02 | Down (3-) |
| SLC6A12 | -0.01 | -0.07 | **-0.16** | 0.15 | Up (3+) | 0.04 | -0.04 | 0.20 | N |
| COMT | 0.05 | 0.07 | **-0.13** | **0.34** | Up (8+/1-) | 0.01 | -0.01 | -0.11 | Mixed (1+/1-) |
| SLC6A11 | 0.19 | 0.00 | **-0.05** | -0.02 | Down (2-/1+) | 0.02 | -0.04 | **0.28** | N |
| GABRB1 | 0.07 | **0.12** | 0.07 | -0.02 | Up (3+/2-) | 0.09 | **0.10** | -0.01 | N |
| GRM5 | **0.16** | 0.00 | **0.07** | -0.11 | Up (4+/1-) | 0.10 | 0.02 | **-0.24** | Up (4+/1-) |
| CALB1 | 0.12 | 0.02 | **0.50** | -0.15 | Down (4-/2+) | 0.08 | 0.07 | **-0.25** | Up (2+) |
| ABAT | **0.16** | 0.08 | 0.06 | 0.04 | Down (8-/4+) | 0.04 | 0.02 | 0.04 | Mixed (1-/1+) |
| MAPK1 | **0.14** | 0.09 | 0.06 | -0.08 | Up (5+/4-) | 0.12 | 0.01 | 0.02 | Down (1-) |
| NSF | **0.11** | **0.10** | 0.19 | -0.09 | Down (7-/4+) | | | 0.09 | Down (1-) |
| SNCA | 0.10 | **0.10** | 0.11 | -0.09 | Up (14+/1-) | **0.13** | 0.06 | **0.14** | Up (4+) |
| GPHN | **0.16** | **0.10** | | -0.01 | Down (8-/5+) | | | **-0.13** | Down (2-) |
| MAOB | 0.14 | **0.22** | 0.11 | 0.05 | Down (4-/2+) | **0.13** | 0.03 | **<u>-0.46</u>** | N |
| GFAP | 0.26 | **0.44** | 0.00 | 0.19 | Down (-4) | 0.06 | 0.07 | -0.16 | Down (2-) |

**Log2FC:** **<u>Bold & Underline</u>**: FDR<0.10, **Bold**=p<0.05

**DrugBudger:** # of results (FDR<0.05) with each direction of effect

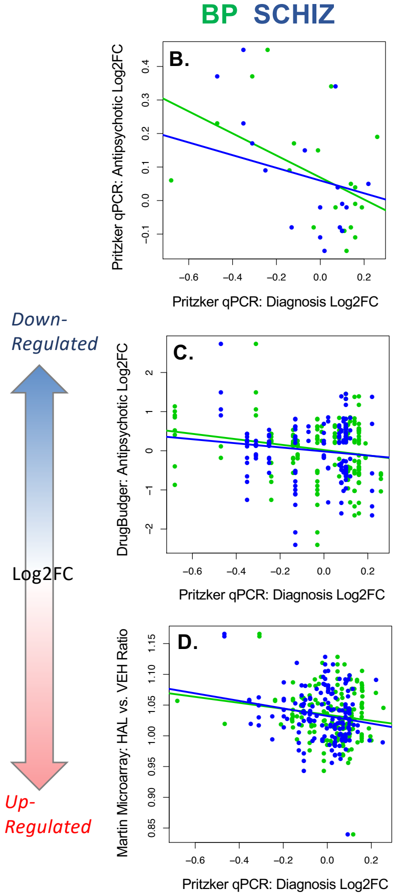

***Figure S 17. Exploratory: The effects of BP and SCHIZ on neurotransmission-related gene expression are often the opposite of the effects of anti-psychotic therapeutics.***
***A)*** *A table overviewing the effects of diagnosis and therapeutic treatment on gene expression (Log2FC) for the 20 genes with the most reliable diagnosis effects in BA10. Altogether, out of all of the results, the down-regulation of HTR2B, SST, BDNF, DRD2, DRD3, and COMT and the upregulation in ABAT, MAOB, NSF, GPHN, and GFAP in association with diagnosis in BA10 seem potentially opposed by therapeutic treatment. In contrast, the upregulation of SNCA in association with diagnosis in BA10 could potentially be an artifact of therapeutics. In general, the table follows the conventions of* ***Figure 3:*** *blue=decreased in the diagnosis group or treatment condition, red=increased in the diagnosis group or treatment condition, bold=p<0.05, bold+underline=FDR<0.10. On the far left side of the figure, there is an abbreviated version of the table in* ***Figure 3****, which illustrates the Log2FC associated with diagnosis from our qPCR dataset and meta-analysis of the Iwamoto et al.* [23] *and Maycox et al.* [24] *BA10 microarray studies. The two tables on the right illustrate* ***1.*** *the effects of effects of antipsychotic and antidepressant drug exposure (Log2FC) estimated while controlling for diagnosis in our dataset. Antipsychotic drug exposure was defined by indication of antipsychotic drug usage within the subjects' clinical records, family interviews, toxicology reports, and coroners reports.* ***2.*** *The effects of antipsychotic and antidepressant exposure on gene expression within the Drug Gene Budger database* [22]*. Drug Gene Budger includes results from both in vitro and in vivo experiments. The*

*ratio of the number of effects in the database that indicate upregulation (+) versus downregulation (-) with drug exposure are indicated in parentheses, regardless of dosage or experiment of origin. N means "not present in database". All effects in the database were significant (FDR<0.05).* ***3.*** *The effects of the antipsychotics chronic haloperidol and olanzapine (vs. vehicle: ratio) on gene expression in the DLPFC in primates as measured by microarray ([21]. To create this figure, the ratio of gene expression from the treatment and vehicle groups from each of the two iterations of the Martin et al. experiment were averaged for each probe, and then aligned with our qPCR results via gene symbol.* ***B-D)*** *We observed a general pattern in which the effects of antipsychotics appeared to oppose the effects of diagnosis. Shown are scatterplots illustrating the negative correlation between the differential expression (Log2FC) associated with antipsychotics and the differential expression (Log2FC) associated with diagnosis in our qPCR study (x-axis: Blue=Bipolar Disorder Log2FC, Green=Schizophrenia Log2FC).* ***B.*** *The effects of diagnosis (Log2FC) on gene expression in BA10 measured in our qPCR study correlated negatively with the effects of antipsychotic drug exposure (Log2FC) estimated while controlling for diagnosis in our dataset (Bipolar vs. Antipsychotics: R=0.464, p=0.0391, Schiz vs. Antipsychotics: R=0.254, p=0.146*not sig). This exploratory analysis was limited to the 20 genes with the most reliable diagnosis effects in BA10 (panel A).* ***C.*** *The effects of diagnosis (Log2FC) on gene expression in BA10 measured in our qPCR study correlated negatively with the effects of antipsychotic drugs on gene expression within the Drug Gene Budger database [22] (BP: p=0.0423, SCHIZ: p=0.0305). This analysis was limited to the 20 genes with the most reliable diagnosis effects in BA10 (169 Drug Gene Budger results total, with gene included in statistical models as a random effect).* ***D.*** *The effects of diagnosis (Log2FC) on gene expression in BA10 measured in our qPCR study tended to correlate negatively with the effects of chronic haloperidol (vs. vehicle: ratio) on gene expression in the DLPFC in primates as measured by microarray ([21], n=162, with gene included in the model as a random effect: SCHIZ: p=0.0602*trend; BP: p=0.179*not sig). We did not see a similar relationship with the average effects of olanzapine (p>0.28). Full statistical reporting for correlations and regression results can be found in* ***Table S 9.***

#### Overlap Between our Diagnosis Results and Differential Gene Expression Related to Substances of Abuse

**A.**

| *<u>Pritzker:</u> Percent of subjects with documented or suspected current usage* | CTRL | SCHIZ | BP | *Total n* |
|---|---|---|---|---|
| Tobacco | 11% | 45% | 33% | *n=20* |
| Alcohol (Heavy) | 4% | 36% | 62% | *n=22* |
| Cannabinoid | 0% | 18% | 33% | *n=11* |
| Stimulants | 0% | 14% | 48% | *n=13* |
| Opioids | 0% | 9% | 29% | *n=8* |
| Overdose | 0% | 32% | 38% | *n=15* |

**B.**

| *<u>Iwamoto et al.:</u> Percent of subjects with documented usage* | CTRL | SCHIZ | BP | MDD | *Total n* |
|---|---|---|---|---|---|
| Smoking at Time of Death | 20% | 46% | 45% | 36% | *n=18* |
| Alcohol (Heavy: Current or Past) | 0% | 23% | 18% | 18% | *n=7* |
| Drugs (Heavy: Current or Past) | 0% | 15% | 36% | 9% | *n=7* |

**C.**

| *Correlation between substance usage for different drug categories* | Overdose | Opioids | Stimulants | Cannabinoid | Alcohol | Tobacco |
|---|---|---|---|---|---|---|
| Tobacco | 0.21 | 0.07 | 0.35 | 0.33 | 0.25 | 1.00 |
| Alcohol | 0.54 | 0.24 | 0.47 | 0.55 | 1.00 | |
| Cannabinoid | 0.44 | 0.34 | 0.50 | 1.00 | | |
| Stimulants | 0.46 | 0.52 | 1.00 | | | |
| Opioids | 0.58 | 1.00 | | | | |
| Overdose | 1.00 | | | | | |

**D.**

| *Similarity of the effects (Log2FC) of substance use variables in different datasets for top 20 diagnosis-related genes (correlation: R)* | Iwamoto_Drugs_Heavy | Iwamoto_Alcohol_Heavy | Iwamoto_Smoking | Kapoor_RNASeq_Alcohol_Abuse | Gandal_Microarray_Meta_AAD | Seney_RNASeq_Opioid_Use_Disorder | Pritzker_qPCR_Overdose | Pritzker_qPCR_Opioids | Pritzker_qPCR_Stimulants | Pritzker_qPCR_Cannabinoid | Pritzker_qPCR_Tobacco | Pritzker_qPCR_Alcohol |
|---|---|---|---|---|---|---|---|---|---|---|---|---|
| Pritzker_qPCR_Alcohol | -0.31 | -0.39 | -0.26 | 0.36 | 0.04 | 0.69 | 0.23 | -0.06 | 0.24 | 0.24 | 0.39 | 1.00 |
| Pritzker_qPCR_Tobacco | -0.16 | -0.49 | -0.63 | 0.56 | 0.69 | 0.43 | 0.79 | 0.63 | 0.88 | 0.83 | 1.00 | |
| Pritzker_qPCR_Cannabinoid | -0.07 | -0.57 | -0.61 | 0.23 | 0.53 | 0.47 | 0.86 | 0.79 | 0.89 | 1.00 | | |
| Pritzker_qPCR_Stimulants | 0.01 | -0.44 | -0.49 | 0.32 | 0.56 | 0.26 | 0.77 | 0.71 | 1.00 | | | |
| Pritzker_qPCR_Opioids | 0.04 | -0.38 | -0.45 | -0.03 | 0.36 | 0.79 | 0.93 | 1.00 | | | | |
| Pritzker_qPCR_Overdose | -0.17 | -0.57 | -0.58 | 0.21 | 0.43 | 0.72 | 1.00 | | | | | |
| Seney_RNASeq_Opioid_Use_Disorder | -0.24 | -0.45 | -0.42 | 0.17 | 0.21 | 1.00 | | | | | | |
| Gandal_Microarray_Meta_AAD | -0.12 | -0.38 | -0.68 | 0.53 | 1.00 | | | | | | | |
| Kapoor_RNASeq_Alcohol_Abuse | -0.36 | -0.44 | -0.45 | 1.00 | | | | | | | | |
| Iwamoto_Smoking | 0.34 | 0.78 | 1.00 | | | | | | | | | |
| Iwamoto_Alcohol_Heavy | 0.64 | 1.00 | | | | | | | | | | |
| Iwamoto_Drugs_Heavy | 1.00 | | | | | | | | | | | |

***Figure S 18. Exposure to substances of abuse was common in our diagnosis groups and associated with surprisingly consistent differential expression.***
***A.*** *The percent of subjects in each diagnosis group with documented exposure or suspected current usage for each of the following substances of abuse. Substance exposure was defined by indication of usage within the subjects' clinical records, family interviews, toxicology reports, and coroners reports. Overdose was indicated by the coroner's report, and occurred in response to both drugs of abuse as well as therapeutics. The total n shows the total number of subjects with substance exposure (across diagnosis groups).* ***B.*** *The percent of subjects in each diagnosis group with documented usage for each of the following substances in the Iwamoto et al. BA10 microarray dataset [23] based on metadata accompanying the dataset.* ***C.*** *Polysubstance use: Subjects that have been exposed to one category of substances frequently have also been exposed to other categories of substances within the Pritzker qPCR dataset. Shown is an asymmetrical correlation matrix illustrating the positive correlations (R values) between exposure for different categories of substances within the Pritzker qPCR dataset.* ***D.*** *Within a set of exploratory analyses, we estimated the differential expression (Log2FC) in BA10 in our qPCR dataset associated with a variety of substances of abuse (tobacco, cannabinoids, stimulants, opioids) while controlling for diagnosis. To reduce false discovery due to multiple comparisons, this exploratory analysis was limited to the 20 genes with the most reliable diagnosis effects in BA10 (listed in* ***Figure 3****). The pattern revealed by our exploratory analysis was surprisingly consistent across substances, as illustrated by this asymmetrical correlation matrix showing the correlation between the differential expression (Log2FC) for each substances (positive*

*correlations (R>0) shown in red, negative correlations (R<0 shown in blue). This positive correlation was also observed with the differential expression associated with substances of abuse (Log2FC) in previously published cortical post-mortem datasets (alcohol abuse: microarray meta-analysis* [18]*, RNA-Seq* [19]*; opioid use disorder with overdose: RNA-Seq* [20]*), but was not observed for the effects of substances of abuse within the Iwamoto et al. microarray dataset* [23]*. Full statistical reporting for correlations and regression results can be found in **Table S 9.***

Due to the increased rate of substance usage in the diagnosis group, it seemed important to explore whether the diagnosis-related gene expression that we were observing in BA10 could be better explained by substances of abuse. The pattern revealed in our exploratory analysis was surprisingly consistent across substances, with the effect of a variety of substances of abuse correlating positively with the effect of diagnosis measured in our original model (example in **Fig 4**, full statistical reporting in ***Table S 9,*** *Tobacco Log2FC vs.: BP Log2FC*: R=0.54, p=0.0141, *SCHIZ Log2FC*: R=0.66, p=0.00149; *Cannabinoid Log2FC vs.: BP Log2FC*: R=0.70, p=0.000707, *SCHIZ Log2FC:* R=0.75, p=0.000157; *Stimulant Log2FC vs.: BP Log2FC:* R=0.67, p=0.00122, *SCHIZ Log2FC:* R=0.74, p=0.000189; *Opioid Log2FC vs.: BP Log2FC:* R=0.90, p=5.04e-08, *SCHIZ Log2FC:* R=0.91, p=2.21e-08) as well as death by overdose (*vs. BP Log2FC:* R=0.83, p=5.45e-06, *SCHIZ Log2FC:* R=0.85, p=2.23e-06). Notably, the differential expression associated with substances of abuse in our dataset was not only correlated with our own diagnosis-related differential expression results, but also the diagnosis-related differential expression in other cortical datasets (*e.g., Opioid Log2FC vs.* : *Iwamoto BP Log2FC:* R=0.43; *Gandal RNA-Seq BP Log2FC:* R=0.79, *Iwamoto SCHIZ Log2FC:* R=0.32, *Gandal RNA-Seq SCHIZ Log2FC:* R=0.65; *Overdose Log2FC vs. : Iwamoto BP Log2FC:* R=0.63; *Gandal RNA-Seq BP Log2FC:* R=0.81, *Iwamoto SCHIZ Log2FC:* R=0.35, *Gandal RNA-Seq SCHIZ Log2FC:* R=0.60; *Cannabinoid Log2FC vs. : Iwamoto BP Log2FC:* R=0.71; *Gandal RNA-Seq BP Log2FC:* R=0.69, *Iwamoto SCHIZ Log2FC:* R=0.48, *Gandal RNA-Seq SCHIZ Log2FC:* R=0.57).

#### A. Exploratory: Effect of substances of abuse on gene expression

| | BA10: Diagnosis | | | Substances of Abuse | | | | | | | | | | | |
|---|---|---|---|---|---|---|---|---|---|---|---|---|---|---|---|
| Gene Symbol | Pritzker_qPCR_Bipolar | Pritzker_qPCR_Schiz | Microarray_MetaAnalysis_Schiz | Pritzker_qPCR_Alcohol | Pritzker_qPCR_Cannabinoid | Pritzker_qPCR_Tobacco | Pritzker_qPCR_Stimulants | Pritzker_qPCR_Opioids | Pritzker_qPCR_Overdose | Seney_RNASeq_Opioid_Use_Disorder | Gandal_Microarray_Meta_AAD | Kapoor_RNASeq_Alcohol_Abuse | Iwamoto_LifetimeAlcohol_Heavy | Iwamoto_LifetimeDrugs_Heavy | Iwamoto_Smoking |
| **HTR2B** | **-0.68** | **-0.98** | -0.05 | 0.36 | -0.35 | -0.17 | -0.35 | **-1.73** | **-0.68** | | **-0.17** | **0.10** | 0.02 | -0.18 | 0.12 |
| **SST** | -0.31 | **-0.47** | -0.22 | **-0.33** | **-0.36** | **-0.34** | -0.12 | **-0.70** | **-0.60** | **-0.46** | -0.04 | -0.04 | 0.32 | 0.22 | 0.32 |
| **DRD4** | **-0.47** | **-0.35** | -0.04 | -0.25 | -0.19 | -0.18 | **-0.33** | **-0.66** | **-0.42** | | **0.40** | 0.09 | -0.01 | -0.02 | -0.07 |
| **BDNF** | -0.24 | **-0.35** | -0.10 | -0.16 | 0.02 | -0.15 | 0.09 | **-0.71** | **-0.44** | **-0.59** | -0.10 | 0.02 | -0.04 | 0.03 | 0.05 |
| **DRD2** | -0.14 | **-0.25** | -0.04 | **-0.18** | -0.02 | 0.01 | 0.06 | 0.06 | -0.06 | | 0.10 | | 0.01 | -0.01 | 0.03 |
| **DRD3** | -0.12 | **-0.31** | -0.04 | -0.12 | -0.14 | **-0.29** | -0.14 | -0.33 | -0.30 | | **-0.14** | | -0.01 | -0.08 | 0.05 |
| **HOMER1** | -0.03 | **-0.13** | 0.04 | -0.09 | -0.01 | -0.08 | -0.06 | -0.10 | -0.13 | -0.04 | -0.17 | **-0.11** | 0.19 | 0.04 | 0.18 |
| **SLC6A12** | -0.01 | -0.07 | **-0.16** | -0.06 | **-0.25** | **-0.27** | **-0.23** | **-0.30** | **-0.31** | -0.23 | | -0.07 | 0.04 | -0.13 | 0.02 |
| **COMT** | 0.05 | 0.07 | **-0.13** | -0.02 | 0.00 | **0.21** | 0.20 | -0.06 | 0.02 | -0.01 | 0.08 | **0.14** | -0.12 | -0.15 | 0.06 |
| **SLC6A11** | 0.19 | 0.00 | **-0.05** | -0.10 | 0.11 | 0.03 | -0.03 | **0.50** | **0.36** | 0.09 | -0.14 | 0.02 | -0.06 | -0.12 | 0.07 |
| **GABRB1** | 0.07 | **0.12** | 0.07 | 0.05 | 0.05 | 0.01 | **0.19** | 0.09 | 0.04 | -0.06 | 0.02 | **0.08** | 0.05 | 0.06 | 0.07 |
| **GRM5** | **0.16** | 0.00 | **0.07** | **0.18** | 0.05 | 0.01 | 0.10 | -0.01 | 0.03 | 0.02 | 0.04 | 0.02 | -0.07 | 0.04 | 0.06 |
| **CALB1** | 0.12 | 0.02 | **0.50** | -0.02 | 0.12 | -0.06 | 0.14 | -0.10 | -0.20 | 0.06 | -0.02 | -0.07 | 0.06 | 0.25 | 0.07 |
| **ABAT** | **0.16** | 0.08 | 0.06 | -0.04 | **-0.14** | -0.05 | 0.01 | -0.06 | -0.07 | -0.03 | **-0.29** | -0.02 | -0.01 | 0.05 | 0.01 |
| **MAPK1** | **0.14** | 0.09 | 0.06 | 0.06 | **0.10** | 0.01 | **0.15** | 0.09 | **0.09** | **0.09** | 0.01 | -0.02 | -0.14 | -0.19 | 0.05 |
| **NSF** | **0.11** | **0.10** | 0.19 | -0.02 | -0.06 | -0.02 | -0.01 | -0.03 | -0.05 | | 0.02 | -0.03 | 0.13 | **0.29** | 0.07 |
| **SNCA** | 0.10 | **0.10** | 0.11 | 0.00 | 0.03 | **-0.10** | -0.01 | 0.13 | 0.08 | 0.12 | 0.05 | 0.00 | **-0.18** | 0.01 | -0.07 |
| **GPHN** | **0.16** | **0.10** | | 0.01 | **-0.10** | -0.02 | 0.03 | -0.09 | **-0.11** | **0.06** | 0.13 | 0.01 | | | |
| **MAOB** | 0.14 | **0.22** | 0.11 | 0.06 | 0.08 | **0.31** | 0.20 | 0.07 | 0.04 | 0.04 | 0.17 | **0.12** | 0.10 | 0.15 | 0.01 |
| **GFAP** | 0.26 | **0.44** | 0.00 | 0.12 | **0.48** | **0.79** | **0.76** | **0.73** | **0.60** | 0.01 | **0.70** | **0.12** | -0.22 | -0.13 | -0.23 |

**Bold & Underline**: FDR<0.10, **Bold**=p<0.05

*Down-Regulated*

Log2FC

*Up-Regulated*

***Figure S 19. Exploratory: The effects of BP and SCHIZ on neurotransmission-related gene expression resemble the effects of substance use.***

*Substance exposure was defined by indication of usage within the subjects' clinical records, family interviews, toxicology reports, and coroners reports. Overdose was indicated by the coroner's report, and occurred in response to both drugs of abuse as well as therapeutics. This exploratory analysis was limited to the 20 genes with the most reliable diagnosis effects in BA10 (**Figure 3**). For several genes, the effects of substance use were particularly large (FDR<0.10) and consistent with evidence from larger studies of substance use (Alcohol Abuse Disorder [18,19] or Opioid Use Disorder[20]) in neighboring frontal cortex: A down-regulation of HTR2B, SST, and BDNF, and an upregulation of GFAP, MAPK1, and GABRB1.* Shown is *a table overviewing the effects of diagnosis and substances of abuse on gene expression (Log2FC) for the 20 genes with the most reliable diagnosis effects in BA10. In general, the table follows the conventions of **Figure 3:** blue=decreased in the diagnosis group or condition, red=increased in the diagnosis group or condition, bold=p<0.05, bold+underline=FDR<0.10. On the far left side of the figure, there is an abbreviated version of the table in **Figure 3**, which illustrates the Log2FC associated with diagnosis from our qPCR dataset and meta-analysis of the Iwamoto et al. [23] and Maycox et al. [24] BA10 microarray studies. The table on the right illustrates **1.** The effects of exposure to substances of abuse (Log2FC) estimated while controlling for diagnosis in our dataset. **2.** The* *published effects of opioid use disorder (Log2FC) in the DLPFC [20] and published effects of alcohol abuse disorder (Log2FC) in the cortex [18] and PFC [19]. **3.** The effects of substances of abuse within an exploratory analysis of the Iwamoto et al. [23] BA10 microarray dataset. Documented usage for each of the following substances was based on metadata accompanying the dataset.*

Importantly, even though the effects of substance usage showed many similarities to the effects of diagnosis, for many genes substance usage appeared to amplify the effect of diagnosis versus explain it **(Figure S 20**). For example, out of the genes that showed the largest effects of opioid usage on gene expression (BDNF, DRD4, HTR2B, SST, SLC6A11, GFAP), only GFAP notably lost the effect of diagnosis when the model included opioid usage (effect of diagnosis: p=0.0691*trend). For the top genes affected by overdose, the effects of diagnosis similarly remained strong after controlling for overdose for SST and DRD4, but were lost for GFAP. For the top genes affected by stimulants, the effect of diagnosis was maintained for GABRB1 after controlling for stimulants, but lost or diminished for MAPK1 and GFAP. These exploratory findings are provocative, suggesting a mechanism by which substance usage might combine with diagnosis to amplify deficits in frontal cortical function, but require further validation in a better-powered independent dataset.

**A. Exploratory: A similar pattern of diagnosis effects is observed after controlling for opioid use**

| | BA10: Diagnosis | | | Model w/ Opioids | | | |
|---|---|---|---|---|---|---|---|
| Gene Symbol | Pritzker_qPCR_Bipolar | Pritzker_qPCR_Schiz | Microarray_MetaAnalysis_Schiz | Pritzker_qPCR_Bipolar_CtrlForOpioids | Pritzker_qPCR_Schiz_CtrlForOpioids | Pritzker_qPCR_Opioids | Seney_RNASeq_Opioid_Use_Disorder |
| HTR2B | **-0.68** | **-0.98** | -0.05 | -0.23 | **-0.90** | **-1.73** | |
| SST | -0.31 | **-0.47** | -0.22 | -0.07 | **-0.38** | **-0.70** | **-0.46** |
| DRD4 | **-0.47** | **-0.35** | -0.04 | **-0.29** | **-0.31** | **-0.66** | |
| BDNF | -0.24 | **-0.35** | -0.10 | -0.01 | **-0.26** | **-0.71** | **-0.59** |
| DRD2 | -0.14 | **-0.25** | -0.04 | -0.09 | **-0.23** | 0.06 | |
| DRD3 | -0.12 | **-0.31** | -0.04 | -0.04 | **-0.27** | -0.33 | |
| HOMER1 | -0.03 | **-0.13** | 0.04 | 0.01 | **-0.11** | -0.10 | -0.04 |
| SLC6A12 | -0.01 | -0.07 | **-0.16** | 0.10 | -0.03 | **-0.30** | -0.23 |
| COMT | 0.05 | 0.07 | **-0.13** | 0.00 | 0.05 | -0.06 | -0.01 |
| SLC6A11 | 0.19 | 0.00 | **-0.05** | 0.03 | -0.09 | **0.50** | 0.09 |
| GABRB1 | 0.07 | **0.12** | 0.07 | 0.05 | **0.11** | 0.09 | -0.06 |
| GRM5 | **0.16** | 0.00 | **0.07** | **0.16** | 0.00 | -0.01 | 0.02 |
| CALB1 | 0.12 | 0.02 | **0.50** | 0.17 | 0.04 | -0.10 | 0.06 |
| ABAT | **0.16** | 0.08 | 0.06 | **0.18** | 0.08 | -0.06 | -0.03 |
| MAPK1 | **0.14** | 0.09 | 0.06 | **0.11** | 0.07 | 0.09 | **0.09** |
| NSF | **0.11** | **0.10** | 0.19 | **0.12** | **0.09** | -0.03 | |
| SNCA | 0.10 | **0.10** | 0.11 | 0.07 | **0.11** | 0.13 | 0.12 |
| GPHN | **0.16** | **0.10** | | **0.19** | **0.11** | -0.09 | **0.06** |
| MAOB | 0.14 | **0.22** | 0.11 | 0.01 | **0.18** | 0.07 | 0.04 |
| GFAP | 0.26 | **0.44** | 0.00 | 0.00 | 0.28 | **0.73** | 0.01 |

**Bold & Underline**: FDR<0.10, **Bold**=p<0.05

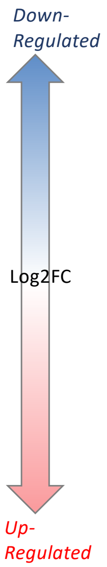

***Figure S 20. Exploratory: A similar pattern of diagnosis effects is observed after controlling for opioid use.***

*Out of the genes that showed the largest effects of opioid usage on gene expression (BDNF, DRD4, HTR2B, SST, SLC6A11, GFAP), only GFAP notably lost the effect of diagnosis when the model included opioid usage (effect of diagnosis: p=0.0691*trend).*
**A)** *A table overviewing the effects of diagnosis on gene expression (Log2FC) in a model with (right) or without (left) controlling for exposure to opioids for the 20 genes with the most reliable diagnosis effects in BA10. In general, the table follows the conventions of* ***Figure 3:*** *blue=decreased in the diagnosis group or condition, red=increased in the diagnosis group or condition, bold=p<0.05, bold+underline=FDR<0.10. On the far left side of the figure, there is an abbreviated version of the table in* ***Figure 3****, which illustrates the Log2FC associated with diagnosis from our qPCR dataset and meta-analysis of the Iwamoto et al.* [23] *and Maycox et al.* [24] *BA10 microarray studies. These analyses did not control for substance use. The table on the right illustrates the same diagnosis effects, but while controlling for opioid exposure in the model. The effects of opioid exposure (Log2FC) in the model is provided as reference, as well as the effects of opioid use disorder (Log2FC) previously documented in the DLPFC in* [20]*.*

***Overlap Between our Diagnosis Results and Symptom-Related Differential Gene Expression***

Since the differential expression in BA10 appeared similar across diagnoses, we would expect that symptoms and related behaviors that are broadly related to the disorders might show a similar profile of differential expression. Therefore, it should perhaps be unsurprising that within our exploratory analyses examining the differential expression associated with particular symptoms and related behaviors in a model controlling for diagnosis, the symptom variable that was best associated with differential expression similar to what we observed for diagnosis in our original model was "Fatigue" (**Figure S 21,** *Fatigue Log2FC vs.: BP Log2FC:* R=0.66, p=0.00142, *SCHIZ Log2FC:* R=0.68, p=0.001). Notably, the differential expression associated with Fatigue in our dataset was not only correlated with our own diagnosis-related differential expression results, but also the diagnosis-related differential expression in other cortical datasets (*e.g., Fatigue Log2FC vs. : Iwamoto BP Log2FC:* R=0.50; *Gandal RNA-Seq BP Log2FC:* R=0.62, *Iwamoto SCHIZ Log2FC:* R=0.36, *Gandal RNA-Seq SCHIZ Log2FC:* R=0.57). For two of our top diagnosis-related genes, the effects of fatigue appeared particularly large, surpassing false detection correction. BDNF showed a large down-regulation in association with Fatigue (Log2FC=-0.466), and controlling for it weakened the down-regulation associated with diagnosis to a nominal trend (p=0.0952) that was only still apparent in SCHIZ (SCHIZ Log2FC=-0.226, BP Log2FC=0.0567). SLC6A11 showed a large upregulation in association with Fatigue (Log2FC=0.365), and controlling for it reversed the direction of effect associated with diagnosis to down-regulation (SCHIZ Log2FC=-0.125, BP Log2FC: -0.0448), neither of which were significant but were now consistent with both the magnitude and direction of nominally-significant effects observed in other datasets (Maycox SCHIZ Log2FC =-0.08, Pritzker Meta-Analysis SCHIZ Log2FC =-0.05, Gandal RNA-Seq SCHIZ Log2FC: -0.10).

**A. Exploratory: Resemblance of the effects of diagnosis to the effects of specific symptoms, depression, and stress**

| Gene Symbol | BA10: Diagnosis | | | BA10: Symptoms Resembling Diagnosis | | | | | MDD | | Stress |
|---|---|---|---|---|---|---|---|---|---|---|---|
| | Pritzker_qPCR_Bipolar | Pritzker_qPCR_Schiz | Microarray_MetaAnalysis_Schiz | Pritzker_qPCR_agitated | Pritzker_qPCR_fatigue | Pritzker_qPCR_trouble_concentrating | Pritzker_qPCR_interactions_with_leg | Pritzker_qPCR_reckless | Iwamoto_Microarray | Gandal_Microarray_Meta | Drug_Budger_CorticoSteroids |
| HTR2B | **-0.68** | **-0.98** | -0.05 | **-0.91** | -0.29 | -0.07 | -0.11 | 0.06 | -0.15 | -0.05 | Down (2-/1+) |
| SST | -0.31 | **-0.47** | -0.22 | 0.22 | -0.22 | 0.12 | -0.14 | -0.01 | -0.44 | **-0.25** | **Down (2-/1+)** |
| DRD4 | **-0.47** | **-0.35** | -0.04 | -0.11 | -0.05 | -0.25 | 0.09 | 0.06 | -0.05 | 0.05 | N |
| BDNF | -0.24 | **-0.35** | -0.10 | 0.29 | **-0.47** | 0.16 | -0.18 | 0.10 | -0.02 | **-0.17** | **Down (7-/1+)** |
| DRD2 | -0.14 | **-0.25** | -0.04 | -0.14 | -0.05 | -0.13 | **0.18** | 0.14 | -0.03 | 0.04 | **Down (7-/2+)** |
| DRD3 | -0.12 | **-0.31** | -0.04 | -0.16 | -0.12 | -0.22 | 0.12 | 0.10 | -0.03 | 0.01 | N |
| HOMER1 | -0.03 | **-0.13** | 0.04 | **0.16** | -0.09 | 0.05 | -0.09 | -0.03 | 0.02 | **-0.07** | Down (6-/1+) |
| SLC6A12 | -0.01 | -0.07 | **-0.16** | **-0.28** | -0.08 | 0.04 | -0.01 | -0.11 | 0.05 | -0.04 | N |
| COMT | 0.05 | 0.07 | **-0.13** | -0.07 | -0.12 | -0.01 | 0.11 | 0.05 | -0.01 | 0.01 | Up (4+/1-) |
| SLC6A11 | 0.19 | 0.00 | **-0.05** | 0.11 | **0.36** | 0.10 | 0.01 | -0.06 | 0.02 | 0.02 | **Mixed (1+/1-)** |
| GABRB1 | 0.07 | **0.12** | 0.07 | 0.06 | 0.06 | 0.01 | **0.21** | **0.14** | 0.08 | 0.00 | Up (1+) |
| GRM5 | **0.16** | 0.00 | **0.07** | 0.13 | -0.06 | 0.07 | 0.15 | 0.03 | 0.03 | -0.02 | Up (5+/2-) |
| CALB1 | 0.12 | 0.02 | **0.50** | 0.11 | -0.03 | 0.03 | 0.18 | 0.15 | 0.14 | 0.01 | Up (1+) |
| ABAT | **0.16** | 0.08 | 0.06 | 0.03 | -0.03 | **0.12** | 0.04 | 0.01 | 0.00 | -0.01 | Up (5+/1-) |
| MAPK1 | **0.14** | 0.09 | 0.06 | **0.17** | 0.00 | -0.03 | **0.10** | **0.14** | 0.15 | 0.01 | Up (4+) |
| NSF | **0.11** | **0.10** | 0.19 | **0.10** | 0.06 | 0.02 | 0.02 | **0.14** | -0.05 | -0.01 | Down (2-/1+) |
| SNCA | 0.10 | **0.10** | 0.11 | **0.18** | 0.03 | -0.04 | 0.03 | 0.07 | -0.04 | -0.04 | Mixed (2+/2-) |
| GPHN | **0.16** | **0.10** | | 0.09 | -0.05 | **0.14** | 0.03 | 0.06 | | -0.01 | Up (5+/2-) |
| MAOB | 0.14 | **0.22** | 0.11 | -0.02 | 0.08 | 0.12 | 0.12 | 0.06 | 0.07 | 0.04 | Up (2+/1-) |
| GFAP | 0.26 | **0.44** | 0.00 | -0.16 | 0.22 | -0.24 | 0.31 | -0.04 | 0.24 | -0.03 | Up (1+) |

*Down-Regulated* — Log2FC — *Up-Regulated*

**Log2FC: Bold & Underline**: FDR<0.10, **Bold**=p<0.05

**DrugBudger:** # of results (FDR<0.05) with each direction of effect

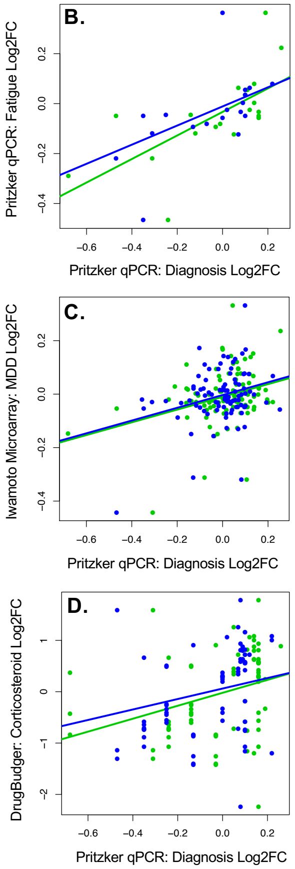

***Figure S 21. Exploratory: The effects of BP and SCHIZ on neurotransmission-related gene expression resemble the effects of fatigue, executive dysfunction, and stress.***
***A)*** *A table overviewing the effects of diagnosis, diagnosis-related symptoms and related behaviors, and stress on gene expression (Log2FC) for the 20 genes with the most reliable diagnosis effects in BA10. In general, the table follows the conventions of* ***Figure 3:*** *blue=decreased in the diagnosis group or condition, red=increased in the diagnosis group or condition, bold=p<0.05, bold+underline=FDR<0.10. On the far left side of the figure, there is an abbreviated version of the table in* ***Figure 3****, which illustrates the Log2FC associated with diagnosis from our qPCR dataset and meta-analysis of the Iwamoto et al.* [23] *and Maycox et al.* [24] *BA10 microarray studies. The three tables on the right illustrate* ***1.*** *The effects of diagnosis-related symptoms and related behaviors (Log2FC) estimated while controlling for diagnosis in our dataset. Symptoms and related behaviors were documented with subjects' clinical records and family interviews. Only the results from the five symptoms and related behaviors with sufficient sample size (>6/subgroup) and significant (FDR<0.10) associations with gene expression that most resemble diagnosis are shown.* ***2.*** *The effects of Major Depressive Disorder (Log2FC) within the Iwamoto et al.* [23] *BA10 microarray dataset and the large Gandal et al.* [18] *cortical microarray meta-analysis.* ***3.*** *The effects of corticosteroids on gene expression within the Drug Gene Budger database* [22]*. Drug Gene Budger includes results from both in vitro and in vivo experiments. The ratio of the number of effects in the database that indicate upregulation (+) versus downregulation (-) with drug exposure are indicated in parentheses, regardless of dosage or experiment of origin. N means "not present in database". All effects in the database were significant (FDR<0.05).* ***B-D)*** *Scatterplots illustrating the positive correlation between the differential expression associated with diagnosis-related symptoms, depression*

*(MDD), or stress and the differential expression (Log2FC) associated with diagnosis in our qPCR study (x-axis: Blue=Bipolar Disorder Log2FC, Green=Schizophrenia Log2FC).* ***B.*** *The effects of diagnosis (Log2FC) on gene expression in BA10 measured in our qPCR study correlated positively with the effect of fatigue as estimated while controlling for diagnosis in our dataset (BP: p=0.00142, SCHIZ: p=0.001). Fatigue was defined by indication that the symptom was present within the subjects' clinical records and interview with family members. This exploratory analysis was limited to the 20 genes with the most reliable diagnosis effects in BA10 (panel A).* ***C.*** *The effects of diagnosis (Log2FC) on gene expression in BA10 measured in our qPCR study correlated positively with the effect of Major Depressive Disorder (Log2FC) on gene expression in BA10 as measured by microarray (reanalyzed by our lab, [23], SCHIZ: p=0.000234; BP: p=0.00193). This exploratory analysis included all neurotransmission-related genes in our qPCR dataset****. D.*** *The effects of diagnosis (Log2FC) on gene expression in BA10 measured in our qPCR study correlated positively with the effects of corticosteroids on gene expression within the Drug Gene Budger database [22] (BP: p=0.0172; SCHIZ: p=0.0136). This exploratory analysis was limited to the 20 genes with the most reliable diagnosis effects in BA10 (panel A). Genes with at least two documented effects of stress within the Stress Mice Portal database [43] are highlighted in bold. Full statistical reporting for correlations and regression results can be found in* ***Table S 9.***

A strong relationship was also observed for "Disorganized Speech" (*Disorganized Speech Log2FC vs.: BP Log2FC:* R=0.82, *SCHIZ Log2FC:* R=0.85), but upon further review this variable had a very small sample size (n=6), and may be considered untrustworthy. This was followed by "Decreased worth and helplessness", which showed similar differential expression as Diagnosis within our dataset (*Decreased worth and helplessness Log2FC vs.: BP Log2FC:* R=0.72, *SCHIZ Log2FC:* R=0.65), but this was not replicated when compared to the diagnosis-related differential expression in other datasets (e.g., *Iwamoto, Gandal RNA-Seq:* all R<0.06). *"Changes in weight"* also showed similar differential expression as Diagnosis in our dataset (*"Changes in weight" Log2FC vs.: BP Log2FC:* R=0.65, *SCHIZ Log2FC:* R=0.64*),* which was mirrored when compared to the effects of BP within the Gandal RNA-Seq dataset (R=0.64), but was not as strongly observed when comparing with effects of diagnosis measured in other datasets *(e.g., Iwamoto, Gandal RNA-Seq*: all R<0.25*).* In contrast, "Interactions with the Legal System" was associated with differential expression that was similar to the effects of diagnosis in our dataset (**Figure S 21,** *"Interactions with the Legal System" Log2FC vs.: BP Log2FC:* R=0.53*, SCHIZ Log2FC:* R=0.62*)* which was strongly replicated when comparing with diagnosis-related gene expression in other datasets (*e.g., "Interactions with the Legal System" Log2FC vs.: Iwamoto BP Log2FC:* R=0.62; *Gandal RNA-Seq BP Log2FC:* R=0.84, *Iwamoto SCHIZ Log2FC:* R=0.51, *Gandal RNA-Seq SCHIZ Log2FC:* R=0.92). This relationship could be mediated by usage of substances of abuse (discussed earlier) which showed a pattern of differential expression more similar to diagnosis within our dataset. The effect of *"Interactions with the Legal System"* appeared particularly large one of our top diagnosis-related genes, GABRB1, which showed an upregulation (Log2FC=0.206) that surpassed false detection correction. When controlling for *"Interactions with the Legal System",* the upregulation associated with diagnosis for GABRB1 was weakened and no longer nominally significant (*BP:* Log2FC=0.00600*, SCHIZ:* Log2FC=0.0622*).*

Also notable were symptom-related variables that did *not* show a pattern of differential expression similar to what we observed for diagnosis. This included affect-related variables, such as "Depressed Mood", "Anxiety", and "Suicide" (all R<-0.29), "Negative Sxs", "Disorganized Behavior Or Catatonic", and other cognitive variables like "Difficulty Concentrating" (all R<-0.37).

Although clearly preliminary, the differential expression associated with symptom-related variables in our exploratory analyses suggest that the differential expression within BA10 and neighboring cortex associated with Bipolar Disorder and Schizophrenia may be more broadly related to diminished self-control (substances of abuse, interactions with the legal system) and fatigue than with disturbances in affect, cognition, or many of the more canonical symptoms associated with the disorders. That said, since the exploratory analyses were run while controlling for diagnosis, it is also possible that we were not well-powered to detect relationships with the symptoms that are most canonical to defining our diagnoses.

***Overlap Between our Diagnosis Results and Stress-Related Differential Gene Expression***

Another issue that arises while interpreting the effects of diagnosis within human post-mortem studies is the question of whether the observed effects are due to the illness itself or due to the stress that accompanies and often amplifies the illness. To gain preliminary insight into this relationship, we outputted the effects of corticosteroids on gene expression documented in the Drug Gene Budger database using the same protocol discussed above. The results showed a remarkable parallel to our diagnosis findings: When considering the full results from all corticosteroids, we found again that the effects of corticosteroids on gene expression were similar to the effects of diagnosis on gene expression in BA10 (**Figure S 21,** n=74 effects, with gene included in the model as a random effect: *Bipolar:* p=0.0172; *Schiz:* p=0.0136).

As a follow-up, we also examined the overlap between our top genes and genes repeatedly implicated within RNA-Seq studies examining the effect of stress on brain gene expression in mice as documented through the Stress Mice Portal (http://hpc-bioinformatics.cineca.it/stress_mice/, [43], accessed 09/2021). Within the database, 1925 unique genes were found to be differentially expressed in at least two stress-related RNA-Seq Bioprojects (out of 18), 27 of which were targets included in our qPCR experiment. Of these, four were amongst our top 20 differentially expressed genes: SST, BDNF, DRD2, and SLC6A11. These results included findings derived from frontal cortical tissue (PFC), indicating a down-regulation of BDNF [46], SST [46], and DRD2 in response to chronic stress [47] similar to what we observed in association with diagnosis.

***Overlap Between our Diagnosis Results and Genetic Variants Associated With Schizophrenia***

To determine whether any of our top differentially expressed genes might be related to genetic variants conferring risk for Schizophrenia, we compared our list of top genes to SZDB2.0, a large database of Schizophrenia genetic research ([41,42], accessed 09/2021). To be included in this comparison, a gene needed to show strong evidence of differential expression in BA10: either FDR<0.10 in our qPCR experiment (gene symbol with grey shading) or p<0.05 (Log2FC in bold text) and consistent direction of effect in two independent datasets from BA10 or BA10 and DLPFC/frontal cortex (20 genes total, **Figure 3**). Within the SZDB2.0 database, there were 571 unique genes implicated by large genome-wide association studies (CLOZUK, PGC2; [48]), seven of which were included as targets within our qPCR experiment, and one of which showed down-regulation in association with diagnosis in BA10 (DRD2). Notably, despite being an important target of both typical and atypical antipsychotic therapeutics, DRD2 has such low-level expression in the cortex that it was deemed undetectable in previous RNA-Seq experiments [18]. We were able to observe down-regulation of DRD2 in association with Schizophrenia in BA10 using the more sensitive methodology of qPCR. Down-regulation of DRD2 in association with Schizophrenia was also observed in an earlier microarray experiment performed using grey-matter focused dissection ([49], as re-analyzed by [13]).

Within the SZDB2.0 database, there were also 408 unique genes within genetic loci with copy number variation (CNV) associated with Schizophrenia [50], only one of which was included as a target in our qPCR study (COMT). We did not observe differential expression of COMT in our qPCR study, but down-regulation of COMT in association with Schizophrenia was observed within both of the BA10 microarray datasets that we re-analyzed [23,24], as well as in our meta-analysis of their effects.

There was also noisier evidence within SZDB2.0 that may suggest that some of our top differentially expressed genes are related to genetic susceptibility to Schizophrenia. Within SZDB2.0, there were 5830 unique genes implicated in association with Schizophrenia by exome sequencing studies, 45 of which were included as qPCR targets. Four of these genes contained schizophrenia-associated variants predicted to be missense/damaging or stop-gained (HTR2B [51], DRD2 [52], DRD3 [51], SLC6A12 [52]) and showed down-regulation in association with diagnosis in BA10, whereas MAOB had a variant predicted to affect protein-protein contact [51] and was upregulated in BA10 in association with diagnosis. Again, several of these genes (HTR2B, DRD2, DRD3) have such low-level expression in the cortex that that they were deemed undetectable in previous RNA-Seq experiments [18], meaning that our qPCR experiment provides novel insight into their differential expression.

Thirty of our qPCR targets were also found within regions implicated by linkage or association studies. Five of these genes were down-regulated in BA10 in association with SCHIZ (DRD4, DRD3, DRD2, BDNF, COMT) and implicated by both a meta-analysis of linkage studies [53,54] and a meta-analysis of association studies [55,56]. Six of our targets were also implicated by differential methylation [57], one of which was down-regulated in BA10 in association with Schizophrenia (BDNF). However the evidence provided from linkage association and differential methylation studies is more difficult to interpret because the overall number of associations present in the SZDB 2.0 database was not readily available.

#### Supplemental Table Legends

***Table S 1. Key Resources Table.***

***Table S 2. Important subject demographics and tissue sample quality metrics.***
*This .xlsx file contains the demographics for all subjects as well as the tissue and RNA quality metrics for their respective samples. Columns include subject ID, brain bank cohort, diagnosis, age (yrs), gender, brain pH, agonal factor score, post-mortem interval (PMI, hrs), secondary dissection/ RNA extraction group, concentration of the purified RNA (ng/ul, averaged from two separate measurements), purity of the purified RNA (260/280 and 260/230, averaged from two separate measurements), and RNA integrity (RIN, 28s/18s rRNA ratio). The final column indicates whether the data from each subject was excluded during data analysis due to failing basic quality control (poor RNA integrity or low purity/concentration).*

***Table S 3. The full list of genes represented in the two qPCR datasets along with their average Cq and rate of missing or low quality measurements ("NA").***
*This .xlsx file contains two spreadsheets named "GABA-GLU dataset" and "DA-5HT dataset". Within each spreadsheet the first column contains the official gene symbol for each gene ("GeneSymbol"), the second column contains the average Cq values for each gene ("Average Cq"), the third column contains the number of missing or low-quality (NA) measurements for each gene ("Number NA"), the fourth column contains the category ("Target Gene" vs. "Reference Gene"), and fifth column indicates whether the gene was excluded from the dataset due to failing quality control. The average Cq and number of missing or low-quality measurements were quantified in the dataset following the initial exclusion of data from three subjects due to poor RNA quality metrics, as well as two replicate samples within the GabaGlu dataset that failed amplification.*

***Table S 4. Balanced design: demographics, tissue and RNA quality by diagnosis.***
*This table includes descriptive statistics (mean+/-sd, range) for each diagnosis group for important biological (age, pH, PMI) and technical (block weight, RNA concentration, RNA purity, RNA integrity) variables. The effect of diagnosis was also assessed using ANOVA, followed by an assessment of the effect of each individual diagnosis (BP, SCHIZ), as derived from a linear regression analysis. These statistics were calculated following the exclusion of three subjects with poor RNA metrics but prior to the imputation of missing data (one subject with missing pH, two subjects with missing RNA metrics).*

***Table S 5. The full concatenated results for the effect of diagnosis and the biological co-variates on gene expression in both qPCR datasets.***
*This .xlsx file contains two spreadsheets. The first spreadsheet ("Definitions") contains an explanation of the statistical methods and the definitions for all abbreviations used. The second spreadsheet ("AllTargetGenes") contains the output associated with all of the fixed effects in a multilevel model containing Diagnosis as well as the biological co-variates Age, Gender, Brain pH, and PMI, and the technical co-variate of qPCR Card (Equation 3). The first column provides the official gene symbol ("GeneSymbol"). Columns B-E contain the subject-level sample size, sample-level sample size, and degrees of freedom ("DF") associated with each of the variables in the model. Columns G-AG contain the output derived from the Type III Wald test comparing the full and reduced model for each fixed effect variable in the model produced by the Anova function, including the Chi-Square statistic ("ChiSquare"), p-value ("Pval"), and false discovery rate ("FDR"). Columns AH-CG contain the output derived from the post-hoc test to extract the*

*results for the individual coefficients for each level of the variables, including the degrees of freedom ("DF"), coefficient ("Beta") and associated standard error ("SE"), T-statistic ("Tstat"), p-value ("Pval"), and false discovery rate ("FDR"). Please note that for the sake of conciseness in this output we did not include the individual coefficients associated with each qPCR card. The third worksheet ("Simplified_JustDiagnosis_Sorted") contains a simplified summary of the same statistical results, focused only on the variable Diagnosis. The output in this worksheet has been sorted by the Diagnosis p-value derived from the Type III Wald test performed by the Anova function.*

***Table S 6. The results from an analysis exploring the sensitivity of the estimation of the effect of diagnosis on gene expression to model specification.***

*This .xlsx file contains five spreadsheets. The first spreadsheet ("Definitions") contains an explanation of the statistical methods and the definitions for all abbreviations used. The next four spreadsheets contain the statistical output for each diagnosis (Schizophrenia or Bipolar Disorder) for all genes included in each qPCR experiment (GABA-GLU or DA5HT) using a variety of model specifications. The first column in each spreadsheet provides the official gene symbol ("GeneSymbol"), the second column contains the category ("Target Gene" vs. "Reference Gene"). Columns C-J contain the effect of each diagnosis (Beta, Log(2) Fold Change, or -ΔΔCq) on the normalized gene expression measurements (-ΔCq) for each gene, as evaluated within each specified multilevel model using the function lme (package nlme v3.1-131 (Pinheiro et al., 2020)) and maximum log-likelihood. Columns K&L provide the minimum and maximum Beta (Log(2) Fold Change) across all models, respectively. Columns M-U contain the nominal p-values associated with those effects, and columns V&W provide the minimum and maximum nominal p-value across all models, respectively.*

***Table S 7. Previously Published BA10 Microarray Results.***

*This .xlsx file contains two spreadsheets. The first spreadsheet ("qPCRGenes_WPreviousEffects") shows the results for all genes (gene symbol) in our qPCR experiment that had been mentioned in the main text or supplementary material of previous publications as having differential expression in BA10 in relationship to BP or SCHIZ diagnosis* [23–26]*. Our qPCR results for those genes are shown for reference (full results and definitions in* ***Table S 5****). To avoid copyright infringement, the results from previous publications are simply annotated with "Up" or "Down" to indicate increased or decreased expression in the diagnosis group, respectively. The full database is available upon request. The second spreadsheet ("GenesWMultiplePreviousEffects") shows the results for all genes (gene symbol) that were mentioned in more than one of the previous publications as having differential expression in association with BP or SCHIZ diagnosis.*

*At first glance, our qPCR differential expression results didn't overlap much with the differential expression results reported in these studies: Out of the 14 target genes from our study that had effects of SCHIZ or BP diagnosis in at least one of these previous studies, we were only able to validate the down-regulation of BDNF in SCHIZ (*[24]*, p=2.86E-02 in our dataset). However, there was also substantial disagreement between the results of the published studies: of the 34 genes with diagnosis effects in more than on study, only 13 showed effects in the same direction in both studies. This led us to suspect that the lack of overlap between our study and previous studies might be due to a combination of low power, outdated annotation, and differences in analysis methods, which inspired us to perform a meta-analysis of the publicly available BA10 microarray data* [23,24]*.*

***Table S 8. Results From Our Re-Analysis of BA10 Microarray Studies and BA10 Microarray Meta-Analysis***

*This .xlsx file contains two spreadsheets. The first spreadsheet ("IwamotoVsMaycox_AllResults") contains the full differential expression output for our re-analysis of BA10 microarray studies by Iwamoto et al. [23] and Maycox et al. [24], joined by EntrezID (with official Gene Symbol annotation). The spreadsheet includes the results for all variables that were included in the model, including co-variates, but the results for each diagnosis (BP, SCHIZ, MDD) are highlighted to make them easier to find using conditional formatting akin to other tables in the main text (Log2FC: blue=down-regulated, red=upregulated, pval/FDR: green=more significant, red=less significant). Statistical abbreviations are the same as in **Table S 5**, with the addition of "A", which represents the average log2 expression over all arrays. Covariate definitions are in the supplemental methods. The second spreadsheet ("MetaAnalysisResults") contains the output from our meta-analysis of the effects of SCHIZ from the re-analyzed Iwamoto et al. and Maycox et al. datasets for each EntrezID (with official Gene Symbol annotation). The estimated Log2FC is called "b", standard error is "SE", nominal p-value is "pval", and false discovery rate is "FDR". The spreadsheet is ordered by p-value, with the most significant results listed first. Conditional formatting is similar to the first spreadsheet.*

***Table S 9. Full statistical reporting for correlations between the differential expression associated with different diagnoses, variables, datasets and conditions.***

*This .xlsx file contains two spreadsheets. The first spreadsheet ("ReplicationOfEffects") provides the correlation (R) and regression and/or multilevel model statistics (Beta, SE, T-stat, p-value) for the comparison of differential expression (Log2FC) identified in our qPCR dataset to the differential expression (Log2FC) identified in other datasets [18,19,21–24,44] for the same (or similar) variables. These variables include diagnosis (BP, SCHIZ), co-variates (pH, Age, RIN), and exploratory variables (opioid exposure, overdose, alcohol abuse, tobacco, suicide, antipsychotics). The second spreadsheet ("CrossDiagnosisComparisons") provides the correlation (R) and regression statistics (Beta, SE, T-stat, p-value) for the comparison of diagnosis-related differential expression (Log2FC) identified in our qPCR dataset to differential expression (Log2FC) associated with a different diagnosis in our own and other datasets [18,23,24,36,49]. The third spreadsheet ("Exploratory_vsDiagnosis") provides the correlation (R) and regression and/or multilevel model statistics (Beta, SE, T-stat, p-value) for the comparison of differential expression (Log2FC) for exploratory variables (therapeutics, substances of abuse, symptoms and related behaviors, [18,20–22]) to the differential expression (Log2FC) associated with diagnosis [18,23]. For the sake of conciseness, statistics are provided only for the exploratory variables discussed in the text because of the similarity of their effects to diagnosis or because the relationship with diagnosis was shown to replicate using independent datasets.*

#### References

1 Li JZ, Meng F, Tsavaler L, Evans SJ, Choudary PV, Tomita H *et al.* Sample matching by inferred agonal stress in gene expression analyses of the brain. *BMC Genomics* 2007; **8**: 336.

2 Tomita H, Vawter MP, Walsh DM, Evans SJ, Choudary PV, Li J *et al.* Effect of agonal and postmortem factors on gene expression profile: quality control in microarray analyses of postmortem human brain. *Biol Psychiatry* 2004; **55**: 346–352.

3 Vawter MP, Tomita H, Meng F, Bolstad B, Li J, Evans S *et al.* Mitochondrial-related gene expression changes are sensitive to agonal-pH state: implications for brain disorders. *Mol Psychiatry* 2006; **11**: 615, 663–679.

4 American Psychiatric Association, American Psychiatric Association (eds.). *Diagnostic and statistical manual of mental disorders: DSM-5*. 5th ed. American Psychiatric Association: Washington, D.C, 2013.

5 Kato T, Murashita J, Kamiya A, Shioiri T, Kato N, Inubushi T. Decreased brain intracellular pH measured by P-31-MRS in bipolar disorder: a confirmation in drug-free patients and correlation with white matter hyperintensity. *Eur Arch Psych Clin Neurosci* 1998; **248**: 301–306.

6 Hamakawa H, Murashita J, Yamada N, Inubushi T, Kato N, Kato T. Reduced intracellular pH in the basal ganglia and whole brain measured by P-31-MRS in bipolar disorder. *Psychiatry Clin Neurosci* 2004; **58**: 82–88.

7 Johnson CP, Follmer RL, Oguz I, Warren LA, Christensen GE, Fiedorowicz JG *et al.* Brain abnormalities in bipolar disorder detected by quantitative T1 rho mapping. *Mol Psychiatr* 2015; **20**: 201–206.

8 Sequeira PA, Martin MV, Vawter MP. The first decade and beyond of transcriptional profiling in schizophrenia. *Neurobiology of Disease* 2012; **45**: 23–36.

9 Livak KJ, Schmittgen TD. Analysis of relative gene expression data using real-time quantitative PCR and the 2(-Delta Delta C(T)) Method. *Methods* 2001; **25**: 402–408.

10 Yuan JS, Reed A, Chen F, Stewart CN. Statistical analysis of real-time PCR data. *BMC Bioinformatics* 2006; **7**: 85.

11 Kuznetsova A, Brockhoff PB, Christensen RHB. lmerTest Package: Tests in Linear Mixed Effects Models. *Journal of Statistical Software* 2017; **82**: 1–26.

12 Atz M, Walsh D, Cartagena P, Li J, Evans S, Choudary P *et al.* Methodological considerations for gene expression profiling of human brain. *J Neurosci Methods* 2007; **163**: 295–309.

13 Hagenauer MH, Schulmann A, Li JZ, Vawter MP, Walsh DM, Thompson RC *et al.* Inference of cell type content from human brain transcriptomic datasets illuminates the effects of age, manner of death, dissection, and psychiatric diagnosis. *PLoS ONE* 2018; **13**: e0200003.

14 Bates D, Mächler M, Bolker B, Walker S. Fitting Linear Mixed-Effects Models Using lme4. *Journal of Statistical Software* 2015; **67**: 1–48.

15 Fox, J, Weisberg, S. *An R Companion to Applied Regression*. Third. Sage: Thousand Oaks, CA, 2019https://socialsciences.mcmaster.ca/jfox/Books/Companion/.

16 Pollard KS, Dudoit S, Laan MJ van der. Multiple Testing Procedures: the multtest Package and Applications to Genomics. In: *Bioinformatics and Computational Biology Solutions Using R and Bioconductor*. Springer, New York, NY, 2005, pp 249–271.

17 Pinheiro J, Bates D, DebRoy S, Sarkar D, R Core Team. *nlme: Linear and Nonlinear Mixed Effects Models*. 2020https://CRAN.R-project.org/package=nlme (accessed 30 Jul2020).

18 Gandal MJ, Haney JR, Parikshak NN, Leppa V, Ramaswami G, Hartl C *et al.* Shared molecular neuropathology across major psychiatric disorders parallels polygenic overlap. *Science* 2018; **359**: 693–697.

19 Kapoor M, Wang J-C, Farris SP, Liu Y, McClintick J, Gupta I *et al.* Analysis of whole genome-transcriptomic organization in brain to identify genes associated with alcoholism. *Transl Psychiatry* 2019; **9**: 89.

20 Seney ML, Kim S-M, Glausier JR, Hildebrand MA, Xue X, Zong W *et al.* Transcriptional Alterations in Dorsolateral Prefrontal Cortex and Nucleus Accumbens Implicate Neuroinflammation and Synaptic Remodeling in Opioid Use Disorder. *Biol Psychiatry* 2021; **90**: 550–562.

21 Martin MV, Mirnics K, Nisenbaum LK, Vawter MP. Olanzapine Reversed Brain Gene Expression Changes Induced by Phencyclidine Treatment in Non-Human Primates. *Mol Neuropsychiatry* 2015; **1**: 82–93.

22 Wang Z, He E, Sani K, Jagodnik KM, Silverstein MC, Ma'ayan A. Drug Gene Budger (DGB): an application for ranking drugs to modulate a specific gene based on transcriptomic signatures. *Bioinformatics* 2019; **35**: 1247–1248.

23 Iwamoto K, Kakiuchi C, Bundo M, Ikeda K, Kato T. Molecular characterization of bipolar disorder by comparing gene expression profiles of postmortem brains of major mental disorders. *Mol Psychiatry* 2004; **9**: 406–416.

24 Maycox PR, Kelly F, Taylor A, Bates S, Reid J, Logendra R *et al.* Analysis of gene expression in two large schizophrenia cohorts identifies multiple changes associated with nerve terminal function. *Mol Psychiatry* 2009; **14**: 1083–1094.

25 Scarr E, Udawela M, Dean B. Changed frontal pole gene expression suggest altered interplay between neurotransmitter, developmental, and inflammatory pathways in schizophrenia. *NPJ Schizophr* 2018; **4**: 4.

26 Scarr E, Udawela M, Dean B. Changed cortical risk gene expression in major depression and shared changes in cortical gene expression between major depression and bipolar disorders. *Aust N Z J Psychiatry* 2019; **53**: 1189–1198.

27 Davis S, Meltzer PS. GEOquery: a bridge between the Gene Expression Omnibus (GEO) and BioConductor. *Bioinformatics* 2007; **23**: 1846–1847.

28 Irizarry RA, Hobbs B, Collin F, Beazer-Barclay YD, Antonellis KJ, Scherf U *et al.* Exploration, normalization, and summaries of high density oligonucleotide array probe level data. *Biostatistics* 2003; **4**: 249–264.

29 Dai M, Wang P, Boyd AD, Kostov G, Athey B, Jones EG *et al.* Evolving gene/transcript definitions significantly alter the interpretation of GeneChip data. *Nucleic Acids Res* 2005; **33**: e175.

30 Fasold M, Binder H. AffyRNADegradation: control and correction of RNA quality effects in GeneChip expression data. *Bioinformatics* 2013; **29**: 129–131.

31 Toker L, Feng M, Pavlidis P. Whose sample is it anyway? Widespread misannotation of samples in transcriptomics studies. *F1000Res* 2016; **5**: 2103.

32 Ritchie ME, Phipson B, Wu D, Hu Y, Law CW, Shi W *et al.* limma powers differential expression analyses for RNA-sequencing and microarray studies. *Nucleic Acids Res* 2015; **43**: e47.

33 Viechtbauer W. Conducting Meta-Analyses in R with the metafor Package. *Journal of Statistical Software* 2010; **36**: 1–48.

34 Carlson M. *org.Hs.eg.db: Genome wide annotation for Human.* 2019https://bioconductor.org/packages/release/data/annotation/html/org.Hs.eg.db.html.

35 Lanz TA, Joshi JJ, Reinhart V, Johnson K, Grantham LE, Volfson D. STEP levels are unchanged in pre-frontal cortex and associative striatum in post-mortem human brain samples from subjects with schizophrenia, bipolar disorder and major depressive disorder. *PLoS ONE* 2015; **10**: e0121744.

36 Narayan S, Tang B, Head SR, Gilmartin TJ, Sutcliffe JG, Dean B *et al.* Molecular profiles of schizophrenia in the CNS at different stages of illness. *Brain Res* 2008; **1239**: 235–248.

37 Reinhart V, Bove SE, Volfson D, Lewis DA, Kleiman RJ, Lanz TA. Evaluation of TrkB and BDNF transcripts in prefrontal cortex, hippocampus, and striatum from subjects with schizophrenia, bipolar disorder, and major depressive disorder. *Neurobiol Dis* 2015; **77**: 220–227.

38 Chang L-C, Jamain S, Lin C-W, Rujescu D, Tseng GC, Sibille E. A conserved BDNF, glutamate- and GABA-enriched gene module related to human depression identified by coexpression meta-analysis and DNA variant genome-wide association studies. *PLoS One* 2014; **9**: e90980.

39 Ponomarev I, Wang S, Zhang L, Harris RA, Mayfield RD. Gene coexpression networks in human brain identify epigenetic modifications in alcohol dependence. *J Neurosci* 2012; **32**: 1884–1897.

40 Bult CJ, Blake JA, Smith CL, Kadin JA, Richardson JE, Mouse Genome Database Group. Mouse Genome Database (MGD) 2019. *Nucleic Acids Res* 2019; **47**: D801–D806.

41 Wu Y, Yao Y-G, Luo X-J. SZDB: A Database for Schizophrenia Genetic Research. *Schizophr Bull* 2017; **43**: 459–471.

42 Wu Y, Li X, Liu J, Luo X-J, Yao Y-G. SZDB2.0: an updated comprehensive resource for schizophrenia research. *Hum Genet* 2020; **139**: 1285–1297.

43 Flati T, Gioiosa S, Chillemi G, Mele A, Oliverio A, Mannironi C *et al.* A gene expression atlas for different kinds of stress in the mouse brain. *Sci Data* 2020; **7**: 437.

44 Seney ML, Huo Z, Cahill K, French L, Puralewski R, Zhang J *et al.* Opposite Molecular Signatures of Depression in Men and Women. *Biol Psychiatry* 2018; **84**: 18–27.

45 Fromer M, Roussos P, Sieberts SK, Johnson JS, Kavanagh DH, Perumal TM *et al.* Gene expression elucidates functional impact of polygenic risk for schizophrenia. *Nat Neurosci* 2016; **19**: 1442–1453.

46 Labonté B, Engmann O, Purushothaman I, Menard C, Wang J, Tan C *et al.* Sex-specific transcriptional signatures in human depression. *Nat Med* 2017; **23**: 1102–1111.

47 Laine MA, Trontti K, Misiewicz Z, Sokolowska E, Kulesskaya N, Heikkinen A *et al.* Genetic Control of Myelin Plasticity after Chronic Psychosocial Stress. *eNeuro* 2018; **5**: ENEURO.0166-18.2018.

48 Pardiñas AF, Holmans P, Pocklington AJ, Escott-Price V, Ripke S, Carrera N *et al.* Common schizophrenia alleles are enriched in mutation-intolerant genes and in regions under strong background selection. *Nat Genet* 2018; **50**: 381–389.

49 Lanz TA, Reinhart V, Sheehan MJ, Rizzo SJS, Bove SE, James LC *et al.* Postmortem transcriptional profiling reveals widespread increase in inflammation in schizophrenia: a comparison of prefrontal cortex, striatum, and hippocampus among matched tetrads of controls with subjects diagnosed with schizophrenia, bipolar or major depressive disorder. *Transl Psychiatry* 2019; **9**: 151.

50 Marshall CR, Howrigan DP, Merico D, Thiruvahindrapuram B, Wu W, Greer DS *et al.* Contribution of copy number variants to schizophrenia from a genome-wide study of 41,321 subjects. *Nat Genet* 2017; **49**: 27–35.

51 Genovese G, Fromer M, Stahl EA, Ruderfer DM, Chambert K, Landén M *et al.* Increased burden of ultra-rare protein-altering variants among 4,877 individuals with schizophrenia. *Nat Neurosci* 2016; **19**: 1433–1441.

52 Howrigan DP, Rose SA, Samocha KE, Fromer M, Cerrato F, Chen WJ *et al.* Exome sequencing in schizophrenia-affected parent-offspring trios reveals risk conferred by protein-coding de novo mutations. *Nat Neurosci* 2020; **23**: 185–193.

53 Lewis CM, Levinson DF, Wise LH, DeLisi LE, Straub RE, Hovatta I *et al.* Genome scan meta-analysis of schizophrenia and bipolar disorder, part II: Schizophrenia. *Am J Hum Genet* 2003; **73**: 34–48.

54 Ng MYM, Levinson DF, Faraone SV, Suarez BK, DeLisi LE, Arinami T *et al.* Meta-analysis of 32 genome-wide linkage studies of schizophrenia. *Mol Psychiatry* 2009; **14**: 774–785.

55 Allen NC, Bagade S, McQueen MB, Ioannidis JPA, Kavvoura FK, Khoury MJ *et al.* Systematic meta-analyses and field synopsis of genetic association studies in schizophrenia: the SzGene database. *Nat Genet* 2008; **40**: 827–834.

56 Sun J, Kuo P-H, Riley BP, Kendler KS, Zhao Z. Candidate genes for schizophrenia: a survey of association studies and gene ranking. *Am J Med Genet B Neuropsychiatr Genet* 2008; **147B**: 1173–1181.

57 Jaffe AE, Gao Y, Deep-Soboslay A, Tao R, Hyde TM, Weinberger DR *et al.* Mapping DNA methylation across development, genotype and schizophrenia in the human frontal cortex. *Nat Neurosci* 2016; **19**: 40–47.
